# An interdisciplinary, randomized, single-blind evaluation of state-of-the-art large language models for their implications and risks in medical diagnosis and management

**DOI:** 10.1101/2025.06.20.25326623

**Authors:** Peikai Chen, Jifu Cai, Jiaying Zhou, Shaoxi Chen, Chenguang Xu, Lihua Yuan, Xiaoying Dai, Xiaowei Chen, Yanzhe Wei, Xia Li, Shaofeng Gong, Xiaolong Liang, Jiancheng Yang, Jun Jin, Kanglin Dai, Yuzhen Cui, Guan-Ming Kuang, Jianshen Xie, Libing Luo, Haibing Xiao, Shijie Yin, Jun Yang, Yulan Yan, Jianliang Chen, Yihua Chen, Qianshen Zhang, Qingshan Zhou, Lina Zhao, Min Wu, Xin Tang, Lei Rong, Zanxin Wang, Weifu Qiu, Yanli Wang, Liwen Cui, Xiangyang Li, Yong Hu, Huiren Tao, Nan Wu, Pearl Pai, Minxin Wei, Michael Kai-tsun To, Kenneth M.C. Cheung

## Abstract

**Background:** State-of-the-art (SOTA) large language models (LLMs) are poised to revolutionize clinical medicine by transforming diagnostic, therapeutic, and interdisciplinary reasoning. Despite their promising capabilities, rigorous benchmarking of these models is essential to address concerns about their clinical proficiency and safety, particularly in high-risk environments.

**Methods:** This study implemented a multi-disciplinary, randomized, single-blind evaluation framework involving 27 experienced specialty clinicians with an average of 25.9 years of practice. The assessment covered 685 simulated and real clinical cases across 13 subspecialties, including both common and rare conditions. Evaluators rated LLM responses on medical strength (0–10 scale, where *>*9.5 signified leading expert proficiency) and hallucination severity (0–5 scale for fabricated or misleading medical elements). Seven SOTA LLMs were tested, including top-ranked models from the ARENA leaderboard, with statistical analyses applied to adjust for confounders such as response length.

**Findings:** The evaluation revealed clinical plausibility in general-purpose LLMs, with Gemini 2.0 Flash leading raw scores and DeepSeek R1 excelling in adjusted analyses. Top models demonstrated proficiency comparable to a physician of 6 years post qualification experience (score ∼6.0), yet significant risks were noted. Instances of incompetence (scores ≤4) were detected across specialties, and 40 hallucination instances involving fabricated conditions, medications, and classification errors. These findings underscore the importance of implementing stringent safeguards to mitigate potential adverse outcomes in clinical applications.

**Interpretation:** While SOTA LLMs show substantial promise in enhancing clinical reasoning and decision-making, their unguarded application in medicine could present serious risks, such as misinformation and diagnostic errors. Human expert oversight remains crucial, particularly given reported incompetence and hallucination risks. Larger, multi-center studies are warranted to evaluate their real-world performance and track their evolution before broader clinical adoption.

## 1 Introduction

State-of-the-art large language models (SOTA LLMs) [1], such as ChatGPT, Claude, Grok, DeepSeek, and Qwen [2, 3], have demonstrated strong performance not only in language tasks but also in mathematics, programming, and domain-specific reasoning, including law, education, and medicine.

Although not specifically trained for such purposes, the values of general-purpose LLMs in medicine, such as diagnosis, genetics, treatment plan, and patient interaction [4, 5], were well noted since the early days of the current LLM-fueled A.I. boom. They also find applications in sub-specialties, including ophthalmology [6], clinical oncology [7], clinical genetics [8], mental health [9], neurology [10, 11], and orthopedics [12]. Additionally, LLMs were shown to have great values for rare diseases [13, 14]. Despite these benefits, LLMs face significant challenges that could impact their clinical utility, including limitations in factual accuracy [15], hallucinations [16], and cognitive and automation bias [17].

It is clear that only top-performing LLMs should be considered for medical applications, making rigorously benchmarking model “SOTAness” essential. Current strategies include supervised evaluations, crowdsourced preference-based ranking, and randomized comparative trials. Examples of the supervised approach include MMLU [18], MedQA [19], and MedMCQA [20], where both questions and answers were available for fast model evaluations. For example, board-exam-style clinical questions have been used to assess LLM performance in rheumatology [21]. The crowdsourcing platform ARENA [22] is a widely recognized leaderboard of LLM ranking. Randomized trials test the difference for using LLM and conventional tools (e.g. web-search or UpToDate) [23]. Each approach has limitations. Supervised benchmarks may promote overfitting, as test items are often open to model developers. Board-like questions lacks the complexity observed in real-case scenarios. Moreover, the reliance on multiple-choice formats precludes assessment of clinical reasoning, which is central to medical competence. Crowdsourced platforms typically lack evaluator qualifications, undermining scoring reliability. Additionally, testing time-window was also often not stringently defined. Randomized trials offer methodological rigor but are ill-suited for tracking rapidly evolving models over time. These paradigms also tend to focus on a few subjects and tend to evaluate the strength or plausibility, but neglect the risk assessment part.

To address these challenges, we developed a multi-disciplinary, single-blind evaluation framework in which only specialist clinicians, at the level of associate consultants or above, were invited to assess the clinical plausibility of responses generated by LLMs. Questions fell into two categories: structured board-exam-style scenarios and real-world patient cases drawn from local practice. To comprehensively reflect clinical complexity, 13 subspecialities, covering internal and surgical medicines, common and rare diseases, were included. To better reflect clinical reasoning, we replaced the conventional multiple-choice format with a rating on the model response texts. Each discipline provided a series of questions, and responses from seven SOTA LLMs were independently evaluated across two dimensions: medical strength and hallucination severity. Medical strength was scored on a 0–10 scale anchored to physician ranks within each specialty. Hallucinations—defined as fabricated or misleading medical content, including fictitious diseases, medications, guidelines, or literature—were scored from 0 to 5. Evaluations were conducted using a custom-built web interface that blinded model identities and randomized response order across questions. Statistical analyses, including multiple linear regression and linear mixed-effects modeling, were applied to quantify model performance while adjusting for confounders such as response length.

## 2 Methods

### 2.1 Study Design

The study was conducted in three rounds (Figure 1) at the University of Hong Kong-Shenzhen Hospital (HKU-SZH), a tertiary general hospital affiliated with the University of Hong Kong: round 1, multidisciplinary teams designed and categorized questions by difficulty and type; round 2, responses generated by LLMs were evaluated by clinical experts in a single-blind manner; round 3, models re-evaluated their own responses. Final results were analyzed using inferential statistics.

**Figure 1:**
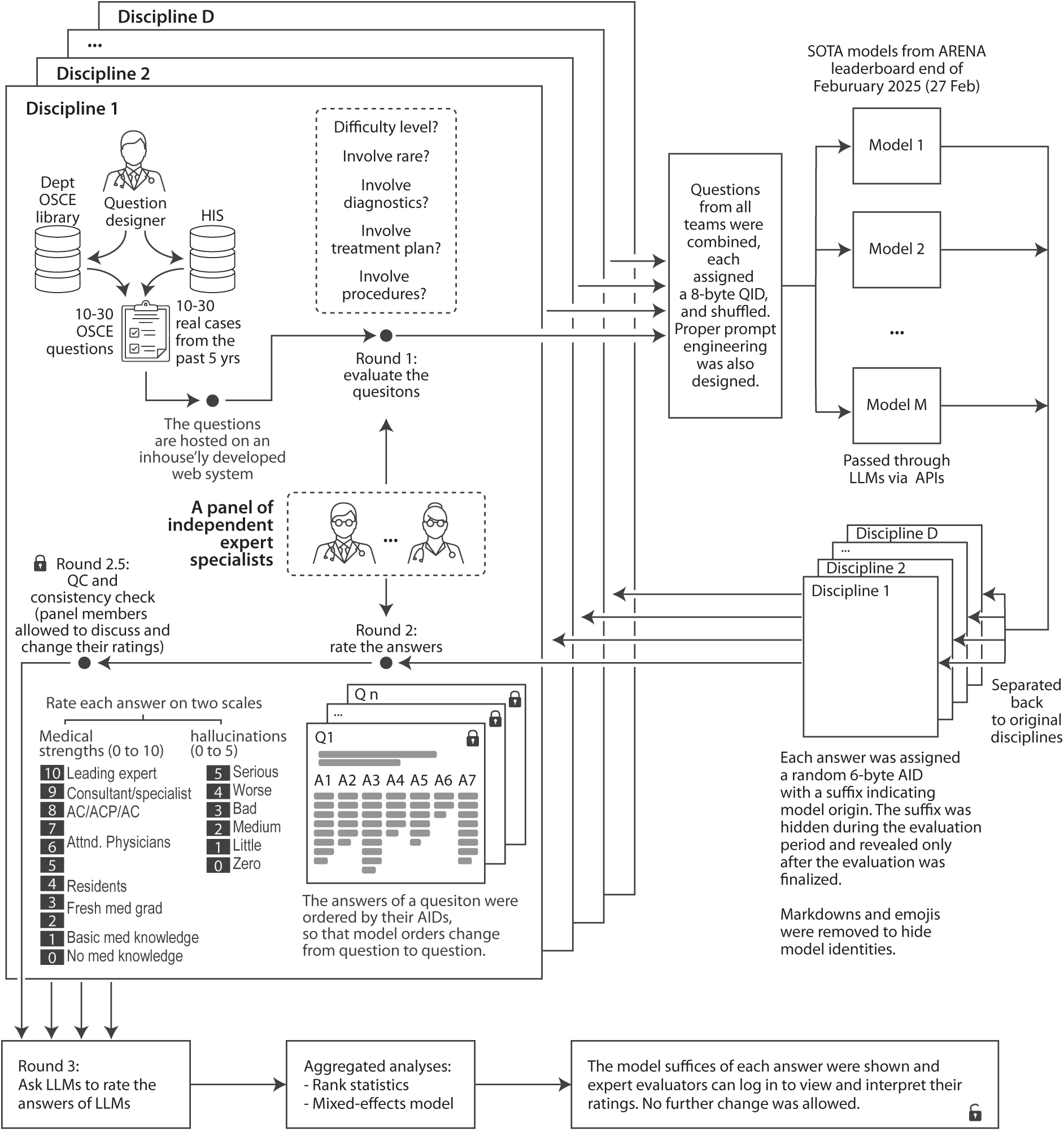
Overall study design. (AC/ACP/AS: associate consultant, associate chief physician, or associate specialist.)

### 2.2 Data collection and question designs

These disciplines were involved: accidents and emergency (A.E.), cardiology, cardiac surgery, pediatrics, neurology, nephrology, orthopedics, prenatal medicine, neonatal medicine (NICU), respiratory medicine, pediatric surgery, and rare diseases (supplementary Fig. S1A). Each discipline included one question designer (on average 14.8 years of medicine practices) and an independent evaluation panel of clinical experts.

Two types of questions were designed: (1) departmental OSCE (Objective Structured Clinical Examination) questions, (2) real scenarios cases. The OSCE is an established instrument of clinical competence [24] used for specialty board qualification examinations. The real cases were retrieved from the local hospital information system (HIS), from experienced clinicians to represent typical cases that are commonly encountered, as well as, unusually rare and difficult cases. Key clinical textual information including age, gender, history of presenting complaint, symptoms, physical examinations, past medical history, family history, drug history, laboratory tests, radiological reports, genetic tests, etc. forms key components of the question stems, in a text-only manner.

To balance statistical power and operational management, on average 24 to 25 questions per question-type per specialty were designed (supplementary Table S1A). All questions were manually checked to avoid unintended information leak. The final cutoff time for question submission was 23:59 on February 28, 2025. Detailed rationales for case selection and question designs by disciplines are provided in the supplementary Methods.

### 2.3 First round evaluation of the questions

All independent evaluators are experienced medically qualified specialists. They rated the questions in terms of level of difficulty (easy, medium, and hard), prevalence of the diseases (common or rare), and whether diagnostic, treatment, and operational reasoning are involved.

### 2.4 Prompt designs and API processing

In all questions, a standardized prompt of the type “You are a specialist in [discipline], please provide a professional diagnosis, treatment or surgical plan for the above case” was designed. No additional restrictions were imposed, such as, on the response lengths.

Questions of all disciplines were then aggregated and shuffled to avoid consecutive same-theme questions that may help the models learn, and were submitted to the models via APIs (application programming interface). Seven models were chosen for evaluations, including 4 of the top 10 models on ARENA on Feb 27, 2025 (supplementary Fig. S1B), and 3 lower-parameter models (deployed locally) of the same series. They were: Gemini 2.0 Flash Thinking exp-01-21 (Gemini 2.0), ChatGPT 4o latest 2025-01-29 (ChatGPT-4oL), DeepSeek-R1 671B (DeepSeek-R1), DeepSeek-R1 70B (DS70B), DeepSeek-R1 32B (DS32B), Qwen 2.5 Max (Qwen Max), and Qwq Latest (Qwen 32B) (supplementary Methods). Grok was not included due to unavailability of APIs at the time of the study. The experiments were run between March 8 and 9, 2025.

For the blinded evaluation by the independent expert panel, responses to each question were displayed side-by-side electronically (supplementary Fig. S1B). To prevent recognition bias, the responses were randomly reordered. Identifiable features such as emojis, excessive line breaks, and whitespace were removed to further mask model identity. Additionally, questions were also randomly ordered between evaluation rounds to mitigate fatigue-related bias.

### 2.5 Validity of blindness

To ensure blinding integrity, question-response and response-model mappings were encrypted and shared with evaluators. Decryption keys would be provided upon project completion to allow independent verification of blinding and confirmation of the absence of tampering.

### 2.6 Second round evaluation of the responses

The independent blinded experts evaluated each response on two aspects: (1) the quality (strength) of the answers (LLM responses) on a scale from 0 to 10 (step interval 0.5); (2) the degree of hallucinations on a scale from 0 to 5 (step interval 0.5). The strength is a measure of medical plausibility. We defined 0 as no knowledge, 0.5 or 1.5 as general knowledge, 2-3.5 as fresh medical graduates, 4-5.5 for residents (*<*3 years after graduation), 6-6.5 for senior residents, fellows, or physicians (3 6 years after graduation), 7-8 for associate consultants (associate specialist, 6-10 years after graduation), 8.5-9 for consultants (attending physician, *>*10 years after graduation), and ≥9.5 for leading experts (supplementary Methods). Hallucinations were also assessed on a scale from 0 to 5, with 1 given to any evidence of fabricated diseases, drugs, guidelines, consensus, and reference etc. A hallucination rating of 1 or above must be substantiated with solid evidence of fabrication, or it will be automatically adjusted to 0.5. This round took place from March 10 to March 24.

Upon completion of round 2, intra-discipline cross-evaluator consistencies were calculated and evaluators were alerted of potential inconsistency (*r <*0.3) where they may choose to adjust or keep their original ratings.

### 2.7 Non-independent and auxiliary evaluators

Four of the question designers who are expert specialists were also invited to participate in the evaluations. One senior physician was also invited to act as an auxiliary evaluator for pediatric surgery. Data from these two sets of evaluators were used for validations but otherwise excluded from drawing main conclusions.

### 2.8 Disqualification of evaluators and questions

One evaluator with low completion rate (*<*80%), and a second with excessive same-score ratings (*>*20%) were removed. A third evaluator withdrew voluntarily due to lack of time.

Ten of the original 695 questions were removed because of incomplete answers, unintended exposure of patient identity, or disclosure of diagnoses. Ultimately, a total of 685 questions were used for analysis.

### 2.9 Third round

The questions and model responses were submitted to the SOTA LLMs via APIs with request for evaluation of the quality of the responses, using the same metrics as the human experts.

### 2.10 Data processing

Per-question raw strength ascending ranks (PQRSA ranks) ranged from 1 to 7.0 (the number of models tested), and are calculated by ranking the raw scores given by one evaluator to answers of a question in ascending order, such that higher ranks correspond to higher raw scores. In cases of ties, the average method was used. PQRSA ranks were treated as integers in the regression analyses.

### 2.11 Inferential statistics

Multiple linear regression, linear mixed-effects (LME) model, and logistic regression were performed to estimate the influence of models, response-length, and question characteristics on the scoring outcomes (supplementary Methods).

## 3 Results

### 3.1 Multi-disciplinary expert evaluation of SOTA models

In all, 30 independent from 13 specialties took part in the study (supplementary Table S1). One evaluator withdrew and two were removed according to our criteria for quality control (Methods, supplementary Fig. S2a, S3, and S4). The 27 qualified evaluators averaged 25.9 years of clinical experience (supplementary Fig. S1A). After excluding 10 questions due to incomplete answers or information leaks (see Methods; Supplementary Table S1A), 685 questions (48.9% recent clinical cases, 51.1% OSCE) were processed. The A.E. team provided two sets of real-case questions, whereas the rare disease team had no OSCE question-set.

Real-world case questions were rated as more difficult than OSCE items, with 31% versus 9% considered ‘hard’, respectively (Supplementary Fig. S2A). In six specialties (cardiology, ICU, respiratory medicine, prenatal diagnosis, pediatrics, and cardiac surgery), evaluators rated over half (*>*50%) of the real-world case questions as ‘hard’. Overall, 18.2% (125/685) of questions pertained to rare diseases. Such cases were concentrated within specific specialties, constituting 100%, 97.7%, and 36.8% in prenatal diagnosis, rare disease category, and neurology, respectively. Knowledge of other question characteristics including involvement of diagnosis, treatment, and operational procedures help evaluate the models’ reasoning capacity and spatial understanding (Supplementary Fig. S2D-F).

### 3.2 Gemini 2.0 had the leading performance in unadjusted medical strength scores

In the second round, we obtained 11,995 scores (SetABC) on medical strengths, of which 9,856 (SetA), 1,824 (SetB), and 315 (SetC) have been obtained on 4,795 questions by 27 independent, 4 non-independent, and 1 auxiliary evaluators, respectively, or 318 (median) scores per evaluator (inter-quartile range [IQR], 296 to 364). The median raw strengths for SetA and SetABC are both 6.5 (and IQR both 5.5 to 7.5), equivalent to the capacities of an experienced physician with 6 years of practices. Upon breaking down into different models, Gemini 2.0 Flash topped the leaderboard with an average score of 7.1 (standard error [s.e.] 0.04), followed by Qwen Max (mean 6.9), DS-r1 671B (mean 6.8), ChatGPT-4o (6.7), Qwq latest (Qwen 32B, at 6.6). DS32B and DS70B were considerably lower, at 5.9 and 5.7, respectively (supplementary Fig. S6A). *Post hoc* Tukey analysis revealed multiple significant pairwise differences in mean strength scores across models, with Gemini, Qwen Max, and DS-r1 671B outperforming ChatGPT-4o, while DS32B and DS70B showed significantly lower performance than most other models (adjusted *p <*0.001 for the majority of comparisons). Discipline-wise, Gemini was the winner in 10 out of all 14 specialties, followed by Qwen max (winner in 01b.AE and 09.Prenatal) and DS-r1 (winner in 01a.AE and 04.Neural) (supplementary Fig. S6).

Gemini outperformed other models in 9 of the 14 real-case question-sets, followed by DeepSeek R1 (01a.AE, 10.Neonatal, and 12.PediSurg), and Qwen Max (01b.AE and 09.Prenatal) (Figure 2B). Gemini also outperformed other models in 8 of the 12 OSCE question-sets, followed by ChatGPT-4o (01a.AE and 09.Prenatal), Qwen Max (06.Ortho), and DeepSeek R1 (04.Neural) (Figure 2B).

**Figure 2:**
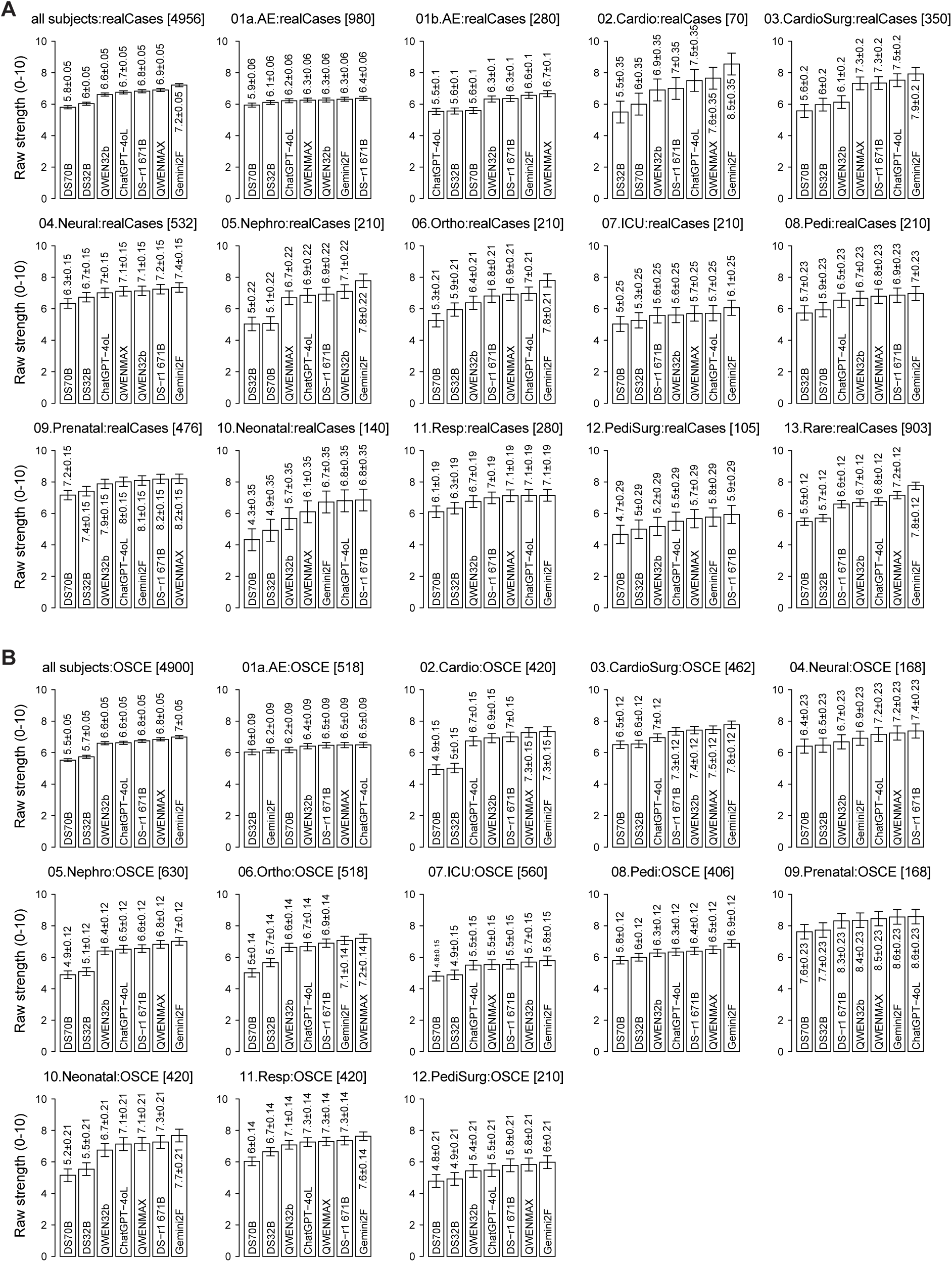
Raw scores on strengths by the independent evaluators (OSCE and real-case questions separated). (A) Real-case questions. (B) OSCE questions

### 3.3 DeepSeek R1 outperformed other models after adjusting for response length

We noted that the models produced highly variable lengths of responses with regard to our questions. For example, on average, Gemini output 3,948 characters per question, over two times longer than the second model Qwen Max (1,829 characters). Qwen 32B, ChatGPT-4o, and DeepSeek R1 output 1,638, 1,429 and 1,303 characters, respectively (supplementary Fig. S6A). Despite ranking fifth in response lengths, DeepSeek R1 achieved third in overall raw strength scores, with no statistical difference (Tukey’s honesty test *p*=0.63) from the second model (Qwen Max). Interestingly, the response lengths also differ by question types, with longer outputs (350 characters) for real-case questions (supplementary Table S2). We tested whether longer output conferred advantages to receiving higher scores. Linear regression model suggests that both raw strength scores and per-question raw strength ranks were significantly dependent on the response lengths (supplementary Table S3). This advantage is not linear, with real-valued response-length only explaining 2.4% and 4.7% of the raw scores and raw ranks, respectively (supplementary Table S3A). Instead, formulating the response lengths as a categorical variable explains most variance, with 11.5% and 24.7% for the raw scores and raw ranks, respectively (supplementary Table S3C). This is consistent with the observation that although Gemini responses were three times longer than other models’, it does not received proportionately more advantages (supplementary Fig. S8A-B). For this reason, we introduced the categorical rank of the response lengths as a co-variate in future models, and not the real-valued or integer lengths.

We first performed a multiple linear model (Model 1), with question type, model, subject, and other characteristics of the questions (difficulty, involvement of rare diseases, diagnosis, treatment plan, and procedures) as covariates, but not the response lengths, which explained 96.7% and 96.4% of the variances in SetA (Model 1a) and SetABC (Model 1b), respectively (supplementary Table S4). For comparison, we introduced a second model (Model 2), which in addition to all covariates of Model 1, also included the response length (in its categorical form). Model 2a and Model 2b explained highly similar variances as their Model 1 counterparts, at 96.8% and 96.4%, respectively (supplementary Table S5A). However, the two models are significantly different (F-test *p <*0.001), suggesting the necessity to include response length as a covariate (supplementary Table S5B). It appears that with the col-linearity between model and response-length, the variance structure was shifted in Model 2, under which DeepSeek R1 was now the strongest model (*β̂* = 6.06), followed by Qwen Max (*β̂* = 5.96), Gemini (*β̂* = 5.94), and ChatGPT-4o (*β̂* = 5.93) (supplementary Table S5). Qwen32B, DS32B, and DS70B were considerably lower, at *β̂* = 5.78, 5.52, and 5.29 respectively. We noted that the response-length conferred 0.17, 0.41, 0.53, 0.61, 0.67, and 0.92 of raw strength scores to Model 2a for each within-question ascending rank of 2 to 7, respectively (supplementary Table S5). The disciplines significantly influenced how the answers were scored, which could be due to both evaluator-biases and intrinsic differences among disciplines.

Instead of using raw scores, we ranked the per-question (and per-evaluator) scores in ascending order, and used the ranks, which would then be independent of evaluator biases, to compare the models. Model 3 and Model 4 had the same sets of covariates as Models 1 and 2, respectively, but with ascending score ranks as independent variable (Table 1, supplementary Table S6A). As expected, the disciplines were now insignificant in both Models 3 and 4, as this ranking has nothing to do with evaluator, subject, or question characteristics. It is only affected by their relative performance within each question. Again, ANOVA results (supplementary Table S7B) were in favor of the more complex model (Model 3a) with response-length as a covariate, under which DeepSeek R1 was also the strongest model (*β̂*_rank_ = 3.49), followed by Qwen Max and Gemini, with both *β̂*_rank_ = 3.35, respectively (Table 1).

**Table 1:**
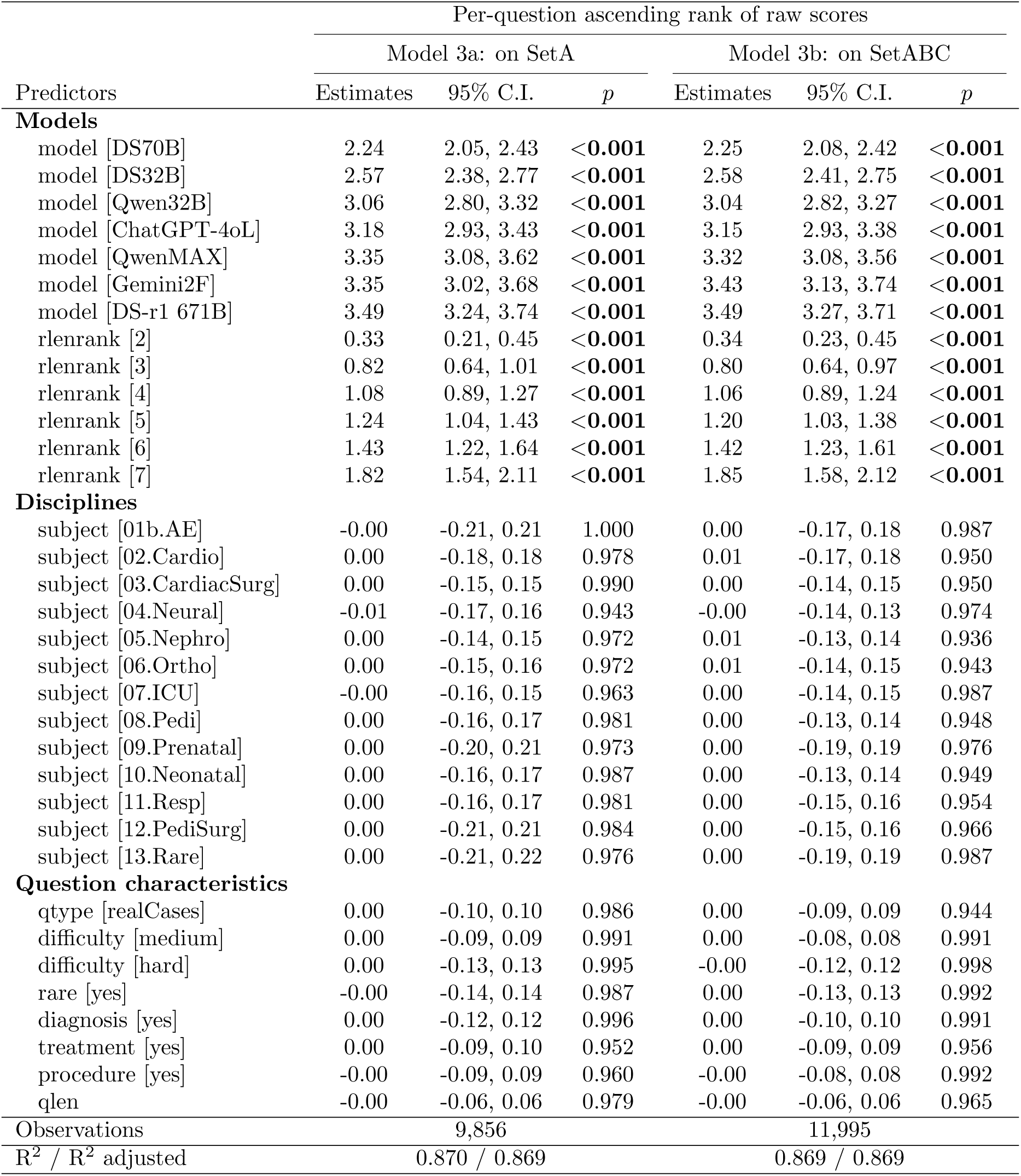
Multiple linear regression results for evaluator-assigned per-question rank scores (1–7; higher = better) across all questions, evaluators, and subjects. DS32B is the reference model. rlenrank denotes model response length ranks (1–7; higher = longer), with a full-table on estimates of ties in supplementary Table S6. The reference levels are: subject (01a.AE), question difficulty (easy), and absence of rare disease, diagnosis, treatment, and procedure content (no for each). Estimates are shown with 95% confidence intervals and p-values.

### 3.4 Linear mixed-effects model revealed SOTA models have reached strength of experienced physicians

Rank-based statistics allow for model comparison but sacrifice precision in intrinsic scores, whereas direct linear modeling of strength on covariates is affected by high inter-rater variability. To overcome this, we fitted a linear mixed-effects (LME) model, where question- and evaluator-specific variations were modeled as random effects, while model, question-type, response-length, subject, and question-specific characteristics were treated as fix-effect variables (Methods). The results are presented in Table 2. Strength was found to be significantly dependent on model and response-length. DS-r1 671B demonstrated the strongest positive estimate (*β̂* = 6.05; 95% CI, 5.07 to 7.03), closely followed by Qwen Max (*β̂* = 5.93; 95% CI, 4.97 to 6.93), Gemini2F (*β̂* = 5.93; 95% C.I., 4.94 to 6.91), and ChatGPT-4oL (*β̂* = 5.91; 95% C.I., 4.94 to 6.89). Conversely, models DS32B and DS70B underperformed with *β̂* = 5.51 and 5.28, respectively. These results are consistent with the rank-based analysis in Table 1.

**Table 2:**
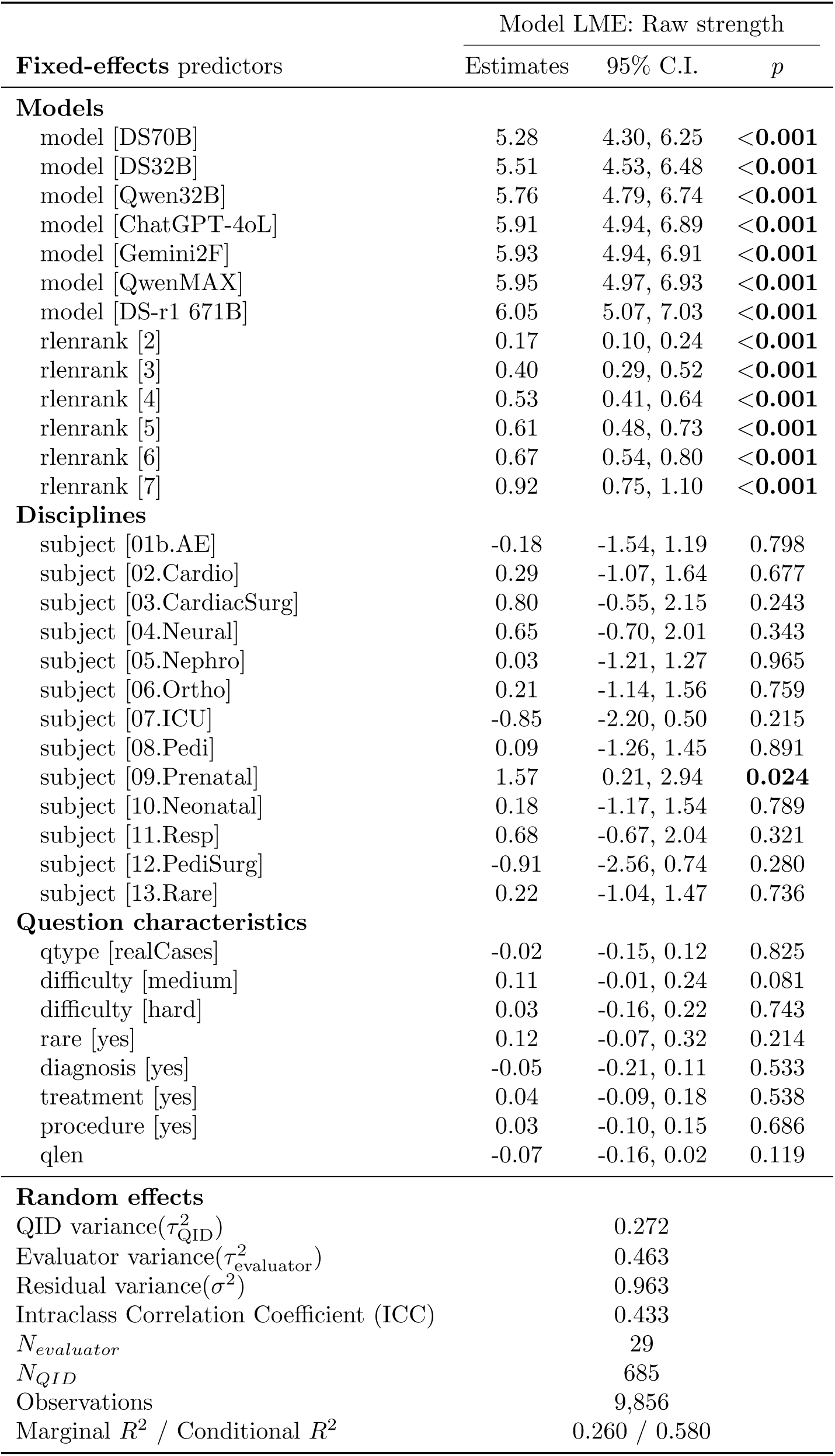
Linear mixed-effects modeling of raw strength scores. References: rlenrank [1.0], qtype: OSCE, subject: 01a.AE, difficulty: easy, rare: no, diagnosis: no, treatment: no, procedure: no. Ties of rlenrank were shown in supplementary Table S7.

| Fixed-effects predictors | Model LME: Raw strength |  |  |
| --- | --- | --- | --- |
|  | Estimates | 95% C.I. | *p* |
| **Models** |  |  |  |
| model [DS70B] | 5.28 | 4.30, 6.25 | <0.001 |
| model [DS32B] | 5.51 | 4.53, 6.48 | <0.001 |
| model [Qwen32B] | 5.76 | 4.79, 6.74 | <0.001 |
| model [ChatGPT-4oL] | 5.91 | 4.94, 6.89 | <0.001 |
| model [Gemini2F] | 5.93 | 4.94, 6.91 | <0.001 |
| model [QwenMAX] | 5.95 | 4.97, 6.93 | <0.001 |
| model [DS-r1 671B] | 6.05 | 5.07, 7.03 | <0.001 |
| rlenrank [2] | 0.17 | 0.10, 0.24 | <0.001 |
| rlenrank [3] | 0.40 | 0.29, 0.52 | <0.001 |
| rlenrank [4] | 0.53 | 0.41, 0.64 | <0.001 |
| rlenrank [5] | 0.61 | 0.48, 0.73 | <0.001 |
| rlenrank [6] | 0.67 | 0.54, 0.80 | <0.001 |
| rlenrank [7] | 0.92 | 0.75, 1.10 | <0.001 |
| **Disciplines** |  |  |  |
| subject [01b.AE] | -0.18 | -1.54, 1.19 | 0.798 |
| subject [02.Cardio] | 0.29 | -1.07, 1.64 | 0.677 |
| subject [03.CardiacSurg] | 0.80 | -0.55, 2.15 | 0.243 |
| subject [04.Neural] | 0.65 | -0.70, 2.01 | 0.343 |
| subject [05.Nephro] | 0.03 | -1.21, 1.27 | 0.965 |
| subject [06.Ortho] | 0.21 | -1.14, 1.56 | 0.759 |
| subject [07.ICU] | -0.85 | -2.20, 0.50 | 0.215 |
| subject [08.Pedi] | 0.09 | -1.26, 1.45 | 0.891 |
| subject [09.Prenatal] | 1.57 | 0.21, 2.94 | **0.024** |
| subject [10.Neonatal] | 0.18 | -1.17, 1.54 | 0.789 |
| subject [11.Resp] | 0.68 | -0.67, 2.04 | 0.321 |
| subject [12.PediSurg] | -0.91 | -2.56, 0.74 | 0.280 |
| subject [13.Rare] | 0.22 | -1.04, 1.47 | 0.736 |
| **Question characteristics** |  |  |  |
| qtype [realCases] | -0.02 | -0.15, 0.12 | 0.825 |
| difficulty [medium] | 0.11 | -0.01, 0.24 | 0.081 |
| difficulty [hard] | 0.03 | -0.16, 0.22 | 0.743 |
| rare [yes] | 0.12 | -0.07, 0.32 | 0.214 |
| diagnosis [yes] | -0.05 | -0.21, 0.11 | 0.533 |
| treatment [yes] | 0.04 | -0.09, 0.18 | 0.538 |
| procedure [yes] | 0.03 | -0.10, 0.15 | 0.686 |
| qlen | -0.07 | -0.16, 0.02 | 0.119 |
| **Random effects** |  |  |  |
| QID variance($\tau_{QID}^2$) | | 0.272 | |
| Evaluator variance($\tau_{evaluator}^2$) | | 0.463 | |
| Residual variance($\sigma^2$) | | 0.963 | |
| Intraclass Correlation Coefficient (ICC) |  | 0.433 |  |
| $N_{evaluator}$ | | 29 | |
| $N_{QID}$ | | 685 | |
| Observations |  | 9,856 |  |
| Marginal $R^2$ / Conditional $R^2$ | | 0.260 / 0.580 | |

Response-length (rlenrank) was the major predictor apart from models, with strongest positive effects observed at levels 7 (*β̂* = 0.92) and 6 (*β̂* = 0.67). Virtually no other predictor had any significant influence on the strength scores except a spike in Prenatal diagnosis (*β̂* = 1.57; 95% CI, 0.21 to 2.94), while question-type and all other question characteristic variables are non-significant. Random-effects analysis indicated substantial variability across evaluators and question identifiers (QIDs), reflected by ICC values (0.43) and variance components 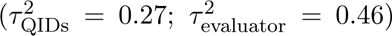. As compared with the linear model (Model 3a), the final LME model explained a moderate portion of variance (marginal *R*^2^ = 0.260), which notably increased when accounting for random effects (conditional *R*^2^ = 0.580), underscoring the importance of individual-level variability among the evaluators and questions. Much of the subject-specific effects in Model 3a cannot be delineated with those of the evaluators in the LME model, and thus their actual effects are inconclusive in the current study. In all, after adjusting for response-length and modeling the random effects, SOTA models including DeepSeek r1, Qwen Max, Gemini 2F, and ChatGPT-4o have reached the strength of ∼ 6.0, equivalent to that of a physician with 3-6 years of practices on our scale.

### 3.5 Predictors for excellence

To assess model performance in identifying high-quality answers, we defined excellence to be score ≥ 8, where 1,862 instances were observed (18.9%), and applied a generalized linear mixed-effects model with evaluator effects as random intercepts. The fitted model showed strong explanatory power (marginal *R*^2^ = 0.48; conditional *R*^2^ = 0.66), with evaluator effects accounting for 35% of variance (Table 3). Compared with the reference model (DS32B), DS-r1 671B (odds ratio [OR] 2.88; 95% CI 1.90 to 4.38), Qwen Max (OR 2.48), ChatGPT-4oL (OR 2.44), and

**Table 3:**
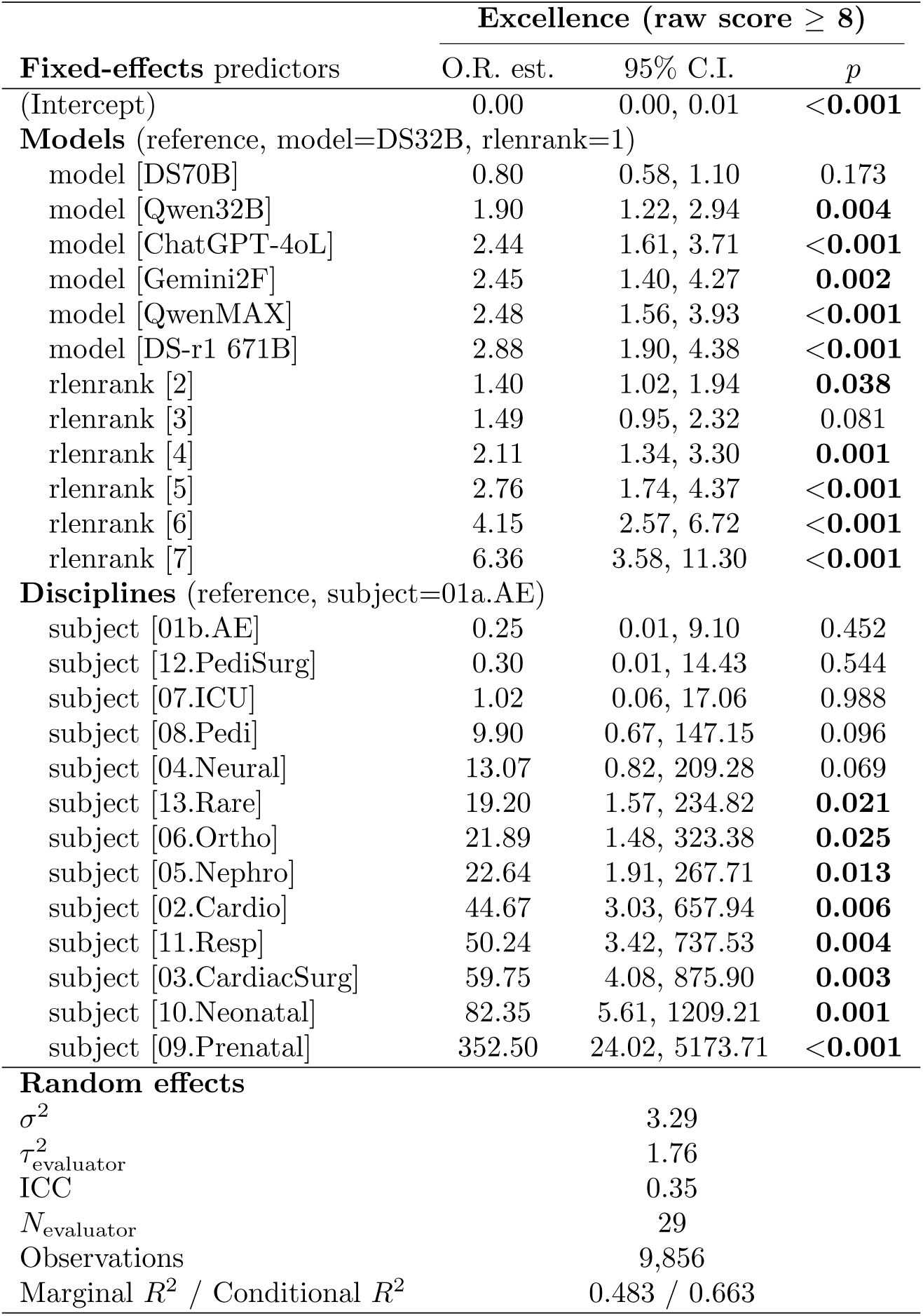
Fixed effects estimates and odds ratios with 95% confidence intervals from logistic mixed model predicting excellence (score ≥ 8), adjusted for evaluator random effects

Gemini2F (OR 2.45) had significantly higher odds of excellence (*p <* 0.01 for all). Increasing response length rank was associated with a stepwise increase in excellence probability. Subspecialty was also a significant predictor for excellence. Prenatal diagnosis, neonatology, cardiac surgery, and respiratory medicine were ranked higher in value, while intensive care, neurology, pediatric surgery and Accident and Emergency (set 2) showed no significant difference from the reference.

### 3.6 Risks for incompetency

We asked the chances of incompetency, defined as receiving a score of 4 or below, across models and subspecialties. In all, 714 such instances were observed (7.2%), with Gemini 2.0 (2.7%) having the lowest ratio of incompetence (Table 4). Interestingly, despite ranking only fourth highest in excellence (19.9%), DS-r1 ranked second lowest (3.4%) in incompetence. We fitted a logistic regression with discipline and model identity as fixed effects, and evaluator identity as a random intercept to adjust for inter-rater scoring variability. Compared with the reference (DS32B), Gemini2F, DS-r1 671B, Qwen32B, Qwen Max, and ChatGPT-4oL had significantly lower odds ratio of incompetence, at 0.16, 0.21, 0.23, and 0.25, respectively (supplementary Tables S12 and S13), while DS70B has a significantly elevated failure rate (OR 1.95, *p <* 0.001). Across subspecialties, intensive care, neonatology, and pediatric surgery exhibited the highest adjusted failure risks (all *p <* 0.01), followed by cardiology, neurology, nephrology, orthopedics, and rare diseases, as compared with the reference (A.E.). Respiratory, pediatrics, and prenatal medicine showed no difference from the reference.

**Table 4:**
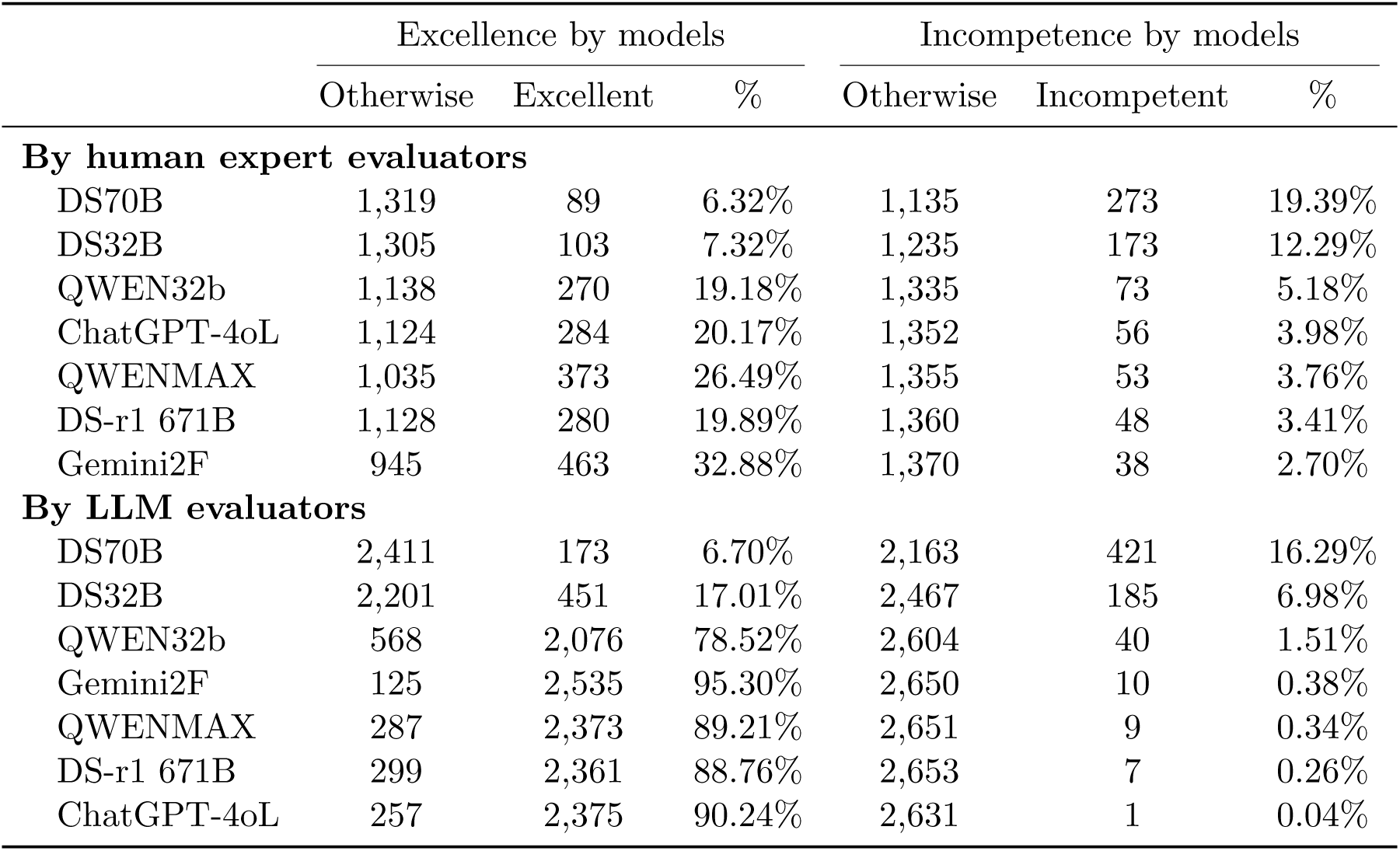
Raw excellence and incompetence ratios by models

### 3.7 Hallucination analyses in model responses

Another form of risks is hallucinations [25], which is defined as fabricated medical facts. Forty hallucinations were reported (supplementary Excel), which include fabricated drugs and dosages (n=5), conditions (n=2), subtyping (n=3), physical examination details in OSCE items (n=7), mis-interpretation of variant pathogenicity due citing wrong consensus (n=15). Such events were reported in pediatrics, nephrology, neurology, prenatal diagnosis, respiratory medicine, and orthopedics.

In prenatal diagnosis, 17 instances of hallucinations, all exclusively in real-case items, predominantly occurred during variant pathogenicity interpretation for rare genetic diseases. The most common errors involved misinterpretation or incorrect application of variant classification criteria such as PVS1, PS3/4, PM1/2/4, and PP1/3. For example, responses to Questions 3, 7 and 23 by Qwen 32B, Q8 by ChatGPT-4o, and Q20 by Gemini, misused the ACMG-AMP criteria [26, 27] and the ClinGen Sequence Variant Interpretation framework. In Q11 of the same subject, five of all seven models (except Qwen Max and DeepSeek r1) misinterpretated functions of *DHTKD1*, a relatively understudied gene with limited and conflicting literature. Additional examples include mis-attribution a kidney-specific phenotype (Q9) to mitochondrial disease by Qwen 32B, and a misdiagnosis of a case (Q19, by DS70B) as spinocerebellar ataxia and falsely equated with Joubert syndrome, a clearly fabricated association. In pediatrics, 12 events (9 in OSCE-and 2 in real-case items) were detected, with alarming ignorance of pediatric age restrictions or dosing. In Q5, Qwen32B recommended doxycycline at nearly twice the evidence-based dose specified in national guidelines. ChatGPT-4o incorrectly endorsed levofloxacin use in children under 18, contradicting national antimicrobial prescribing principles. DeepSeek models (70B and 32B) fabricated family and exposure histories (Q4), while Qwen 32B (Q9) invented physical exam findings not included in the case. In real-case pediatric questions, hallucinations appeared in diagnostic reasoning. For example, in Q25, Qwen32B diagnosed NK cell deficiency based solely on a single low count, ignoring contextually normal immunoglobulins and a known history of coconut oil aspiration. In contrast, DeepSeek R1 671B identified the same laboratory finding but interpreted it conservatively, bringing a high-score. In Q38, ChatGPT-4o cited incorrect reference ranges for G6PD, leading to a false diagnosis of G6PD deficiency. In respiratory medicine (Q11), the suggestion that osimertinib-related ILD requires drug discontinuation oversimplified clinical decision-making and contradicts with current gold standard [28, 29]. Instances of fabricated literature were reported in Orthopedics, and fabricated drugs in nephrology and neurology, where non-existing wrong drug names were suggested (supplementary Excel).

### 3.8 Asking LLMs to rate LLM responses

We asked how the LLM would rate responses of their own and by other LLMs, in the same scale as our human evaluators. Only the four SOTA models were able to process this request, whereby ratings on 4,623 on 675 questions were obtained. The LLMs appeared to have high mutual consensus, with pairwise correlations ranging from 0.71 to 0.79 (supplementary Table S13). Performing a multiple linear regression on the same set of variables as Model 2 but with LLM evaluator as an additional covariate, we found that LLMs were indeed more generous than human evaluators, with their agreed best model ChatGPT-4o rated 7.2 on average. Both human experts and LLMs agreed that DS32B and DS70B are the worst performers, and that DS70B underperformed DS32B (Table 4, supplementary Table S14). The LLMs also appeared to favor longer responses too. As compared with A.E., responses in neurology, nephrology, pediatrics, and respiratory medicine were significantly more approved, while other subjects showed no difference. Increasing question difficulties did significantly decrease the scores, while question type and rare disease were not. Overall, these results were rather consistent how human experts evaluated the models.

## 4 Discussion

SOTA LLMs are game-changers in medicine, and their safe use are paramount. The study presents a comprehensive multi-disciplinary expert-evaluation of SOTA LLMs in clinical application, using OSCE and real-case questions as lead questions. There are several important findings.

First, as compared with recent studies [33, 34], our rigorous methodological framework employ unique designs, including real-case, physician-experience based metrics, and human expert evaluation. We argue that linking LLM plausibility to physician experiences, which has the potential to further perform cross-sectional and longitudinal tracking on a larger scale. Expert evaluation and QC measures ensures the quality of the scores. As opposed to multiple-choice testing, content-based evaluation also allows for scrutinizing reasoning competence and clinical practicality, reducing overfitting and potential fraud. The study was also conducted in a short time-window on latest models, reducing unfairness caused by the time factor.

Second, from the statistical analyses of over 11,000 data-points by 27 qualified experts on 685 questions, we found that Gemini 2.0 Flash topped the leaderboard, but with over three times longer responses. On a rank-based or on linear mixed-effects model and adjusting for response-length, DeepSeek r1 was the lead, indicating its robustness in medical reasoning despite shorter responses. The LME model further revealed that SOTA models including DeepSeek r1 and Gemini 2.0 Flash have achieved the level of competency equivalent to a physician with 3-6 years of post-qualification experience. This likely underestimates, rather than overestimates the models’ capabilities, as evaluators have no reasons to inflate the performance of technologies that could threaten their professional roles, which is consistent with the LLMs-rated results.

Third, while SOTA models demonstrate impressive capabilities across various medical disciplines, with no significant interdisciplinary differences in their performances. Significant limitations restrict their clinical applicability. Notably, a concerning 7.2% rate of diagnostic incompetence and 40 instances of fabricated medical facts, including erroneous diagnoses, drug recommendations, and genetic interpretations, were observed. For example, in a simulated A.E. case with typical inferior ST-segment elevation myocardial infarction (STEMI) complicated by right ventricular infarction and hypotension (90/50 mmHg), most models offered routine reminders to monitor blood pressure when using nitroglycerin, while DeepSeek r1 demonstrated a more profound understanding of the pathophysiology, by integrating the patient’s low blood pressure and signs of right heart failure such as jugular venous distension, to infer the right ventricular involvement and explicitly recommend avoiding nitroglycerin uses, which fully aligns with the American College of Cardiology guidelines [30]. Fabrications pertaining to variant site interpretation in prenatal diagnosis were detected, consistent with the known limitations of LLM’s use in clinical genetics [31, 32]. Such inaccuracies underline the inherent risks of relying on these models for patient management. Based on these findings, it is clear that LLM models cannot replace human physicians in clinical decision-making. Both medical professionals and the public must remain cognizant of these limitations, ensuring that the deployment of these technologies in healthcare settings is carefully evaluated and rigorously supervised.

There are some drawbacks in our study. First, despite its closer connection to current medical practices, human expert evaluation is subjective. But this approach is safer to outcome-based designs such as clinical trials on real patients. Second, model response lengths appeared to be a major confounder of performance scores, which can be constrained via prompt engineering. Future work will involve more disciplines, models, and multi-center experts. Inherent human factors (fatigue, bias) can be mitigated with larger studies.

## 5 Conclusion

The rigorous methodology implemented in this study facilitated a comprehensive multi-disciplinary, randomized, and blinded expert evaluation of SOTA LLMs using structured scenarios and real-world hospital cases. This approach provides a robust framework applicable for future assessments of similar technologies.

While LLMs demonstrated remarkable capabilities in clinical reasoning, their inherent limitations, including diagnostic inaccuracies and hallucinations, underscore their inability to fully replace human clinicians in patient care. These findings highlight the importance of cautious integration of LLMs into medical practice, emphasizing that their use should complement, not substitute, the expertise of medical professionals. The advancement of prompt engineering and targeted training may mitigate certain weaknesses, enabling safer and more effective deployment in specific healthcare settings.

## Ethical approval

Ethical approval for this study was obtained from the institutional review board with ID Ethics-2025-078. Informed consent was waived. Questions concerning real patients were checked for any personal identifiers, which were then removed, before submitting to models.

## Contributions

KMCC and PKC conceived the project. KMCC and MKTT coordinated the efforts. PKC designed the methodology, developed and implemented the web system, designed and carried out the analyses, and wrote the first draft. JFC, JYZ, SXC, and CGX are question designers and non-independent expert evaluators for the neurology, pediatrics, A.E., and NICU teams. LHY is the auxiliary evaluator for pediatric surgery. LHY, XYD, XWC, YZW, XL, SFG, XLL, and JCY are question designers for their affiliated departments. The co-first authors were all involved in designing methodology. JJ, KLD, YZC, GMK, JSX, LBL, HBX, SJY, JY, YLY, JLC, YHC, QSZhang, QSZhou, LNZ, MW, XT, LR, ZXW, WFQ, YLW, LWC, XYL, MXW, HRT, NW, and PP are the independent evaluators in their respective affiliations’ disciplines (see also supplementary Fig. S1). All evaluators vouched for the professionalism and neutrality in the evaluation process. Co-first authors co-wrote the first draft, and all authors contributed to, revised and approved the final draft.

## Data Availability

The code for processing the LLM responses, running the web-system, and for analyzing the resulting data were deposited on an open repository (\url{https://github.com/HKUSZH/LLMMed}). The JSON files for the model responses, evaluation scores, and sample questions, were also uploaded to the same link. Complete question-sets are available upon request to the corresponding authors.

https://github.com/HKUSZH/LLMMed

## Acknowledgments

We thank the support of the Sanming Project of Medicine in Shenzhen (SZSM202211004 and SZSM202311022), Shenzhen Clinical Research Center for Rare Diseases (LCYSSQ20220823091402005), Shenzhen Key Medical Discipline Construction Fund (SZXK2020084 and SZXK077), and the Shenzhen Science and Technology Major Project of China (KJZD20240903102759061). We thanked the three disqualified evaluators, X, Y, and Z, for their engagement. Certain parts of their ratings are still valuable though not included the final analyses. We understand their heavy clinical duties may have a role in their “disqualification”. We thank Mr Zhong Cheng for helping with some API processing, Mr Dong Chen for coordinating with the orthopedic team, and Prof Pak Chung Sham (HKU), Drs Abraham Wai and Chongfei Jiang, Prof Nai Ding (ZJU), and Mr Kevin Tam for useful discussions. PKC thanks the support of Shenzhen Peacock Plan (No.20210830100C) and Futian Talent Program.

## 5.1 Software and data availability

The code for processing the LLM responses, running the web-system, and for analyzing the resulting data were deposited on an open repository (https://github.com/HKUSZH/LLMMed). The JSON files for the model responses, evaluation scores, and sample questions, were also uploaded to the same link. Complete question-sets are available upon request to the corresponding authors.

## References

[1] Vaswani, Ashish, Noam Shazeer, Niki Parmar, Jakob Uszkoreit, Llion Jones, Aidan N. Gomez, Lukasz Kaiser, and Illia Polosukhin. “Attention is all you need.” Advances in neural information processing systems 30 (2017).

[2] Sapkota, Ranjan, Shaina Raza, and Manoj Karkee. “Comprehensive analysis of transparency and accessibility of chatgpt, deepseek, and other sota large language models.” arXiv preprint arXiv:2502.18505 (2025).

[3] Guo, Daya, Dejian Yang, Haowei Zhang, Junxiao Song, Ruoyu Zhang, Runxin Xu, Qihao Zhu, et al. “Deepseek-r1: Incentivizing reasoning capability in llms via reinforcement learning.” arXiv preprint arXiv:2501.12948 (2025).

[4] Singhal, Karan, Shekoofeh Azizi, Tao Tu, S. Sara Mahdavi, Jason Wei, Hyung Won Chung, Nathan Scales et al. “Large language models encode clinical knowledge.” Nature 620, no. 7972 (2023): 172–180.

[5] Thirunavukarasu, Arun James, Darren Shu Jeng Ting, Kabilan Elangovan, Laura Gutierrez, Ting Fang Tan, and Daniel Shu Wei Ting. “Large language models in medicine.” Nature medicine 29, no. 8 (2023): 1930–1940.

[6] Lim, Zhi Wei, Krithi Pushpanathan, Samantha Min Er Yew, Yien Lai, Chen-Hsin Sun, Janice Sing Harn Lam, David Ziyou Chen et al. “Benchmarking large language models’ performances for myopia care: a comparative analysis of ChatGPT-3.5, ChatGPT-4.0, and Google Bard.” EBioMedicine 95 (2023).

[7] Rydzewski, Nicholas R., Deepak Dinakaran, Shuang G. Zhao, Eytan Ruppin, Baris Turkbey, Deborah E. Citrin, and Krishnan R. Patel. “Comparative evaluation of LLMs in clinical oncology.” Nejm Ai 1, no. 5 (2024): AIoa2300151.

[8] McGrath, Scott P., Beth A. Kozel, Sara Gracefo, Nykole Sutherland, Christopher J. Danford, and Nephi Walton. “A comparative evaluation of ChatGPT 3.5 and ChatGPT 4 in responses to selected genetics questions.” Journal of the American Medical Informatics Association 31, no. 10 (2024): 2271–2283.

[9] Heinz, Michael V., Daniel M. Mackin, Brianna M. Trudeau, Sukanya Bhattacharya, Yinzhou Wang, Haley A. Banta, Abi D. Jewett, Abigail J. Salzhauer, Tess Z. Griffin, and Nicholas C. Jacobson. “Randomized Trial of a Generative AI Chatbot for Mental Health Treatment.” NEJM AI 2, no. 4 (2025): AIoa2400802.

[10] Du, Xinsong, John Novoa-Laurentiev, Joseph M. Plasek, Ya-Wen Chuang, Liqin Wang, Gad A. Marshall, Stephanie K. Mueller, et al. “Enhancing early detection of cognitive decline in the elderly: a comparative study utilizing large language models in clinical notes.” EBioMedicine 109 (2024).

[11] Moura, Lidia, David T. Jones, Irfan S. Sheikh, Shawn Murphy, Michael Kalfin, Benjamin R. Kummer, Allison L. Weathers et al. “Implications of large language models for quality and efficiency of neurologic care: emerging issues in neurology.” Neurology 102, no. 11 (2024): e209497.

[12] Sosa, Branden R., Michelle Cung, Vincentius J. Suhardi, Kyle Morse, Andrew Thomson, He S. Yang, Sravisht Iyer, and Matthew B. Greenblatt. “Capacity for large language model chatbots to aid in orthopedic management, research, and patient queries.” Journal of Orthopaedic Research@ 42, no. 6 (2024): 1276–1282.

[13] Mehnen, Lars, Stefanie Gruarin, Mina Vasileva, and Bernhard Knapp. “ChatGPT as a medical doctor? A diagnostic accuracy study on common and rare diseases.” MedRxiv (2023): 2023–04.

[14] Zelin, Charlotte, Wendy K. Chung, Mederic Jeanne, Gongbo Zhang, and Chunhua Weng. “Rare disease diagnosis using knowledge guided retrieval augmentation for ChatGPT.” Journal of Biomedical Informatics 157 (2024): 104702.

[15] Truhn, Daniel, Jorge S. Reis-Filho, and Jakob Nikolas Kather. “Large language models should be used as scientific reasoning engines, not knowledge databases.” Nature medicine 29, no. 12 (2023): 2983–2984.

[16] Goodman, Rachel S., J. Randall Patrinely, Cosby A. Stone, Eli Zimmerman, Rebecca R. Donald, Sam S. Chang, Sean T. Berkowitz et al. “Accuracy and reliability of chatbot responses to physician questions.” JAMA network open 6, no. 10 (2023): e2336483–e2336483.

[17] Ong, Jasmine Chiat Ling, Shelley Yin-Hsi Chang, Wasswa William, Atul J. Butte, Nigam H. Shah, Lita Sui Tjien Chew, Nan Liu et al. “Medical ethics of large language models in medicine.” NEJM AI 1, no. 7 (2024): AIra2400038.

[18] Hendrycks, Dan, Collin Burns, Steven Basart, Andy Zou, Mantas Mazeika, Dawn Song, and Jacob Steinhardt. “Measuring massive multitask language understanding.” arXiv preprint arXiv:2009.03300 (2020).

[19] Jin, Di, Eileen Pan, Nassim Oufattole, Wei-Hung Weng, Hanyi Fang, and Peter Szolovits. “What disease does this patient have? a large-scale open domain question answering dataset from medical exams.” Applied Sciences 11, no. 14 (2021): 6421.

[20] Pal, Ankit, Logesh Kumar Umapathi, and Malaikannan Sankarasubbu. “Medmcqa: A large-scale multi-subject multi-choice dataset for medical domain question answering.” In Conference on health, inference, and learning, pp. 248–260. PMLR, 2022.

[21] Flores-Gouyonnet, Jaime, María C. Cuéllar-Gutiérrez, Gabriel Figueroa-Parra, Bradly Kimbrough, Elena K. Joerns, Erika Navarro-Mendoza, Cynthia S. Crowson, et al. “Performance of large language models in rheumatology board-like questions: accuracy, quality, and safety.” The Lancet Rheumatology (2025).

[22] Chiang, Wei-Lin, Lianmin Zheng, Ying Sheng, Anastasios Nikolas Angelopoulos, Tianle Li, Dacheng Li, Banghua Zhu et al. “Chatbot arena: An open platform for evaluating llms by human preference.” In Forty-first International Conference on Machine Learning. 2024.

[23] Goh, Ethan, Robert Gallo, Jason Hom, Eric Strong, Yingjie Weng, Hannah Kerman, Joséphine A. Cool, et al. “Large language model influence on diagnostic reasoning: a randomized clinical trial.” JAMA Network Open 7, no. 10 (2024): e2440969–e2440969.

[24] Newble, David. “Techniques for measuring clinical competence: objective structured clinical examinations.” Medical education 38, no. 2 (2004): 199–203.

[25] Giuffré, Mauro, Kisung You, and Dennis L. Shung. “Evaluating ChatGPT in medical contexts: the imperative to guard against hallucinations and partial accuracies.” Clinical Gastroenterology and Hepatology 22, no. 5 (2024): 1145–1146.

[26] Richards, Sue, Nazneen Aziz, Sherri Bale, David Bick, Soma Das, Julie Gastier-Foster, Wayne W. Grody et al. “Standards and guidelines for the interpretation of sequence variants: a joint consensus recommendation of the American College of Medical Genetics and Genomics and the Association for Molecular Pathology.” Genetics in medicine 17, no. 5 (2015): 405–423.

[27] Riggs, Erin Rooney, Erica F. Andersen, Athena M. Cherry, Sibel Kantarci, Hutton Kearney, Ankita Patel, Gordana Raca et al. “Technical standards for the interpretation and reporting of constitutional copy-number variants: a joint consensus recommendation of the American College of Medical Genetics and Genomics (ACMG) and the Clinical Genome Resource (ClinGen).” (2020): 245–257.

[28] F Ma, and H P Dai. “Expert consensus on the diagnosis and management of antineoplastic drug-related interstitial lung disease”, Chinese Journal of Oncology 44, no. 7 (2022).

[29] Brahmer, Julie R., Christina Lacchetti, Bryan J. Schneider, Michael B. Atkins, Kelly J. Brassil, Jeffrey M. Caterino, Ian Chau et al. “Management of immune-related adverse events in patients treated with immune checkpoint inhibitor therapy: American Society of Clinical Oncology Clinical Practice Guideline.” Journal of Clinical Oncology 36, no. 17 (2018): 1714–1768.

[30] Writing Committee Members, Elliott M. Antman, Daniel T. Anbe, Paul Wayne Armstrong, Eric R. Bates, Lee A. Green, Mary Hand et al. “ACC/AHA guidelines for the management of patients with ST-elevation myocardial infarction—executive summary: a report of the American College of Cardiology/American Heart Association Task Force on Practice Guidelines (Writing Committee to Revise the 1999 Guidelines for the Management of Patients With Acute Myocardial Infarction).” Journal of the American College of Cardiology 44, no. 3 (2004): 671–719.

[31] Kim, Junyoung, Kai Wang, Chunhua Weng, and Cong Liu. “Assessing the utility of large language models for phenotype-driven gene prioritization in the diagnosis of rare genetic disease.” The American Journal of Human Genetics 111, no. 10 (2024): 2190–2202.

[32] Duong, Dat, and Benjamin D. Solomon. “Artificial intelligence in clinical genetics.” European Journal of Human Genetics (2025): 1–8.

[33] Tordjman, Mickael, Zelong Liu, Murat Yuce, Valentin Fauveau, Yunhao Mei, Jerome Hadjadj, Ian Bolger et al. “Comparative benchmarking of the DeepSeek large language model on medical tasks and clinical reasoning.” Nature Medicine (2025): 1–1.

[34] Sandmann, Sarah, Stefan Hegselmann, Michael Fujarski, Lucas Bickmann, Benjamin Wild, Roland Eils, and Julian Varghese. “Benchmark evaluation of DeepSeek large language models in clinical decision-making.” Nature Medicine (2025): 1–1.

