## supplementary Methods, Tables, and Figures for "An interdisciplinary, randomized, single-blind evaluation of state-of-the-art large language models for their implications and risks in medical diagnosis and management"

*{[1]Dept. Orthopedics, [3]AIBD Lab, [4]Dept. Neurology, [5]Dept. Pediatrics , [6]Dept. Accidents and Emergency, [7]Neonatal ICU (NICU), [8]Dept. Pediatric Surgery, [9]Dept. Prenatal Diagnosis (Clinical Genetics), [10]Dept. Nephrology, [11]Dept. Respiratory Medicine, [12]Intensive Care Unit (ICU), [13]Dept. Cardiac Surgery, [14]Dept. Cardiology}, The University of Hong Kong - Shenzhen Hospital (HKU-SZH), Shenzhen 518053, China. [2]Shenzhen Clinical Research Center for Rare Diseases, Shenzhen 518053, China. [15]Dept. Pediatric Orthopedics, Children's Hospital, Zhejiang University School of Medicine, Hangzhou 310052, China. [16]Dept. Orthopedics, Peking Union Medical College Hospital (PUMCH), Chinese Academy of Medical Sciences, Beijing 100730, China.*

*[*]These authors contributed equally*

*[†]Corresponding authors*

**S2 Supplementary Figures** **15**

**S3 Supplementary Tables** **22**

### List of Figures

S1 Overall design. (A) 13 teams were assembled, consisting of question designers (one for each subject) and independent expert evaluators (at least two for each subject, except for pediatric surgery). Four of the question designers were also involved in evaluating the answers, but their scores were for reference only. Numbers in circle indicate years in medical practices, with years in brackets relating to year the physician obtain the national medical licensing examination (NMLE) , which was enacted in 1999 [1]. (B) Screenshot showing the SOTA models by the end of February 2025 (Feb 27) on a popular leaderboard ARENA. Four models were selected for their API accessibility. (C) Screenshot of our in-house'ly developed web system for model evaluations. . . . 15

S2 Round 1 evaluation consistency and consensus results. (A) Individual evaluators' conformation with their respective group's consensus assessments on four dimensions of the questions: difficulty, rare disease, diagnosis, treatment, procedures in the first round. Names with * are designers or non-independent evaluators. Their assessments serve as references. (B) Among the two types of questions, bar-charts showing the degree of difficulty distribution per subject/team. Numbers on top indicate total number of questions per subject. C-F, Among the two types of questions, bar-charts showing the distributions of rare disease (C), diagnosis (D), treatment plan (E) and operational procedures (F) involved, per subject/team. . . . 16

S3 Quality assessments in round 2. (A) Bar chart illustrating the proportion of questions assigned identical scores by each evaluator. evaluators exhibiting an excessive rate of identical scores (>20%) were deemed non-informative and excluded from the analysis. (B) Bar chart displaying the completion rate of evaluations per evaluator. evaluators who failed to assess a sufficient fraction of the questions were considered non-informative and subsequently disqualified. . . . 17

S4 Fatigue and progress monitoring for Round 2 evaluation. (A) Bar plots depicting the correlation between each evaluator's scores (both independent and non-independent) and model response lengths (measured using Pearson's $r$) across increasing question order. The observed upward trend in correlation along the sequence of questions (as they appeared on the web platform) may suggest the potential influence of evaluator fatigue, leading to a decline in the quality of ratings over time. (B) Cumulative progress curves for each individual evaluator, showing their evaluation progress since the start of the second-round evaluation on March 10 at 8:00 PM local time. The legend identifies evaluators by their initials, and the numbers in parentheses represent the estimated durations required to achieve 5% to 95% of total progress. C. Instances of technical issues are indicated by black asterisks (*), where evaluators reported losing their scores after logging out. These disruptions were likely caused by a restart of the web system during their scoring sessions, leading to drops in their respective progress curves. 18
S5 Round 2 pairwise consistency assessment among intra-subject evaluators. (A) Box-and-whisker plots depicting the distribution of intra-discipline consistency across evaluator pairs, with each plot representing the distribution of scores between two evaluators within the same discipline. (B) Same as (A), but showing Spearman's correlation coefficients. . . . . . . . . . 19
S6 Raw scores on strengths and corresponding raw ranks, by the independent evaluators (OSCE and real-case questions combined) (A) Bar plots showing the raw scores for all subjects combined (topleft panel) or individually (other panels). The numbers in square brackets indicate numbers of records, which are the products of the numbers of qualified questions, models and independent evaluators. (B) Similar arrangements as in (A), except the raw ranks were shown. The raw ranks (range from 1 to 7) are based on the ranking of the seven responses for each question and by each evaluator. Ties were handled with the "average" method. . . . . . . . . . . . . . . . . . . . . . . . . . . . . . . . . . . . . . . . . . . . . 20
S7 Marked baseline variations in round 2 evaluation metrics. (A) Bar plots showing the distribution of response lengths across models. (B) Box-and-whisker plots summarizing strength scores assigned to each model in Round 2. (C) Box-and-whisker plots illustrating score variability across individual evaluators, revealing inter-evaluator differences in scoring tendencies. Initials with asterisks (*) denote non-independent expert evaluators; initials with hash marks (#) indicate question designers providing reference scores, excluded from primary analyses. . 21

### List of Tables

S1 Questions. OSCE: Objective Structured Clinical Examination. realCases: real medical cases selected from HKU-SZH. Numbers in brackets indicate disqualified questions (total 10). . . . 22
S2 Regression estimates and statistical significance for model response length (rlen, in 1,000 characters) as a function of question length (qlen, in 1,000 characters), question type (qtype), language model (model, with "DS32B" as the reference category), and discipline (subject, with "01a.AE" as the reference category). The dataset comprises 4,795 observations, corresponding to 7 responses by 7 models per each of the 685 questions. Significant values ($p < 0.05$) are in bold. . . . . . . . . . . . . . . . . . . . . . . . . . . . . . . . . . . . . . . . 23
S3 Regression Results for Strengths and Strength ranks, with respect to response lengths (rlen), as real-valued (A), ranking of rlen (rlenrank) as integers (B), and ranking of rlen as categorical (C) variables, respectively. . . . . . . . . . . . . . . . . . . . . . . . . . . . . . . . . . . . 24
S4 Summary of multiple linear regression results for evaluation raw scores (ranging from 0 to 10, with higher scores indicating better performance) in relation to a set of covariates, including language model (model), discipline (subject, with "01a.AE" as the reference category), question type (qtype, with "OSCE" as the reference), question difficulty (difficulty, with "easy" as the reference category), and factors related to rare diseases, diagnosis, treatment, and operational procedures, across all independent evaluators, with "no" as the reference category for the last four covariates. Model 1a includes 9,856 records (setA) from 27 independent qualified evaluators. Model 1b was fitted with the same covariates but on setABC, which included these 9,856 records, 1,824 records from 4 non-independent evaluators, and 315 records from question designer for the pediatric surgery team (12.PediSurg), where there is only one evaluator. . . . . . . . . . . . . . . . . . . . . . . . . . . . . . . . . . . . . . . . . . . . . 25

S5 (A) Results of multiple linear regression models predicting evaluator-assigned raw scores (range: 0–10, higher indicating better performance) for medical strength and quality across all independent evaluators, subjects, and question types. The variable rlenrank represents model response length rankings (1 to 7, with higher values indicating longer responses); ranks 1.5, 3.5, and 4.5 reflect tied positions. The covariate uses as the reference level. References: DS32B, 01a.AE, easy for model, subject, covariates, and difficulty, respectively. Rare disease involvement, diagnostic content, treatment planning, and procedural content all have (no) as the reference . Results are reported as point estimates with 95% confidence intervals (C.I.) and associated p-values. Model 2a and Model 2b were fitted on setA and setABC, respectively. (B) ANOVA comparison of Models. ANOVA, Analysis of Variance; AIC: Akaike Information Criterion; RSS: Residual sum of squares; DF: Degree of freedom. . . . . . . . . . . . . . . . . . 26
S6 Multiple linear regression results for evaluator-assigned per-question rank scores (1–7; higher = better) across all questions, evaluators, and subjects. rlenrank denotes model response length ranks (1–7; higher = longer), with ties at 1.5, 3.5, and 4.5. The reference levels are: subject (01a.AE), qtype (OSCE), question difficulty (easy), and absence of rare disease, diagnosis, treatment, and procedure content (no for each). Estimates are shown with 95% confidence intervals and p-values. . . . . . . . . . . . . . . . . . . . . . . . . . . . . . . . . 27
S7 A result table summarizing the multiple linear regression results for the per-question evaluation ranks (1 to 7, higher is better) with respect to question type (qtype), models (model, with “DS32B” as the reference), disciplines (subject, with 01.AE as the reference), difficulty (with easy as the reference), rare diseases, diagnosis, treatment and operational procedures. The reference levels for the last covariates are all [no]. The 9,856 records cover all independent evaluators' assessments on the model responses' medical strength and quality. . . . . . . . . . 28
S8 Linear mixed-effects modeling of raw strength scores, on setA and setABC. ICC: Intraclass Correlation Coefficient. References: rlenrank [1.0], qtype: OSCE, subject: 01a.AE, difficulty: easy, rare: no, diagnosis: no, treatment: no, procedure: no. Model LME1 and Model LME2 were fitted on SetA and SetABC, respectively. . . . . . . . . . . . . . . . . . . . . . . . . . . . 29
S9 Comparison of linear model (LM) and linear mixed-effects model (LME). . . . . . . . . . . . 30
S10 Fixed effects estimates and odds ratios with 95% confidence intervals from logistic mixed model predicting excellence (score $\geq 8$), adjusted for evaluator random effects . . . . . . . . . 30
S11 Fixed effects estimates and odds ratios with 95% confidence intervals from logistic mixed model predicting excellence (score $\geq 8$), adjusted for evaluator random effects . . . . . . . . . 31
S12 Fixed effects estimates and odds ratios (OR) with 95% confidence intervals from generalized linear mixed model predicting failure (score $\leq 4$), adjusted for evaluator random effects. . . . 32
S13 Fixed effects estimates and odds ratios with 95% confidence intervals from logistic mixed model predicting failure (score $\leq 4$), adjusted for evaluator random effects and response lengths. Note that there were 714 instances of incompetence, causing fitting of the full model below difficult. . . . . . . . . . . . . . . . . . . . . . . . . . . . . . . . . . . . . . . . . 33
S14 Pearson correlation matrix between LLMs when they were asked to score the responses, based on 18,768 data-points from scores on 4,692 responses (belonging to 675 questions). LLMs were asked to rate all seven responses per question, which included their own responses in the previous round. . . . . . . . . . . . . . . . . . . . . . . . . . . . . . . . . . . 33
S15 Linear regression results for LLM-rated scores. . . . . . . . . . . . . . . . . . . . . . . . . 34

### S1 Supplementary Methods

#### S1.1 Testing languages

In alignment with clinical practice in China, all questions were presented in Chinese, and model responses were also in Chinese.

#### S1.2 Strength score definition

The strength is a measure of medical plausibility [1], as below:

| Corresponding scores | China | US | UK | Approx. # Years After Graduation |
|---|---|---|---|---|
| 0 | | no medical knowledge | | no med edu |
| 0.5 or 1.5 | | general knowledge | | no med edu |
| 2-3.5 | | fresh medical graduates | | 0 |
| 4-5.5 | 住院医师 (Resident Doctor) | Intern | Foundation Year 1 Doctor (FY1) | 0–1 years |
| 4-5.5 | 住院医师 (Resident Doctor) | Resident | Foundation Year 2 Doctor (FY2) | 1–2 years |
| 6-6.5 | 主治医师 (Attending physician) | Fellow (Specialist Training) | Core Trainee (CT1–CT2/3) | 3–6 years |
| 7-8 | 副主任医师 (Assoc chief physician) | Attending physician (Specialist Training) | Specialty registrar (ST3–ST6/7) | 6–10 years |
| 8.5-9 | 主任医师 (Chief physician) | Senior Attending / Consultant | Consultant (ST3–ST6/7) | 10+ years |
| $\geq$ 9.5 | leading experts | leading experts | leading experts | 10+ years |

#### S1.3 Question designs for A.E.

To comprehensively assess LLMs in emergency setting, we developed a customized Objective Structured Clinical Examination (OSCE) aligned with the expected competence of graduating emergency medicine residents. The OSCE comprised 30 stations, each designed and rigorously reviewed by a panel of clinical experts to reflect authentic acute and emergency (A.E.) scenarios. Stations targeted eight core competencies: (1) history-taking and clinical decision-making, (2) physical examination, (3) clinical teaching, (4) procedural skills, (5) communication, (6) resuscitation, (7) psychiatric emergencies, and (8) clinical management. Standardized, expert-validated scoring rubrics were applied across all stations to ensure objective, reproducible evaluation. Within this framework, the LLM was required to independently interpret clinical inputs and perform as an emergency medicine specialist.

In parallel, we designed two sets of real cases admitted to the A.E. department :

- **01a.AE**: 72 consecutive real cases from patients admitted to A.E. of HKU-SZH in January 2025. These cases were selected from complete clinical records. Case inclusion emphasized diagnostic complexity and clinical relevance to capture the breadth of real-world emergency presentations.
- **01b.AE**: The 20 real-world cases were drawn from patients admitted to the resuscitation area of the emergency department at HKU-SZH in January 2025, following emergency stabilization. These included: cerebrovascular events (n=4; stroke and intracerebral hemorrhage), trauma (n=3; including falls and blunt injuries), cardiovascular emergencies (n=2; acute myocardial infarction and hypertensive crisis), respiratory emergencies (n=2; aspiration pneumonia and COPD exacerbation), sepsis (n=2), gastrointestinal bleeding (n=1), pediatric emergencies (n=4; including HMPV infection, influenza, acute laryngitis, and aspiration), and toxicological emergencies (n=2; drug overdose and poisoning).

Both sets of A.E. real cases were ultimately hospitalized. Inclusion criteria for real cases required de-identified data and representation of core emergency medicine syndromes (e.g., chest pain, trauma, poisoning) and key resuscitation competencies to enhance external validity. Each case contained a complete clinical information chain, including chief complaint, history of present illness, past medical history, physical examination, and initial laboratory or imaging results. Cases involving unresolved medicolegal issues or insufficient de-identification were excluded.

For testing, the LLM was again instructed to function as an emergency physician. De-identified case data were input in a single prompt, including patient demographics, chief complaint, history, physical exam findings, and initial test results. The model was then asked to generate a preliminary diagnosis, diagnostic rationale, and management plan. Model outputs were evaluated for clinical accuracy and decision-making ability in the context of acute, high-acuity presentations.

##### S1.4 Question designs for cardiology

A total of 45 cardiology cases were selected from patient records at the Department of Cardiology, HKU–SZH, spanning the past five years and covering patients aged 18 to 78 years. Of these, 30 were standardized OSCE-style cases and 5 were real clinical cases.

The OSCE cases were developed from typical inpatient presentations of common cardiovascular conditions, including acute myocardial infarction, acute heart failure, cardiac arrest, atrial fibrillation with Wolff–Parkinson–White syndrome, pericardial effusion, and pericarditis. These cases were standardized by cardiology faculty and used to assess core clinical competencies of resident trainees. Given their structured format and clear diagnostic pathways, current LLMs are expected to perform reliably on this subset.

In contrast, the real-case scenarios represented diagnostically complex or therapeutically ambiguous cases involving multisystem comorbidities, unresolved diagnoses, or conflicting treatment options. These cases required nuanced, individualized clinical reasoning, for which current LLMs are expected to have significant limitations. Accordingly, clinical decision-making—particularly by senior physicians—remains irreplaceable at this stage. However, with ongoing advances in model development, LLMs may eventually assist in streamlining complex clinical workflows and supporting physicians in managing challenging cases.

##### S1.5 Question designs for cardiac surgery

A total of 58 cardiac surgery examination items, collected from admissions between 2024 and 2025, were included. These comprised 33 OSCE scenarios and 25 real patient cases. Patient data underwent rigorous anonymization and data privacy protection procedures prior to analysis.

**OSCE design** The 33 OSCE items were collaboratively developed by cardiac surgery professors, consultants, and associate consultants. The content was designed to reflect common clinical scenarios encountered by adult cardiac surgeons, encompassing coronary artery disease, valvular heart disease, great vessel disease, and congenital heart defects. Additionally, comprehensive clinical scenarios involving coexisting conditions such as infections, hyperthyroidism, and pregnancy were included to evaluate the candidates' integrative management capabilities. A unique rare-disease scenario involving Marfan syndrome was also presented. The examination emphasized core competencies required for cardiac surgeons, assessing multiple dimensions including patient history-taking, doctor-patient communication skills, diagnostic and differential diagnostic abilities, as well as comprehensive therapeutic decision-making and professional knowledge.

**Real-case clinical scenarios** The 25 real-case scenarios consisted of patients diagnosed primarily with valvular heart diseases who required surgical intervention. All cases were inpatients from HKU-SZH in the past year (2024). The primary aim was to assess A.I. algorithms in their capability to diagnose disease accurately, determine optimal surgical strategies, evaluate perioperative risks, select appropriate valve types, and manage perioperative care comprehensively. Participants or A.I. algorithms were prompted to answer structured clinical questions including: primary diagnosis, recommended treatment approach, appropriate surgical technique, surgical risk assessment with corresponding mitigation strategies, decision-making between valve repair or replacement, and valve type selection (bioprosthetic vs. mechanical). Additionally, participants assessed the feasibility of minimally invasive surgical approaches. The case spectrum included mitral valve disease, aortic valve disease, and tricuspid valve disease, with most cases characterized by high clinical complexity involving concurrent cardiac pathologies such as ascending aortic aneurysm, coronary artery disease, and atrial fibrillation. These complexities required simultaneous surgical management, thereby rigorously evaluating AI's capability to handle intricate clinical scenarios and adhere strictly to appropriate surgical indications and evidence-based practice guidelines.

##### S1.6 Question designs for neurology

The primary objective of the neurology panel was to evaluate the capability of SOTA LLMs to perform at the level of a board-certified neurologist, defined as a physician who has successfully completed standardized residency training in neurology, in the diagnosis and management of neurological disorders. To achieve this, a comprehensive assessment was designed, comprising two components:

- real-world clinical cases (40 questions) retrieved from hospital information system of HKU-SZH.
- OSCE, including imaging interpretation and related clinical knowledge analysis (8 questions), and simulated patient consultations (4 questions).

The clinical cases consisted of 40 questions, from inpatient cases at HKU-SZH. We provided demographic information (age, gender), a concise medical history including presenting illness, past medical history, relevant personal and family history, key physical examination findings, and significant laboratory and imaging reports without images. Models were required to provide a diagnosis, diagnostic rationale, and a treatment plan. The cases encompassed a broad spectrum of neurological disorders, including common and rare diseases, organic and functional conditions, and acute and chronic illnesses. Specific diseases included Alzheimer's disease, acute ischemic stroke, amyotrophic lateral sclerosis, autoimmune GFAP astrocytopathy, myasthenia gravis, multiple sclerosis, multiple system atrophy, neuromyelitis optica spectrum disorder (NMOSD), Parkinsonian syndrome, anti-NMDAR antibody encephalitis, Parkinson's disease, epilepsy, Huntington's disease, essential tremor, hepatic encephalopathy, cardiogenic syncope, psychogenic non-epileptic seizures, migraine with aura, Lewy body dementia, subacute combined degeneration, viral meningitis, Creutzfeldt-Jakob disease, idiopathic intracranial hypertension, abducens nerve palsy, multiple cranial neuritis, benign paroxysmal positional vertigo, chronic inflammatory demyelinating polyneuropathy, Guillain-Barré syndrome, dermatomyositis, and carpal tunnel syndrome.

The imaging interpretation and clinical knowledge analysis component included 8 questions, each providing detailed descriptions of typical imaging findings without images to assess the model's ability to interpret radiological manifestations. Models were further evaluated on their understanding of related clinical issues, such as treatment principles for the identified condition. The simulated patient consultations comprised 4 questions, designed to mimic a physician's clinical encounter by presenting brief clinical scenarios. Models were tasked with identifying key history-taking points, physical examination priorities, a preliminary diagnosis, and a management plan, with each simulation allowing only a single round of interaction.

The scoring system prioritized diagnostic accuracy as the primary endpoint. The appropriateness of medical recommendations was also assessed, focusing on the diagnostic rationale, such as justification based on clinical data, and the proposed treatment plan, including alignment with evidence-based guidelines. Responses were evaluated for clinical relevance, coherence, and adherence to standard neurological practice.

##### S1.7 Question designs for nephrology

A total of 10 real-world nephrology cases were selected from patients treated at HKU-SZH over the past three years. These cases encompassed a range of kidney-related conditions, including chronic glomerulonephritis, acute kidney injury and chronic kidney disease post-renal transplantation, autoimmune-related nephritis, nephrotic syndrome complicated by both chronic kidney disease and acute kidney injury, autosomal dominant polycystic kidney disease (ADPKD), and hypophosphatemia.

Several cases were intentionally chosen to reflect real-world diagnostic uncertainty, such as glomerulonephritis in patients who declined renal biopsy. In such scenarios, nephrologists often rely on inferential reasoning to make provisional diagnoses and initiate empirical treatment. These cases were designed to assess whether LLMs could synthesize available clinical information—including history, physical findings, and laboratory data—to reasonably infer the underlying pathology and propose appropriate next steps in investigation and management based on current guidelines.

One case of hypophosphatemia was included to evaluate whether LLMs could identify a rare but clinically relevant etiology. In this case, extensive literature review by the nephrology team led to the identification of an underlying tumor as the cause; the patient's condition improved significantly following surgical resection. This case was used to examine whether LLMs could, when provided with relevant clinical history, draw upon up-to-date literature and reasoning to suggest a correct diagnostic pathway.

Additionally, several cases involving ADPKD were included. The diagnosis and management of ADPKD have evolved rapidly in recent years, with multiple international expert consensus statements published. In January 2025, the first KDIGO guideline on ADPKD was released. While the latest LLMs may not yet have integrated the KDIGO 2025 guideline into their training data, expert consensus prior to its release already provided a solid framework for clinical decision-making. These cases were used to assess the models' ability to: (1) recognize early diagnostic features of ADPKD, (2) predict disease progression, (3) determine eligibility for tolvaptan therapy based on patient history, and (4) provide appropriate reproductive counseling.

An additional 30 OSCE-style questions were selected from the internal training and assessment materials used for nephrology residents. These questions focused on common and high-frequency conditions in nephrology, including acute kidney injury, end-stage renal disease, nephrotic syndrome, hypertensive nephropathy, gouty nephropathy, urinary tract infections, IgA nephropathy, and diabetic nephropathy.

These cases were designed to evaluate whether LLMs can simulate clinical reasoning in a format aligned with real-world physician workflows. Specifically, models were assessed on their ability to: (1) identify key points in history-taking, (2) prioritize relevant physical examinations, (3) select appropriate diagnostic tests, (4) assess disease severity and risk, and (5) provide reasonable and guideline-consistent management recommendations. The aim was to determine if LLMs can emulate the stepwise, clinically oriented decision-making process expected of trained nephrology residents.

##### S1.8 Orthopedics case design

A total of 52 orthopedic cases were included, consisting of 37 structured scenarios (OSCE-type questions) and 15 real clinical cases derived from inpatients admitted between 2023 and 2025. These cases were curated to ensure comprehensive coverage across all major orthopedic subspecialties, including spine, trauma and joint, sports medicine, pediatric orthopedics, and musculoskeletal oncology.

**Structured OSCE cases** All OSCE-type questions were randomly sampled from the internal Orthopedic Specialist Examination Bank of HKU-SZH, a repository not accessible to the public. The scenarios were jointly designed by orthopedic professors, consultants, and associate consultants in alignment with the *CanMEDS* Roles framework established by the Royal College of Physicians and Surgeons of Canada. Case topics included:

- Spine (n=9): lumbar disc herniation, lumbar spondylolysis with spondylolisthesis
- Trauma and Joint (n=9): knee osteoarthritis, stenosing tenosynovitis, post-TKA complications, patellar fracture, avascular necrosis of the femoral head, lateral epicondylitis
- Sports Medicine (n=8): anterior cruciate ligament (ACL) tear, meniscal tear
- Pediatrics (n=6): scoliosis, femoral neck fracture
- Oncology (n=5): giant cell tumor of bone

Each case was presented in a clinical scenario format, assessing diagnostic accuracy, differential diagnosis, disease severity assessment, treatment principles, and management planning. These cases primarily reflect common conditions encountered during residency training and aim to evaluate whether LLMs can perform at the level of a junior orthopedic trainee.

**Real clinical cases** Fifteen real cases were extracted from inpatient records at HKU-SZH (2023–2025) and anonymized to remove all identifiable patient information. These cases were intentionally selected to complement the OSCE set in both scope and difficulty, including both typical and complex presentations across subspecialties:

- Spine (n=4): spinal infections, atypical cervical spondylotic myelopathy, lumbar stenosis with compression fractures and kyphosis, ankylosing spondylitis with anterior dural sac ectasia
- Trauma and Joint (n=6): femoral neck fracture, knee osteoarthritis, carpal tunnel syndrome, ankylosed hips in ankylosing spondylitis, Charcot joint, polytrauma from high fall
- Pediatrics (n=2): syndromic clubfoot, cerebral palsy
- Sports Medicine (n=2): rotator cuff injury, meniscal tear
- Oncology (n=1): chordoma

Each real-case scenario required LLMs to provide diagnostic reasoning, recommend appropriate investigations, and suggest treatment plans. Some cases, such as atypical cervical myelopathy, Charcot arthropathy, polytrauma, and rare tumors, were purposefully chosen to exceed the expected competency of junior trainees, serving instead to evaluate whether LLMs can approximate consultant-level diagnostic and therapeutic acumen.

**Advanced clinical reasoning prompts** Several cases included challenge questions to assess the LLM's ability to reason through current clinical controversies. Examples included: whether to prioritize total hip arthroplasty or spinal osteotomy in ankylosing spondylitis with fused hips and kyphotic deformity; updated management strategies for Charcot joints; and clinical decision-making in ankylosing spondylitis with dural sac expansion. These complex prompts assessed the model's capacity to track evolving medical literature and provide evidence-based, justifiable opinions.

**Overall design considerations** The combined use of OSCE-style cases and real-world clinical scenarios enabled a stratified assessment of LLMs across a range of diagnostic complexities. By diversifying question types and clinical domains, the design aimed to efficiently probe the diagnostic and management capabilities of LLMs within the orthopedic specialty.

##### S1.9 Question designs of adult ICU

A total of 55 assessment items were developed for the ICU evaluation, comprising 40 OSCE questions and 15 real-case scenarios derived from actual ICU admissions between 2023 and 2025 at HKU-SZH. The question design was guided by gold standards including the *Specialty Training Textbook of Critical Care Medicine* published by the Chinese Medical Association and the *National Qualification Examination Textbook for Intermediate-Level Critical Care Medicine*. The content spans all major sub-specialties within critical care, including shock, sepsis, hemodynamics, mechanical ventilation, severe infections, trauma, and nutritional support.

Structured OSCE questions were randomly selected from an internal examination database developed by Dept. Critical Care Medicine at HKU-SZH. All items were designed collaboratively by consultant and associate consultant intensivists. The topics cover foundational knowledge in pathophysiology (6 items), acute respiratory failure and respiratory support (6 items), shock and hemodynamic analysis (4 items), coagulation disorders and trauma-related scenarios (3 items), and integrated clinical case analysis (8 items).

Most questions were case-based and focused on clinical reasoning, encompassing aspects such as diagnosis and differential diagnosis, disease severity assessment, therapeutic principles, and treatment planning. These items are commonly used to assess residents in standardized training programs as well as physicians preparing for ICU specialization exams. The scenarios primarily reflect high-frequency and high-impact ICU conditions, particularly respiratory failure, airway management, hemodynamic monitoring, renal replacement therapy, sedation and analgesia, and central nervous system complications. Although not exhaustive, the design aimed to comprehensively assess clinical decision-making, evidence-based practice, and professional cognition, aligning with the core competencies expected of advanced-level ICU physicians.

The 15 real-case scenarios were constructed from anonymized clinical data of adult ($> 18$ years old) ICU inpatients between 2023 and 2025. These cases were selected to complement the OSCE items in both content and difficulty, and to represent a wide range of subspecialties. Each case was crafted to simulate real-world diagnostic and therapeutic challenges, including mechanical ventilation settings, hemodynamic interpretation, and advanced pharmacological or pathophysiological knowledge. Many of these cases exceeded the expectations for junior physicians and were designed to challenge even consultant-level intensivists.

These real cases were used to evaluate whether the AI system could demonstrate clinical reasoning capabilities at a consultant level. The scenarios involved complex, often ambiguous clinical conditions. Examples include perioperative management of patients with myasthenia gravis secondary to thymoma, anticoagulation dilemmas in advanced cancer patients with concurrent bleeding and thrombosis risks, and end-stage patients with refractory gastrointestinal bleeding due to erosive esophagitis without surgical options. Additional complexity stemmed from diagnostic uncertainty, treatment conflicts, organ dysfunction, and controversies in current medical practice.

Some scenarios were purposefully constructed around ongoing academic debates to test the AI's ability to engage with cutting-edge medical discussions and to articulate evidence-based viewpoints. The assessment thus encompassed both structured and open-ended items to evaluate the AI's proficiency in diagnostic accuracy, therapeutic planning, clinical reasoning, and its potential to provide individualized, goal-directed care in high-stakes critical care environments.

By combining structured OSCE with real-case scenarios and employing diverse questioning styles, we aimed to simulate realistic ICU settings and evaluate the AI model's core competencies in critical care medicine. The focus was placed on prevalent and high-impact conditions such as sepsis, shock, ARDS, acute kidney injury, renal replacement therapy, and mechanical ventilation. This multidimensional evaluation allowed for preliminary assessment of the model's clinical aptitude across diagnosis, differential diagnosis, decision-making, treatment optimization, quality control, and research guidance. It also offered insight into the model's potential to provide individualized care strategies for critically ill patients with complex conditions.

##### S1.10 Question designs for pediatrics

A total of 45 pediatric cases were included, derived from real patient admissions to the Department of Pediatrics at HKU–SZH between 2018 and 2023. Patient ages ranged from 28 days to 18 years.

There were 15 real-case scenarios, selected to represent diagnostically and therapeutically relevant cases encountered in pediatric inpatient care over the past three years. Of these, 7 involved rare diseases across pediatric subspecialties including endocrinology, nephrology, immunology, and hematology. Diagnoses included Noonan syndrome, type III Gaucher disease, Bartter syndrome type II, Schimke immuno-osseous dysplasia (SIOD), hereditary spherocytosis, IMAGE-I syndrome, and hemophagocytic lymphohistiocytosis. Three of these cases included whole-exome sequencing data, one provided histopathology, and the remaining were supported by characteristic clinical features or laboratory results. The other 8 real-case scenarios involved complex but non-rare conditions such as Kawasaki disease, exogenous lipoid pneumonia, acute bacterial meningitis, type 1 diabetes mellitus, germ cell tumor, nephrotic syndrome, acute pancreatitis, and acute leukemia. Two of these included histopathological confirmation, while the rest were supported by definitive history, classic clinical presentation, and relevant laboratory findings. The design rationale assumed that LLMs should achieve highly accurate diagnosis in genetically confirmed cases and offer more precise treatment recommendations in scenarios with clearly defined clinical features (but without genetic information). These cases were curated to assess the extent to which LLMs could support clinical decision-making in complex, real-world pediatric contexts.

An additional 30 cases were selected in OSCE format from the pediatric examination and competition item bank used for postgraduate year (PGY) 1–3 residents at the same institution. These were standardized cases based on common pediatric inpatient diagnoses from the past five years, including Mycoplasma pneumonia, rotavirus gastroenteritis, hand-foot-mouth disease, acute urinary tract infection, scarlet fever, and diabetic ketoacidosis. All cases were constructed and validated by senior pediatric faculty to assess residents' applied clinical reasoning, history-taking, communication, and procedural skills.

Due to the age-specific pharmacologic considerations, the indirect nature of pediatric history-taking, and the specialized communication skills required in pediatrics, current LLMs are expected to demonstrate limitations in this domain. While physician expertise remains essential and irreplaceable at present, advances in model training and alignment may, in the near future, enable AI systems to assist in streamlining routine pediatric clinical tasks.

##### S1.11 Question designs for prenatal medicine

Two assessment formats were employed. OSCE-style questions (n=12) followed a standardized clinical framework covering chief complaint, history, physical examination, and relevant specialist evaluations. Real-case assessments , based on anonymized clinical records, included comprehensive clinical data such as genetic testing and karyotype analysis. This design enabled evaluation of both structured clinical reasoning and diagnostic depth in complex prenatal genetic contexts.

Prenatal diagnostic cases were selected from 34 patients presenting with clinically suspected genetic disorders between January 2020 and January 2025, with inclusion limited to those with confirmed pathogenic or likely pathogenic genetic variants. Cases were excluded if they involved a single, typical phenotype limited to one organ system (e.g., osteogenesis imperfecta, non-syndromic hearing loss), lacked definitive genetic results, or presented with isolated fetal findings such as increased nuchal translucency or amniotic fluid abnormalities.

##### S1.12 Neonatal Intensive Care Unit (NICU) case designs

A total of 40 NICU questions were designed in the evaluation, comprising 30 OSCE-style questions and 10 real-patient cases. The real cases were drawn from neonates admitted to the NICU at HKU–SZH between December 1, 2023, and February 20, 2025, ranging in age from 14 minutes to 26 days post-birth.

**OSCE-Style case design** Of the 30 OSCE questions:

- Sixteen were adapted from the institutional NICU specialist examination bank and jointly designed by NICU professors, consultants, and associate consultants. These scenarios incorporated the *CanMEDS Roles* framework (Canada) and focused on classic neonatal conditions such as necrotizing enterocolitis, neonatal sepsis, and intraventricular hemorrhage in preterm infants. Each question was designed to assess a range of competencies—including history-taking, communication, diagnosis, differential diagnosis, and integrated management—through multidimensional lenses such as communicator, collaborator, and manager roles. Questions emphasized evidence-based practice, adult learning principles, and competency-based milestones reflective of advanced neonatal subspecialty training.

- Eight questions addressed common neonatal conditions, including neonatal pneumonia, jaundice, and the management of small-for-gestational-age infants. These evaluated core clinical reasoning expected of early-stage neonatal residents.

- Six simulation-based scenarios tested adherence to the *2020 Neonatal Resuscitation Program (NRP)* guidelines, focusing on situational judgment and crisis resource management skills.

**Real-case design** Ten real-world NICU cases were selected to assess model performance across a spectrum of clinical complexities:

- Common conditions: such as transient tachypnea of the newborn, neonatal jaundice, and neonatal infections were used to evaluate standardized decision-making and protocol application (e.g., phototherapy thresholds).
- Critical conditions: included meconium aspiration syndrome, persistent pulmonary hypertension, extremely low birth weight infants, neonatal meningitis, and perinatal asphyxia. These tested the models' ability to handle complex, high-risk decisions.
- Rare conditions: such as tracheoesophageal fistula and teratoma were selected to examine the models' diagnostic sensitivity and completeness in differential diagnosis under unfamiliar presentations.

This case set was designed to explore the applicability and limitations of LLMs in neonatal intensive care. It enabled head-to-head comparisons between LLMs and physicians in clinical reasoning, decision-making, and contextual simulation. The tiered design—spanning resident- to specialist-level tasks, and common to rare case complexity—also allowed evaluation of LLMs' potential in medical education. In particular, the study assessed whether LLMs could enhance clinical training by supporting junior doctors and encoding expert-level reasoning into advanced learning modules.

##### S1.13 Question designs for respiratory medicine

The respiratory medicine dataset comprised 50 cases, including 20 real-patient cases and 30 exam-style cases.

The 20 real cases were collected from hospitalized patients in the Department of Respiratory Medicine at the HKU–SZH between August 2024 and February 2025, with equal representation of male and female patients (10 each). Diagnoses spanned both common and rare respiratory conditions. Infectious diseases included pneumonia in immunocompromised hosts, viral pneumonia, viral pneumonia with myocarditis, aspiration pneumonia, sepsis, tuberculous pleuritis, pulmonary mycosis, pulmonary tuberculosis, lung abscess, parasitic infection, typhoid fever, and urinary tract infection—each with one case. Other diagnoses included bronchial asthma, hyperventilation syndrome (two cases), eosinophilic lung disease, cardiogenic pulmonary edema, benign bronchial tumor, anti-tumor drug–related interstitial lung disease, and autoimmune-associated interstitial lung disease—each with one case unless otherwise specified. All cases were confirmed using current gold-standard diagnostic criteria and presented in structured vignettes including patient age, sex, occupation, symptoms, history of present illness, relevant past/personal/family history, physical examination, and key laboratory, imaging, or pathological findings. The design aimed to replicate real-world clinical complexity and enable the evaluation of AI diagnostic reasoning and treatment planning.

The 30 exam-style cases were selected from national certification question banks for resident physician qualification, clinical licensure, and attending physician board examinations. These cases included 21 male and 9 female patients and emphasized common and diagnostically challenging respiratory conditions. The set included: four cases of acute exacerbation and complications of chronic obstructive pulmonary disease; two cases of bronchial asthma; and twelve cases of infectious diseases, including pulmonary tuberculosis, pneumonia in immunocompromised hosts, aspiration pneumonia, tuberculous pleuritis, lung abscess, and viral pneumonia with myocarditis. Additionally, there was one case each of acute respiratory distress syndrome, lung cancer, pulmonary embolism, post-embolism syndrome, organizing pneumonia, pneumoconiosis, myasthenia gravis–related respiratory failure, idiopathic pulmonary arterial hypertension, idiopathic pulmonary fibrosis, tracheal foreign body, and cardiogenic pulmonary edema. These cases were designed to assess clinical reasoning and differential diagnosis, while acknowledging that LLMs, unlike human trainees, lack test-taking experience and direct patient interaction. As such, their ability to interpret nuanced history and physical exam findings may be limited. The question set was developed with graduated difficulty to support a comprehensive evaluation of LLM performance across a range of diagnostic and clinical decision-making tasks.

##### S1.14 Pediatric surgery case design

A total of 45 questions were developed for pediatric surgery, including 30 OSCE-type structured scenarios and 15 real patient cases. All cases were drawn from the clinical archives of the past three years at a tertiary pediatric surgical center, targeting core competencies required during the first to third years of standardized residency training in pediatric surgery.

The question topics spanned a wide range of common and rare conditions across subspecialties, including gastrointestinal surgery, hepatobiliary surgery, urology, cardiothoracic surgery, and neonatal surgery.

Representative conditions included appendicitis, intussusception, volvulus, intestinal atresia, Meckel's diverticulum, esophageal atresia, Hirschsprung disease, anal atresia, sacrococcygeal teratoma, umbilical hernia, inguinal hernia, hydrocele, biliary atresia, burn injuries, urinary tract infections, hypospadias, posterior urethral valves, hyperhidrosis, and congenital malrotation.

**OSCE scenario design** The 30 OSCE questions emphasized anatomical and embryological foundations, emergency management, pathophysiology, and diagnostic imaging, aiming to evaluate comprehensive clinical reasoning. For example:

- Question 1 tested understanding of embryological defects underlying volvulus and intestinal atresia.
- Questions 4 and 9 examined the relationship between anatomical location, complications, and differential diagnosis of gastrointestinal bleeding.
- Question 5 integrated anatomical landmarks with surgical key points to prevent recurrence.
- Questions 8 and 10 assessed the integration of chromosomal loci, imaging, and functional testing, representing gene-anatomy-function triads.
- Questions 3, 11, and 27 evaluated acute surgical management and early recognition of complications.
- Questions 16 and 18 emphasized the role of imaging in distinguishing pathophysiological states.
- Questions 23 and 30 focused on evaluation of disorders of sexual development.
- Questions 26 and 29 assessed delayed diagnosis of neonatal conditions.

**Real-case scenarios** The 15 real-case questions reflected real-world complexities, including individualized decision-making and multimodal evaluation. Topics addressed:

- Timing of surgery based on age and anatomy
- Gender differences in surgical technique
- Integration of imaging error analysis and functional rehabilitation
- Acute burn management and long-term care
- Use of neonatal screening and tumor markers
- Links between localized infection and systemic sepsis
- Etiologic patterns by age and the prioritization of non-operative treatment
- Differentiation between functional and structural disorders
- Correlation between anatomical development and classical symptom presentation

**Assessment strategy** All questions were designed to promote multidimensional clinical reasoning across embryology, anatomy, pathophysiology, and diagnostic logic. The structure followed the chain of "symptom–examination–diagnosis–treatment," reflecting real clinical decision-making pathways. To evaluate LLMs on realistic medical challenges, we intentionally increased question difficulty by omitting certain clues and required LLMs to interpret surgical and procedural language in a medically precise manner. This strategy assessed both the breadth and depth of LLM understanding in pediatric surgical contexts.

##### S1.15 Question designs for rare diseases

Forty-five real cases from the pediatric orthopedic department were selected as real-case questions for rare diseases. These were cases were inpatients seeking medical services at HKU-SZH in the past 5 years, and cover osteogenesis imperfecta (OI) subtypes I (COL1A1/2), III (COL1A1/2), IV (COL1A1/2), V (IFITM5), X (FKBP10), and XV (Wnt), hypophosphatasia (HPP), pseudoachondroplasia (PSACH), mucopolysaccharidosis Type II (MPS II, Hunter syndrome), developmental dysplasia of the hip (DDH), spondylometaphyseal dysplasia, spastic cerebral palsy, achondroplasia (ACH), fibrous dysplasia, spinal muscular atrophy (SMA II), spondyloepiphyseal dysplasia congenita (SEDC). These conditions are among the more common diseases in rare disease. Patient age, gender, and physical examination reports as well as patient natural history, including fractures, bone mineral densities (BMDs), surgical histories, Cobb angle, lower-limb discrepancies (LLDs), and drug histories are included in the questions. Some information of genetic mutations, were intentionally withdrew (removed) while others were intentionally kept before submitting to APIs. The evaluators were notified of such moves. The reason for this design is that genetic mutations are highly effective in making correct diagnosis, especially for rare diseases. We expect LLMs to be highly capable in utilizing such information to make correct decisions, and intended to test this hypotheses. As such, these two types of moves (removing and keeping genetic information) serve as negative and positive controls, respectively.

Since pediatric orthopedic residents are trained under the orthopedic department, we did not have a separate set of OSCE for rare diseases for this subject.

##### S1.16 Prompt designs

Each question was strengthened with an emphasis on the role and subject of the specialty, with a sentence of the form "your are a specialist in [subject], please present a professional diagnostic analyses and a treatment plan for the above case", before feeding into the models via APIs (application programming interfaces).

##### S1.17 Model query

The questions was fed into the models via APIs on March 8 and 9 2025, from several workstations at HKU-SZH. The code was available on github [https://github.com/HKUSZH/LLMMed/tree/main/round2]

##### S1.18 Local deployment of models

Gemini 2.0, DeepSeek-R1, Qwen Max were accessed via official APIs, while ChatGPT-4o, which was not directly available to the authors, were accessed via third-party proxies. Three models were deployed locally, including DeepSeek R1 32B [DS32B], DeepSeek R1 70B [DS70B], and QwQ 32B: latest [QWEN 32B]. The models were deployed via [ollama] with direct model downloads from the ollama website. The DS32B was deployed on a Dell t7960 machine installed with Linux Ubuntu with an nVidia A5000 Ada Generation GPU. The DS70B was deployed on a Dell t7960 machine installed with Windowns 11 with two nVidia A5000 Ada Generation GPUs. The QWEN 32B was deployed on a Dell R750 (2U) rack server with two nVidia RTX 3090 GPUs.

##### S1.19 Post-processing of model responses

The model responses were further processed, to remove emojis, excessive white spaces and line breaks, to reduce recognizability of model identities.

##### S1.20 Progress monitoring

All of the scores were stored in a JSON file. During the evaluation rounds, every a few hours, the JSON score file was downloaded and saved with a time-stamp. This served as a snapshot of the project, and help monitor progress and screen for ultra-speedy evaluators. In total, 141 snapshots were taken, with analyses shown in supplementary Fig. S4B.

##### S1.21 Fatigue monitoring

Fatigue presents a significant challenge in expert human evaluation, particularly given the clinical responsibilities and time constraints of senior medical specialists. Scoring arbitrariness may emerge as evaluations progress, especially toward later stages. To assess potential fatigue-related bias, we computed, for each evaluator and question, the correlation coefficient between response length and assigned score across all model outputs. An increasing correlation over time was interpreted as suggestive of fatigue-induced variability, raising concerns about score reliability in later portions of the evaluation. One evaluator (X) appeared to display such pattern, and upon communication, X withdrew and results from X were excluded for further analyses.

##### S1.22 Consensus rating of question characteristics

Consensus rating of question characteristics, including relatedness to rare diseases (yes/no), diagnosis (yes/no), treatment planning (yes/no), and operational procedures (yes/no), were determined by simple majority voting.

##### S1.23 Intra-discipline inter-rater consistency

Upon completion of round 2, intra-discipline cross-evaluator consistencies were calculated on a per question basis, whereby the seven scores corresponding to the seven models from one evaluator were compared with another via Pearson's correlation coefficient. Questions with low or even negative correlations ($r$ <0.3) were sent back to the corresponding experts for double check, but score adjustment was not mandatory and evaluators might keep their original score. To monitor the progress, results of the score were saved intermittently.

##### S1.24 Question and answer IDs

Questions of the 13 disciplines were aggregated, assigned random question ID (QIDs), and shuffled to avoid consecutive same-theme questions that may cause the models to learn with time, before submitted to the models via APIs (application programming interface).

To prevent recognition bias, all returned model responses were assigned randomized 6-byte alphanumeric answer IDs (AIDs) post-experiment. These AIDs were used to alphabetically reorder responses, ensuring models appeared in varying positions across questions.

##### S1.25 Linear models and mixed-effects modeling for inferring intrinsic capacities of LLMs

To infer the relations between the strength scores of the model responses and the independent variables or covariates, we fitted a multiple linear regression model:

$$\begin{aligned}\text{strength}_i \;=\; & \beta_1 \cdot \text{model}_i + \beta_2 \cdot \text{rlenrank}_i + \\ & \beta_3 \cdot \text{qtype}_i + \beta_4 \cdot \text{subject}_i + \beta_5 \cdot \text{difficulty}_i + \\ & \beta_6 \cdot \text{rare}_i + \beta_7 \cdot \text{diagnosis}_i + \beta_8 \cdot \text{treatment}_i + \\ & \beta_9 \cdot \text{procedure}_i + \beta_{10} \cdot \text{qlen}_i + \varepsilon_i\end{aligned}$$

where $\text{strength}_i$ is the raw strength for the $i$-th record, model$\in \{\texttt{DS32B}, \texttt{DS70B}, \texttt{DS r1 671B}, \texttt{Qwen32B}, \texttt{Qwen Max}, \texttt{ChatGPT-4o latest}, \texttt{Gemini 2.0 Flash}\}$, qtype$\in \{\texttt{OSCE}, \texttt{real-case}\}$, rlenrank$\in \{\texttt{1}, \texttt{1.5}, \texttt{2}, \texttt{3}, \texttt{3.5}, \texttt{4}, \texttt{4.5}, \texttt{5}, \texttt{6}, \texttt{7}\}$, difficulty$\in \{\texttt{easy}, \texttt{medium}, \texttt{hard}\}$, rare$\in \{\texttt{No}, \texttt{Yes}\}$, diagnosis$\in \{\texttt{No}, \texttt{Yes}\}$, treatment$\in \{\texttt{No}, \texttt{Yes}\}$, procedure$\in \{\texttt{No}, \texttt{Yes}\}$, and $\text{qlen}_i$ is the question length. $\beta_1$ to $\beta_{10}$ are the corresponding coefficients, and $\varepsilon_i$ models the residual.

While the question characteristics and evaluator biases might play a role in score variability, our main interests were how the models, disciplines (subjects), question types (qtype), affect the scores. As such, we modeled the question- and evaluator-specific variability as random effects, and the model-, discipline-, qtype-specific effects as fixed effects, and employed a linear mixed-effects model to fit the scores, expressed as:

$$\begin{aligned}\text{strength}_{ij} \;=\; & \beta_1 \cdot \text{model}_i + \beta_2 \cdot \text{rlenrank}_i + \\ & \beta_3 \cdot \text{subject}_i + \beta_4 \cdot \text{qtype}_i + \beta_5 \cdot \text{difficulty}_i + \\ & \beta_6 \cdot \text{rare}_i + \beta_7 \cdot \text{diagnosis}_i + \beta_8 \cdot \text{treatment}_i + \\ & \beta_9 \cdot \text{procedure}_i + \beta_{10} \cdot \text{qlen}_i + \varepsilon_i + \\ & + u_{0i}^{\text{evaluator}} + u_{0j}^{\text{QID}} + \varepsilon_{ij}\end{aligned}$$

where $u_{0i}^{\text{evaluator}} \sim \mathcal{N}(0, \tau^2_{\text{evaluator}})$ is the random intercept for evaluator $i$; $u_{0j}^{\text{QID}} \sim \mathcal{N}(0, \tau^2_{\text{QID}})$ is the random intercept for question $j$, and $\varepsilon_{ij} \sim \mathcal{N}(0, \sigma^2)$ is the residual error. The model was fitted using restricted maximum likelihood (REML) via the `lme4` package in R (4.3.1).

#### S2 Supplementary Figures

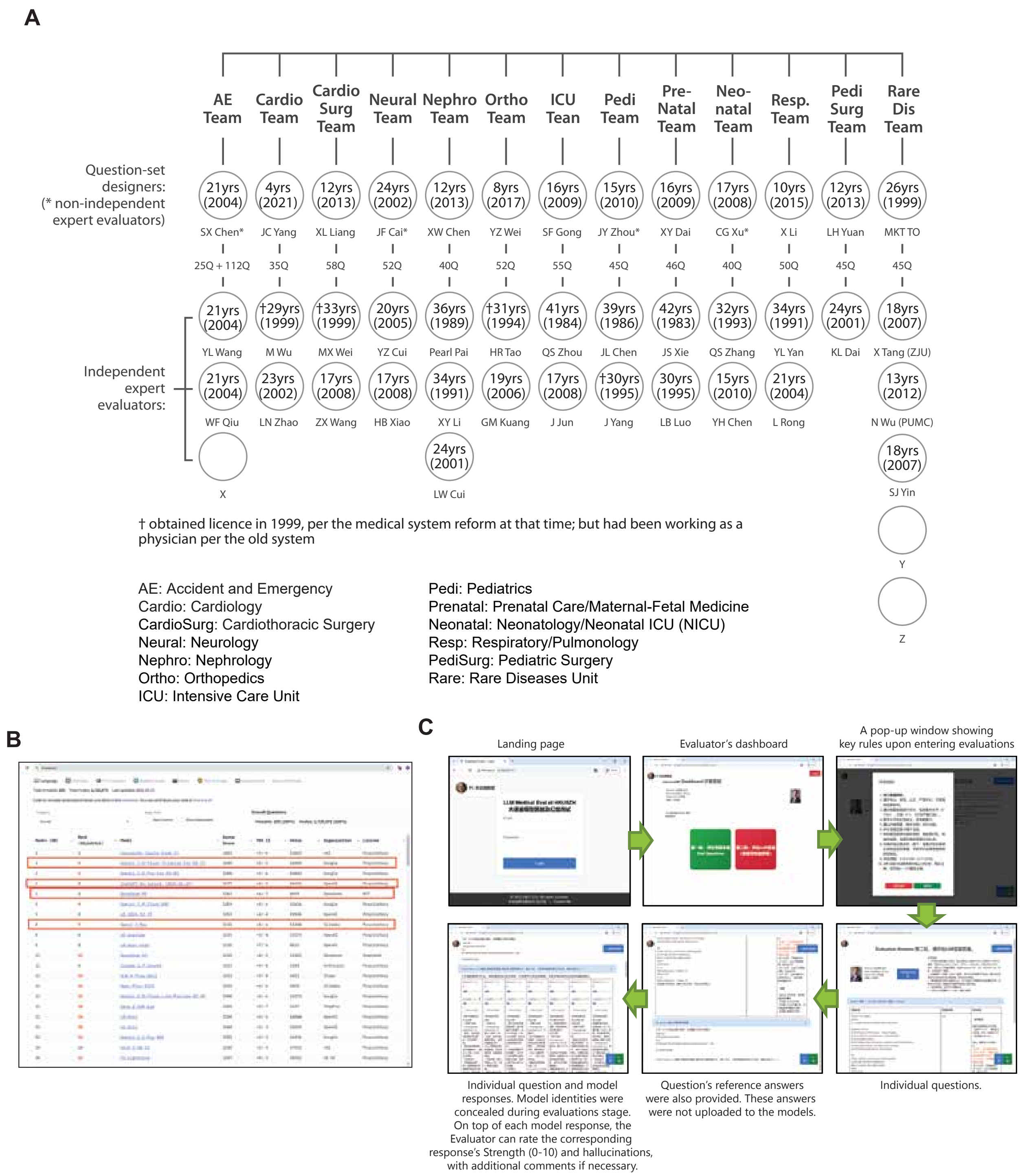

Figure S1: Overall design. (A) 13 teams were assembled, consisting of question designers (one for each subject) and independent expert evaluators (at least two for each subject, except for pediatric surgery). Four of the question designers were also involved in evaluating the answers, but their scores were for reference only. Numbers in circle indicate years in medical practices, with years in brackets relating to year the physician obtain the national medical licensing examination (NMLE) , which was enacted in 1999 [1]. (B) Screenshot showing the SOTA models by the end of February 2025 (Feb 27) on a popular leaderboard ARENA. Four models were selected for their API accessibility. (C) Screenshot of our in-house'ly developed web system for model evaluations.

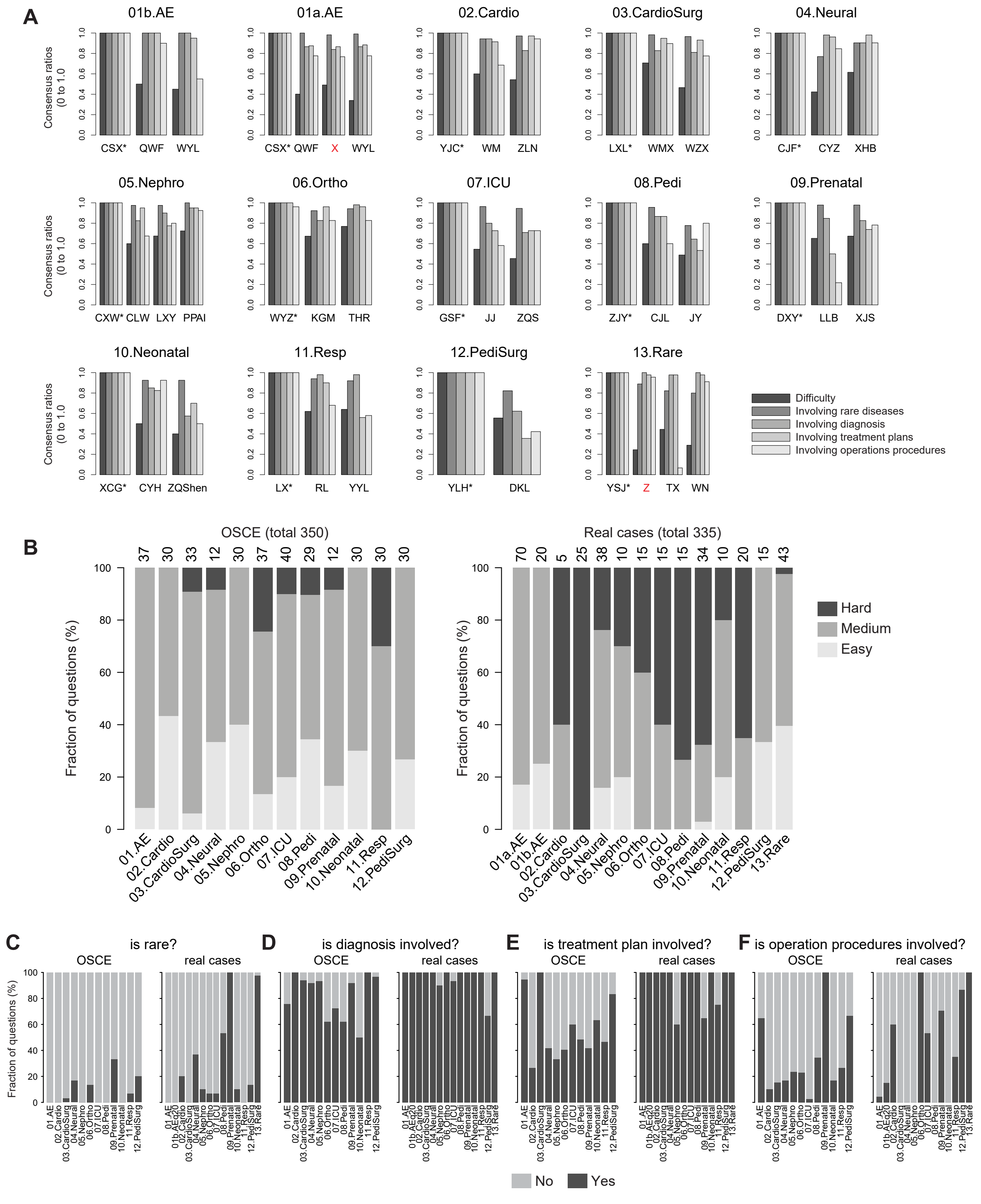

Figure S2: Round 1 evaluation consistency and consensus results. (A) Individual evaluators' conformation with their respective group's consensus assessments on four dimensions of the questions: difficulty, rare disease, diagnosis, treatment, procedures in the first round. Names with * are designers or non-independent evaluators. Their assessments serve as references. (B) Among the two types of questions, bar-charts showing the degree of difficulty distribution per subject/team. Numbers on top indicate total number of questions per subject. C-F, Among the two types of questions, bar-charts showing the distributions of rare disease (C), diagnosis (D), treatment plan (E) and operational procedures (F) involved, per subject/team.

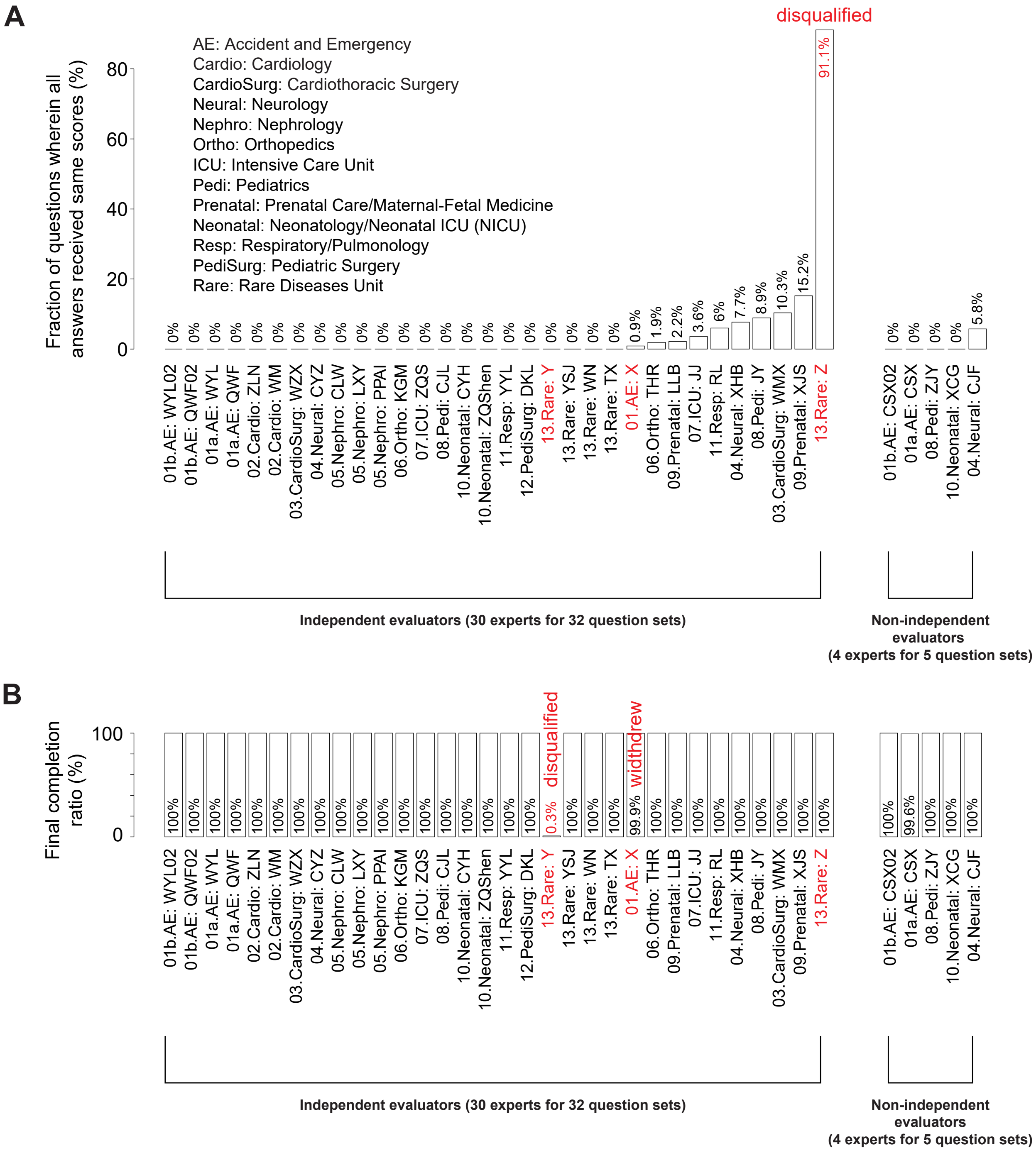

Figure S3: Quality assessments in round 2. (A) Bar chart illustrating the proportion of questions assigned identical scores by each evaluator. evaluators exhibiting an excessive rate of identical scores (>20%) were deemed non-informative and excluded from the analysis. (B) Bar chart displaying the completion rate of evaluations per evaluator. evaluators who failed to assess a sufficient fraction of the questions were considered non-informative and subsequently disqualified.

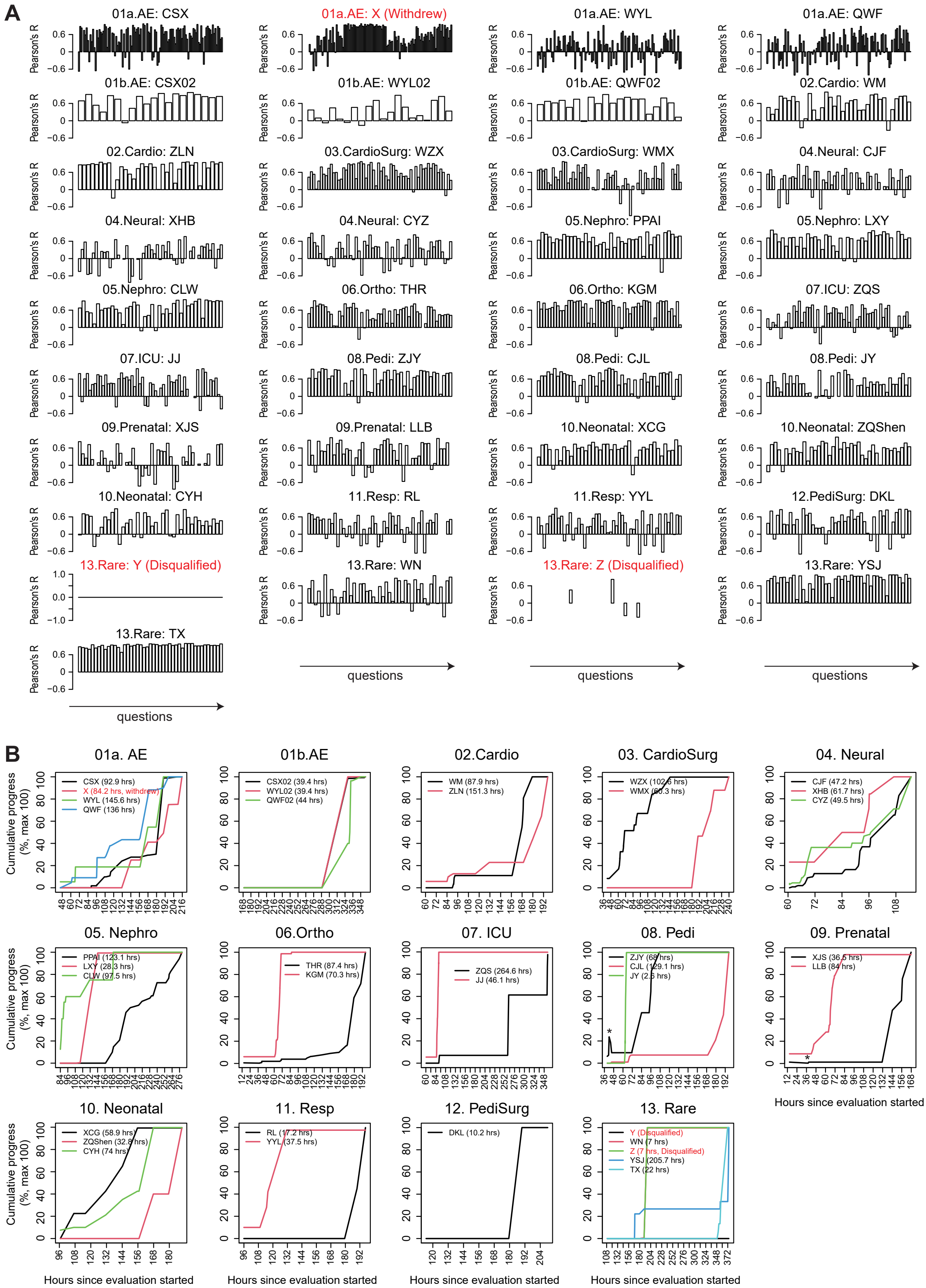

Figure S4: Fatigue and progress monitoring for Round 2 evaluation. (A) Bar plots depicting the correlation between each evaluator's scores (both independent and non-independent) and model response lengths (measured using Pearson's $r$) across increasing question order. The observed upward trend in correlation along the sequence of questions (as they appeared on the web platform) may suggest the potential influence of evaluator fatigue, leading to a decline in the quality of ratings over time. (B) Cumulative progress curves for each individual evaluator, showing their evaluation progress since the start of the second-round evaluation on March 10 at 8:00 PM local time. The legend identifies evaluators by their initials, and the numbers in parentheses represent the estimated durations required to achieve 5% to 95% of total progress. C. Instances of technical issues are indicated by black asterisks (*), where evaluators reported losing their scores after logging out. These disruptions were likely caused by a restart of the web system during their scoring sessions, leading to drops in their respective progress curves.

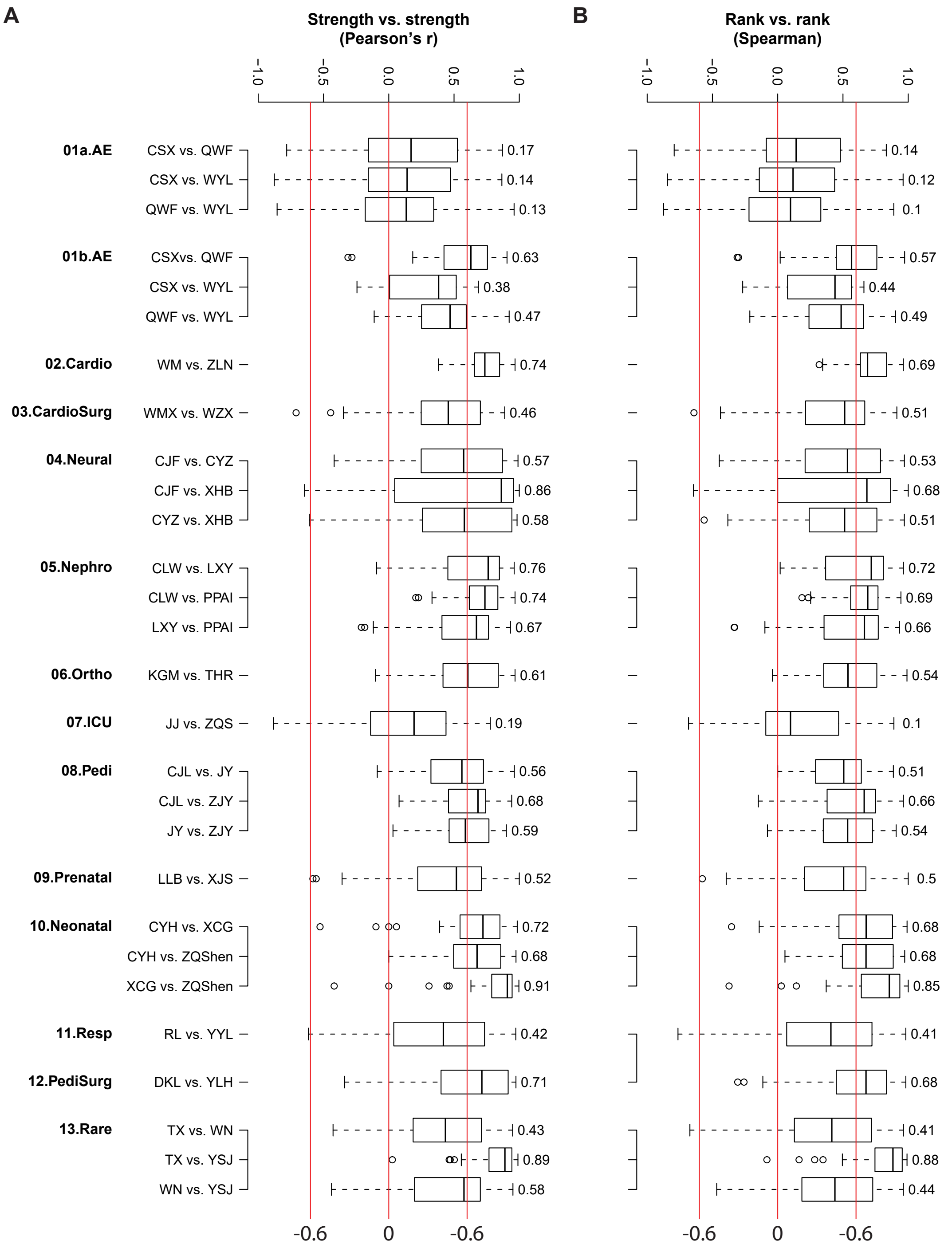

Figure S5: Round 2 pairwise consistency assessment among intra-subject evaluators. (A) Box-and-whisker plots depicting the distribution of intra-discipline consistency across evaluator pairs, with each plot representing the distribution of scores between two evaluators within the same discipline. (B) Same as (A), but showing Spearman's correlation coefficients.

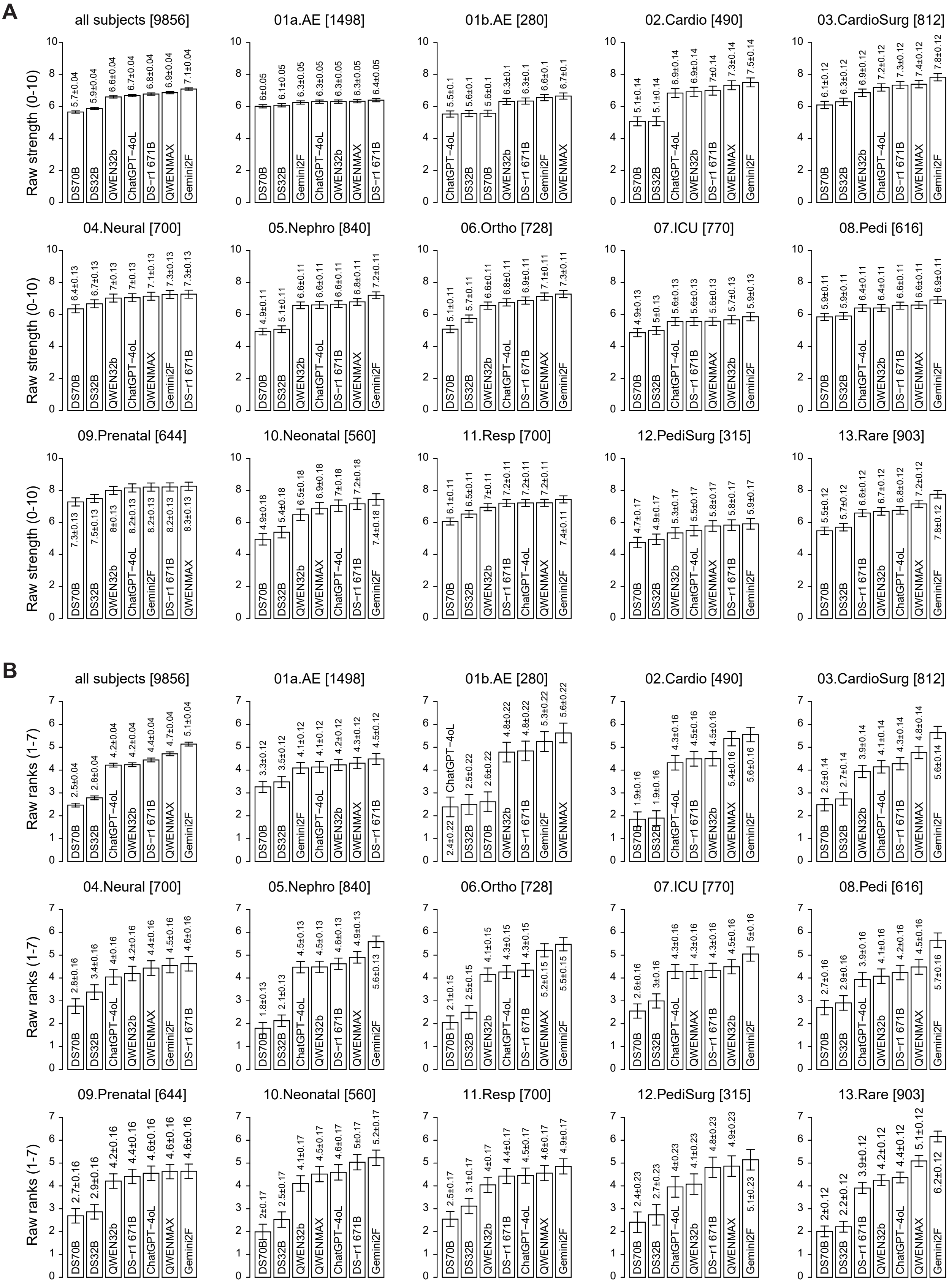

Figure S6: Raw scores on strengths and corresponding raw ranks, by the independent evaluators (OSCE and real-case questions combined) (A) Bar plots showing the raw scores for all subjects combined (topleft panel) or individually (other panels). The numbers in square brackets indicate numbers of records, which are the products of the numbers of qualified questions, models and independent evaluators. (B) Similar arrangements as in (A), except the raw ranks were shown. The raw ranks (range from 1 to 7) are based on the ranking of the seven responses for each question and by each evaluator. Ties were handled with the "average" method.

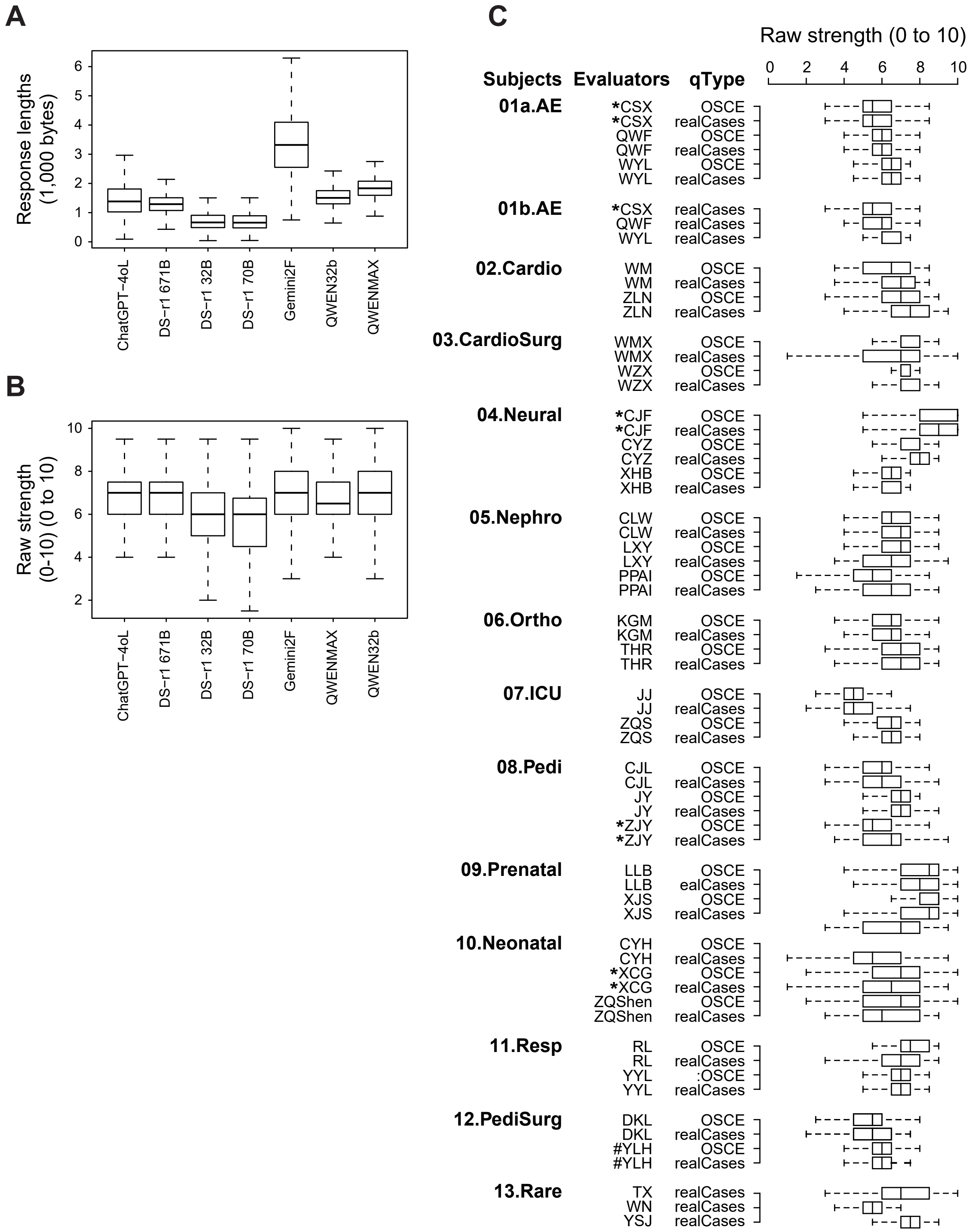

Figure S7: Marked baseline variations in round 2 evaluation metrics. (A) Bar plots showing the distribution of response lengths across models. (B) Box-and-whisker plots summarizing strength scores assigned to each model in Round 2. (C) Box-and-whisker plots illustrating score variability across individual evaluators, revealing inter-evaluator differences in scoring tendencies. Initials with asterisks (*) denote non-independent expert evaluators; initials with hash marks (#) indicate question designers providing reference scores, excluded from primary analyses.

#### S3 Supplementary Tables

Table S1: Questions. OSCE: Objective Structured Clinical Examination. realCases: real medical cases selected from HKU-SZH. Numbers in brackets indicate disqualified questions (total 10).

(A)

| Subjects/Teams | Team Abbreviations | Number of OSCE questions | Number of real-case questions | Number of independent evaluators | Number of non-independent evaluators |
|---|---|---|---|---|---|
| AE first batch | [01a.AE] | 37 (+3) | 70 (+2) | 2 | 1 |
| AE second batch | [01b.AE] | 0 | 20 | 2 | 1 |
| Cardiology | [02.Cardio] | 30 | 5 | 2 | 0 |
| Cardiac surgery | [03.CardiacSurg] | 33 | 25 | 2 | 0 |
| Neurology | [04.Neural] | 12 | 38 (+2) | 2 | 1 |
| Nephrology | [05.Nephro] | 30 | 10 | 3 | 0 |
| Orthopedics | [06.Ortho] | 37 | 15 | 2 | 0 |
| ICU | [07.ICU] | 40 | 15 | 2 | 0 |
| Pediatrics | [08.Pedi] | 29 (+1) | 15 | 2 | 1 |
| Prenatal | [09.Prenatal] | 12 | 34 | 2 | 0 |
| Neonatal | [10.Neonatal] | 30 | 10 | 2 | 1 |
| Respiratory | [11.Resp] | 30 | 20 | 2 | 0 |
| Pediatric Surgery | [12.PediSurg] | 30 | 15 | 1 | 0 |
| Rare Diseases | [13.Rare] | 0 | 43 (+2) | 3 | 0 |
| col-median | | 30.0 | 17.5 | | |
| col-average | | 25.0 | 23.9 | | |
| col-sum | | 350 (+4) | 335 (+6) | | |

(B)

| | qtype | Median question length (characters) | IQR |
|---|---|---|---|
| 01a.AE | OSCE | 357 | 271 – 441 |
| | realCases | 1493.5 | 1370.25 – 1717.8 |
| 01b.AE | realCases | 684.5 | 303.75 – 782 |
| 02.Cardio | OSCE | 199.5 | 136.75 – 291 |
| | realCases | 1089 | 672 – 1142 |
| 03.CardiacSurg | OSCE | 52 | 46 – 86 |
| | realCases | 3003 | 2492 – 3726 |
| 04.Neural | OSCE | 110 | 86.75 – 127.5 |
| | realCases | 368.5 | 299.25 – 418 |
| 05.Nephro | OSCE | 706.5 | 469.25 – 1004.8 |
| | realCases | 304.5 | 185 – 383.8 |
| 06.Ortho | OSCE | 223 | 119 – 1150 |
| | realCases | 971 | 911.5 – 1297.5 |
| 07.ICU | OSCE | 99 | 44.5 – 148.8 |
| | realCases | 488 | 313 – 1274 |
| 08.Pedi | OSCE | 249 | 131 – 523 |
| | realCases | 627 | 554.5 – 829.5 |
| 09.Prenatal | OSCE | 278 | 140 – 761 |
| | realCases | 114 | 71.25 – 200.8 |
| 10.Neonatal | OSCE | 206.5 | 108.25 – 332.5 |
| | realCases | 722 | 494.25 – 882.5 |
| 11.Resp | OSCE | 368 | 325.25 – 472.3 |
| | realCases | 597.5 | 534.5 – 842.3 |
| 12.PediSurg | OSCE | 65 | 61 – 68 |
| | realCases | 55 | 53 – 93 |
| 13.Rare | realCases | 613 | 430 – 977 |

Table S2: Regression estimates and statistical significance for model response length (rlen, in 1,000 characters) as a function of question length (qlen, in 1,000 characters), question type (qtype), language model (model, with "DS32B" as the reference category), and discipline (subject, with "01a.AE" as the reference category). The dataset comprises 4,795 observations, corresponding to 7 responses by 7 models per each of the 685 questions. Significant values ($p < 0.05$) are in bold.

| | rlen (response-lengths, in 1,000 characters) | | | |
|---|---|---|---|---|
| Predictors | Estimates | Standard Error | 95% C.I. | *p* |
| **Models** | | | | |
| model [DS70B] | 0.40 | 0.10 | 0.21, 0.59 | <**0.001** |
| model [DS32B] | 0.41 | 0.10 | 0.22, 0.60 | <**0.001** |
| model [DS-r1 671B] | 1.00 | 0.10 | 0.81, 1.18 | <**0.001** |
| model [ChatGPT-4oL] | 1.12 | 0.10 | 0.93, 1.31 | <**0.001** |
| model [QWEN32b] | 1.33 | 0.10 | 1.14, 1.52 | <**0.001** |
| model [QWENMAX] | 1.52 | 0.10 | 1.33, 1.71 | <**0.001** |
| model [Gemini2F] | 3.64 | 0.10 | 3.45, 3.83 | <**0.001** |
| **Disciplines** | | | | |
| subject [01b.AE] | -0.13 | 0.16 | -0.44, 0.18 | 0.407 |
| subject [02.Cardio] | -0.07 | 0.13 | -0.32, 0.18 | 0.580 |
| subject [03.CardiacSurg] | 0.13 | 0.10 | -0.08, 0.33 | 0.226 |
| subject [04.Neural] | -0.12 | 0.12 | -0.35, 0.10 | 0.291 |
| subject [05.Nephro] | 0.08 | 0.12 | -0.15, 0.31 | 0.486 |
| subject [06.Ortho] | 0.08 | 0.11 | -0.13, 0.29 | 0.460 |
| subject [07.ICU] | 0.36 | 0.11 | 0.14, 0.57 | <**0.001** |
| subject [08.Pedi] | 0.02 | 0.11 | -0.20, 0.25 | 0.860 |
| subject [09.Prenatal] | -0.01 | 0.12 | -0.25, 0.22 | 0.909 |
| subject [10.Neonatal] | 0.21 | 0.12 | -0.03, 0.44 | 0.083 |
| subject [11.Resp] | 0.51 | 0.11 | 0.30, 0.73 | <**0.001** |
| subject [12.PediSurg] | -0.12 | 0.12 | -0.35, 0.12 | 0.327 |
| subject [13.Rare] | -0.23 | 0.12 | -0.46, 0.00 | 0.053 |
| **Question characteristics** | | | | |
| qlen (per 1,000 characters) | 0.12 | 0.05 | 0.03, 0.21 | **0.009** |
| qtype [realCases] | 0.35 | 0.06 | 0.22, 0.47 | <**0.001** |
| Observations | 4,795 | | | |
| $R^2$ / $R^2$ adjusted | 0.587 / 0.585 | | | |

Table S3: Regression Results for Strengths and Strength ranks, with respect to response lengths (rlen), as real-valued (A), ranking of rlen (rlenrank) as integers (B), and ranking of rlen as categorical (C) variables, respectively.

(A) response lengths (rlen, per 1,000 characters) as a real-valued, independent variable.

| | Raw strengths (0-10) | | | | Strength ranks (1-7) | | | |
|---|---|---|---|---|---|---|---|---|
| Predictors | Estimates | Std. Error | 95% C.I. | $p$ | Estimates | Std. Error | 95% C.I. | $p$ |
| (Intercept) | 6.32 | 0.02 | 6.29, 6.36 | <**0.001** | 3.66 | 0.02 | 3.61, 3.70 | <**0.001** |
| rlen | 0.11 | 0.01 | 0.10, 0.13 | <**0.001** | 0.21 | 0.01 | 0.19, 0.23 | <**0.001** |
| Observations | 9,856 | | | | 9,856 | | | |
| $R^2$ / $R^2$ adjusted | 0.024 / 0.024 | | | | 0.047 / 0.047 | | | |

(B) ranking of rlen (rlenrank) per each answer as an integer-valued, independent variable.

| | Raw strengths (0-10) | | | | Strength ranks (1-7) | | | |
|---|---|---|---|---|---|---|---|---|
| Predictors | Estimates | Std. Error | 95%C.I. | $p$ | Estimates | Std. Error | 95%C.I. | $p$ |
| (Intercept) | 5.60 | 0.03 | 5.54, 5.66 | <**0.001** | 2.25 | 0.04 | 2.17, 2.32 | <**0.001** |
| rlenrank | 0.23 | 0.01 | 0.21 – 0.24 | <**0.001** | 0.44 | 0.01 | 0.42, 0.45 | <**0.001** |
| Observations | 9,856 | | | | 9,856 | | | |
| $R^2$ / $R^2$ adjusted | 0.102 / 0.102 | | | | 0.224 / 0.224 | | | |

(C) ranking of rlen as categorical, independent variables.

| | Raw strengths (0-10) | | | | Strength ranks (1-7) | | | |
|---|---|---|---|---|---|---|---|---|
| Predictors | Estimates | Std. Error | 95% C.I. | $p$ | Estimates | Std. Error | 95% C.I. | $p$ |
| (Intercept) | 5.67 | 0.04 | 5.60, 5.74 | <**0.001** | 2.42 | 0.04 | 2.33, 2.50 | <**0.001** |
| rlenrank [1.5] | 0.37 | 0.39 | -0.39, 1.14 | 0.339 | 0.54 | 0.47 | -0.38, 1.45 | 0.248 |
| rlenrank [2] | 0.25 | 0.05 | 0.15, 0.35 | <**0.001** | 0.45 | 0.06 | 0.34, 0.57 | <**0.001** |
| rlenrank [3] | 0.91 | 0.05 | 0.81, 1.01 | <**0.001** | 1.64 | 0.06 | 1.52, 1.75 | <**0.001** |
| rlenrank [3.5] | 1.58 | 0.67 | 0.26, 2.90 | **0.019** | 2.58 | 0.81 | 1.00, 4.16 | **0.001** |
| rlenrank [4] | 1.04 | 0.05 | 0.94, 1.13 | <**0.001** | 1.91 | 0.06 | 1.79, 2.03 | <**0.001** |
| rlenrank [4.5] | 0.58 | 0.67 | -0.74, 1.90 | 0.388 | 2.58 | 0.81 | 1.00, 4.16 | **0.001** |
| rlenrank [5] | 1.08 | 0.05 | 0.98, 1.18 | <**0.001** | 2.02 | 0.06 | 1.91, 2.14 | <**0.001** |
| rlenrank [6] | 1.19 | 0.05 | 1.09, 1.29 | <**0.001** | 2.30 | 0.06 | 2.18, 2.41 | <**0.001** |
| rlenrank [7] | 1.45 | 0.05 | 1.35, 1.55 | <**0.001** | 2.74 | 0.06 | 2.62, 2.86 | <**0.001** |
| Observations | 9,856 | | | | 9,856 | | | |
| $R^2$ / $R^2$ adjusted | 0.115 / 0.115 | | | | 0.247 / 0.246 | | | |

Table S4: Summary of multiple linear regression results for evaluation raw scores (ranging from 0 to 10, with higher scores indicating better performance) in relation to a set of covariates, including language model (model), discipline (subject, with "01a.AE" as the reference category), question type (qtype, with "OSCE" as the reference), question difficulty (difficulty, with "easy" as the reference category), and factors related to rare diseases, diagnosis, treatment, and operational procedures, across all independent evaluators, with "no" as the reference category for the last four covariates. Model 1a includes 9,856 records (setA) from 27 independent qualified evaluators. Model 1b was fitted with the same covariates but on setABC, which included these 9,856 records, 1,824 records from 4 non-independent evaluators, and 315 records from question designer for the pediatric surgery team (12.PediSurg), where there is only one evaluator.

| | Model 1a: Strength scores | | | Model 1b: Strength scores | | |
|---|---|---|---|---|---|---|
| Predictors | Estimates | 95% C.I. | $p$ | Estimates | 95% C.l. | $p$ |
| **Models** | | | | | | |
| model [DS70B] | 5.41 | 5.27, 5.55 | <**0.001** | 5.22 | 5.09, 5.35 | <**0.001** |
| model [DS32B] | 5.63 | 5.49, 5.77 | <**0.001** | 5.45 | 5.32, 5.58 | <**0.001** |
| model [QWEN32b] | 6.35 | 6.21, 6.49 | <**0.001** | 6.16 | 6.03, 6.29 | <**0.001** |
| model [ChatGPT-4oL] | 6.43 | 6.29, 6.57 | <**0.001** | 6.24 | 6.11, 6.37 | <**0.001** |
| model [DS-r1 671B] | 6.53 | 6.39, 6.67 | <**0.001** | 6.35 | 6.23, 6.48 | <**0.001** |
| model [QWENMAX] | 6.61 | 6.47, 6.76 | <**0.001** | 6.42 | 6.29, 6.55 | <**0.001** |
| model [Gemini2F] | 6.84 | 6.70, 6.98 | <**0.001** | 6.76 | 6.63, 6.89 | <**0.001** |
| **Disciplines** | | | | | | |
| subject [01b.AE] | -0.18 | -0.34, -0.02 | **0.030** | -0.18 | -0.32, -0.04 | **0.012** |
| subject [02.Cardio] | 0.30 | 0.16, 0.43 | <**0.001** | 0.43 | 0.30, 0.57 | <**0.001** |
| subject [03.CardiacSurg] | 0.82 | 0.70, 0.93 | <**0.001** | 0.95 | 0.84, 1.06 | <**0.001** |
| subject [04.Neural] | 0.65 | 0.53, 0.78 | <**0.001** | 1.29 | 1.18, 1.39 | <**0.001** |
| subject [05.Nephro] | 0.03 | -0.08, 0.14 | 0.577 | 0.17 | 0.06, 0.28 | **0.002** |
| subject [06.Ortho] | 0.21 | 0.09, 0.32 | **0.001** | 0.36 | 0.24, 0.48 | <**0.001** |
| subject [07.ICU] | -0.84 | -0.96, -0.72 | <**0.001** | -0.69 | -0.80, -0.57 | <**0.001** |
| subject [08.Pedi] | 0.09 | -0.03, 0.22 | 0.150 | 0.08 | -0.03, 0.19 | 0.155 |
| subject [09.Prenatal] | 1.53 | 1.38, 1.69 | <**0.001** | 1.68 | 1.53, 1.83 | <**0.001** |
| subject [10.Neonatal] | 0.19 | 0.06, 0.31 | **0.004** | 0.39 | 0.28, 0.50 | <**0.001** |
| subject [11.Resp] | 0.70 | 0.57, 0.82 | <**0.001** | 0.83 | 0.71, 0.95 | <**0.001** |
| subject [12.PediSurg] | -0.93 | -1.09, -0.77 | <**0.001** | -0.47 | -0.59, -0.34 | <**0.001** |
| subject [13.Rare] | 0.14 | -0.02, 0.30 | 0.080 | 0.32 | 0.17, 0.47 | <**0.001** |
| **Question characteristics** | | | | | | |
| qtype [realCases] | -0.01 | -0.08, 0.07 | 0.869 | -0.03 | -0.10, 0.04 | 0.457 |
| difficulty [medium] | 0.09 | 0.02, 0.16 | **0.009** | 0.09 | 0.02, 0.15 | **0.007** |
| difficulty [hard] | 0.00 | -0.10, 0.10 | 0.988 | 0.03 | -0.07, 0.13 | 0.542 |
| rare [yes] | 0.17 | 0.06, 0.28 | **0.002** | 0.18 | 0.08, 0.28 | <**0.001** |
| diagnosis [yes] | -0.07 | -0.16, 0.02 | 0.126 | -0.02 | -0.10, 0.06 | 0.601 |
| treatment [yes] | 0.05 | -0.03, 0.12 | 0.227 | 0.02 | -0.05, 0.09 | 0.551 |
| procedure [yes] | 0.05 | -0.02, 0.12 | 0.126 | 0.03 | -0.04, 0.09 | 0.429 |
| qlen | -0.07 | -0.11, -0.02 | **0.007** | -0.06 | -0.10, -0.01 | **0.019** |
| Observations | | 9,856 | | | 11,995 | |
| $R^2$ / $R^2$ adjusted | | 0.967 / 0.967 | | | 0.964 / 0.964 | |

Table S5: (A) Results of multiple linear regression models predicting evaluator-assigned raw scores (range: 0–10, higher indicating better performance) for medical strength and quality across all independent evaluators, subjects, and question types. The variable rlenrank represents model response length rankings (1 to 7, with higher values indicating longer responses); ranks 1.5, 3.5, and 4.5 reflect tied positions. The covariate uses as the reference level. References: DS32B, 01a.AE, easy for model, subject, covariates, and difficulty, respectively. Rare disease involvement, diagnostic content, treatment planning, and procedural content all have (no) as the reference . Results are reported as point estimates with 95% confidence intervals (C.I.) and associated p-values. Model 2a and Model 2b were fitted on setA and setABC, respectively. (B) ANOVA comparison of Models. ANOVA, Analysis of Variance; AIC: Akaike Information Criterion; RSS: Residual sum of squares; DF: Degree of freedom.

(A)

| | Model 2a: Strength scores | | | Model 2b: Strength scores | | |
|---|---|---|---|---|---|---|
| Predictors | Estimates | 95% C.I. | *p* | Estimates | 95% C.I. | *p* |
| **Models** | | | | | | |
| model [DS70B] | 5.29 | 5.14, 5.44 | **<0.001** | 5.10 | 4.97, 5.24 | **<0.001** |
| model [DS32B] | 5.52 | 5.37, 5.67 | **<0.001** | 5.34 | 5.20, 5.47 | **<0.001** |
| model [QWEN32b] | 5.78 | 5.58, 5.97 | **<0.001** | 5.57 | 5.39, 5.76 | **<0.001** |
| model [ChatGPT-4oL] | 5.93 | 5.74, 6.12 | **<0.001** | 5.72 | 5.55, 5.90 | **<0.001** |
| model [Gemini2F] | 5.94 | 5.69, 6.19 | **<0.001** | 5.80 | 5.56, 6.05 | **<0.001** |
| model [QWENMAX] | 5.96 | 5.76, 6.17 | **<0.001** | 5.76 | 5.57, 5.94 | **<0.001** |
| model [DS-r1 671B] | 6.06 | 5.88, 6.25 | **<0.001** | 5.87 | 5.69, 6.04 | **<0.001** |
| rlenrank [1.5] | 0.61 | -0.07, 1.30 | 0.079 | 0.24 | -0.38, 0.86 | 0.453 |
| rlenrank [2] | 0.17 | 0.08, 0.26 | **<0.001** | 0.17 | 0.09, 0.26 | **<0.001** |
| rlenrank [3] | 0.41 | 0.27, 0.54 | **<0.001** | 0.43 | 0.30, 0.56 | **<0.001** |
| rlenrank [3.5] | 0.68 | -0.51, 1.87 | 0.264 | 1.12 | 0.10, 2.14 | 0.032 |
| rlenrank [4] | 0.53 | 0.39, 0.67 | **<0.001** | 0.55 | 0.41, 0.68 | **<0.001** |
| rlenrank [4.5] | 1.16 | -0.03, 2.35 | 0.056 | 1.16 | -0.09, 2.41 | 0.068 |
| rlenrank [5] | 0.61 | 0.46, 0.75 | **<0.001** | 0.60 | 0.46, 0.75 | **<0.001** |
| rlenrank [6] | 0.67 | 0.51, 0.83 | **<0.001** | 0.70 | 0.55, 0.84 | **<0.001** |
| rlenrank [7] | 0.92 | 0.71, 1.14 | **<0.001** | 0.97 | 0.76, 1.19 | **<0.001** |
| **Disciplines** | | | | | | |
| subject [01b.AEq20] | -0.18 | -0.34, -0.02 | 0.030 | -0.18 | -0.32, -0.04 | 0.012 |
| subject [02.Cardio] | 0.30 | 0.16, 0.43 | **<0.001** | 0.43 | 0.30, 0.57 | **<0.001** |
| subject [03.CardiacSurg] | 0.82 | 0.70, 0.93 | **<0.001** | 0.95 | 0.84, 1.06 | **<0.001** |
| subject [04.Neural] | 0.65 | 0.53, 0.77 | **<0.001** | 1.28 | 1.18, 1.39 | **<0.001** |
| subject [05.Nephro] | 0.03 | -0.08, 0.15 | 0.551 | 0.17 | 0.06, 0.28 | 0.002 |
| subject [06.Ortho] | 0.21 | 0.09, 0.33 | **<0.001** | 0.36 | 0.25, 0.48 | **<0.001** |
| subject [07.ICU] | -0.85 | -0.96, -0.73 | **<0.001** | -0.69 | -0.81, -0.58 | **<0.001** |
| subject [08.Pedi] | 0.09 | -0.03, 0.22 | 0.139 | 0.08 | -0.03, 0.19 | 0.147 |
| subject [09.Prenatal] | 1.54 | 1.38, 1.69 | **<0.001** | 1.68 | 1.53, 1.83 | **<0.001** |
| subject [10.Neonatal] | 0.19 | 0.06, 0.31 | **0.003** | 0.39 | 0.28, 0.50 | **<0.001** |
| subject [11.Resp] | 0.70 | 0.57, 0.82 | **<0.001** | 0.83 | 0.71, 0.96 | **<0.001** |
| subject [12.PediSurg] | -0.93 | -1.08, -0.77 | **<0.001** | -0.47 | -0.59, -0.34 | **<0.001** |
| subject [13.Rare] | 0.15 | -0.01, 0.31 | 0.068 | 0.32 | 0.17, 0.47 | **<0.001** |
| **Question characteristics** | | | | | | |
| qtype [realCases] | -0.01 | -0.08, 0.07 | 0.870 | -0.03 | -0.10, 0.04 | 0.473 |
| difficulty [medium] | 0.09 | 0.02, 0.16 | **0.009** | 0.09 | 0.02, 0.15 | **0.007** |
| difficulty [hard] | 0.00 | -0.10, 0.10 | 0.991 | 0.03 | -0.07, 0.13 | 0.529 |
| rare [yes] | 0.17 | 0.06, 0.27 | **0.003** | 0.18 | 0.08, 0.28 | **<0.001** |
| diagnosis [yes] | -0.07 | -0.16, 0.02 | 0.132 | -0.02 | -0.10, 0.06 | 0.590 |
| treatment [yes] | 0.05 | -0.03, 0.12 | 0.203 | 0.02 | -0.05, 0.10 | 0.500 |
| procedure [yes] | 0.05 | -0.02, 0.12 | 0.146 | 0.03 | -0.04, 0.09 | 0.447 |
| qlen | -0.07 | -0.11, -0.02 | **0.006** | -0.06 | -0.10, -0.01 | **0.018** |
| Observations | | 9,856 | | | 11,995 | |
| $R^2$ / $R^2$ adjusted | | 0.968 / 0.967 | | | 0.964 / 0.964 | |

(B) ANOVA and AIC model comparison between Model 1a and Model 2a.

| Models | Residual DF | RSS | DF | Sum of Sq | F-value | *p*-value | AIC |
|---|---|---|---|---|---|---|---|
| Model 1a | 9,829 | 14,343 | - | - | - | - | 31724.18 |
| Model 2a | 9,820 | 14,208 | 9 | 135.84 | 10.433 | $\mathbf{3.25 \times 10^{-16}}$ | 31648.39 |

Table S6: Multiple linear regression results for evaluator-assigned per-question rank scores (1–7; higher = better) across all questions, evaluators, and subjects. rlenrank denotes model response length ranks (1–7; higher = longer), with ties at 1.5, 3.5, and 4.5. The reference levels are: subject (01a.AE), qtype (OSCE), question difficulty (easy), and absence of rare disease, diagnosis, treatment, and procedure content (no for each). Estimates are shown with 95% confidence intervals and p-values.

| | Per-question ascending rank of raw scores | | | | | |
|---|---|---|---|---|---|---|
| | Model 3a: on SetA | | | Model 3b: on SetABC | | |
| Predictors | Estimates | 95% C.I. | *p* | Estimates | 95% C.I. | *p* |
| **Models** | | | | | | |
| model [DS70B] | 2.24 | 2.05, 2.43 | **<0.001** | 2.25 | 2.08, 2.42 | **<0.001** |
| model [DS32B] | 2.57 | 2.38, 2.77 | **<0.001** | 2.58 | 2.41, 2.75 | **<0.001** |
| model [QWEN32b] | 3.06 | 2.80, 3.32 | **<0.001** | 3.04 | 2.82, 3.27 | **<0.001** |
| model [ChatGPT-4oL] | 3.18 | 2.93, 3.43 | **<0.001** | 3.15 | 2.93, 3.38 | **<0.001** |
| model [QWENMAX] | 3.35 | 3.08, 3.62 | **<0.001** | 3.32 | 3.08, 3.56 | **<0.001** |
| model [Gemini2F] | 3.35 | 3.02, 3.68 | **<0.001** | 3.43 | 3.13, 3.74 | **<0.001** |
| model [DS-r1 671B] | 3.49 | 3.24, 3.74 | **<0.001** | 3.49 | 3.27, 3.71 | **<0.001** |
| rlenrank [1.5] | 0.55 | -0.36, 1.46 | 0.233 | 0.18 | -0.61, 0.97 | 0.661 |
| rlenrank [2] | 0.33 | 0.21, 0.45 | **<0.001** | 0.34 | 0.23, 0.45 | **<0.001** |
| rlenrank [3] | 0.82 | 0.64, 1.01 | **<0.001** | 0.80 | 0.64, 0.97 | **<0.001** |
| rlenrank [3.5] | 1.67 | 0.09, 3.25 | **0.039** | 1.68 | 0.38, 2.97 | **0.011** |
| rlenrank [4] | 1.08 | 0.89, 1.27 | **<0.001** | 1.06 | 0.89, 1.24 | **<0.001** |
| rlenrank [4.5] | 1.73 | 0.15, 3.31 | **0.032** | 1.73 | 0.15, 3.31 | **0.032** |
| rlenrank [5] | 1.24 | 1.04, 1.43 | **<0.001** | 1.20 | 1.03, 1.38 | **<0.001** |
| rlenrank [6] | 1.43 | 1.22, 1.64 | **<0.001** | 1.42 | 1.23, 1.61 | **<0.001** |
| rlenrank [7] | 1.82 | 1.54, 2.11 | **<0.001** | 1.85 | 1.58, 2.12 | **<0.001** |
| **Disciplines** | | | | | | |
| subject [01b.AE] | -0.00 | -0.21, 0.21 | 1.000 | 0.00 | -0.17, 0.18 | 0.987 |
| subject [02.Cardio] | 0.00 | -0.18, 0.18 | 0.978 | 0.01 | -0.17, 0.18 | 0.950 |
| subject [03.CardiacSurg] | 0.00 | -0.15, 0.15 | 0.990 | 0.00 | -0.14, 0.15 | 0.950 |
| subject [04.Neural] | -0.01 | -0.17, 0.16 | 0.943 | -0.00 | -0.14, 0.13 | 0.974 |
| subject [05.Nephro] | 0.00 | -0.14, 0.15 | 0.972 | 0.01 | -0.13, 0.14 | 0.936 |
| subject [06.Ortho] | 0.00 | -0.15, 0.16 | 0.972 | 0.01 | -0.14, 0.15 | 0.943 |
| subject [07.ICU] | -0.00 | -0.16, 0.15 | 0.963 | 0.00 | -0.14, 0.15 | 0.987 |
| subject [08.Pedi] | 0.00 | -0.16, 0.17 | 0.981 | 0.00 | -0.13, 0.14 | 0.948 |
| subject [09.Prenatal] | 0.00 | -0.20, 0.21 | 0.973 | 0.00 | -0.19, 0.19 | 0.976 |
| subject [10.Neonatal] | 0.00 | -0.16, 0.17 | 0.987 | 0.00 | -0.13, 0.14 | 0.949 |
| subject [11.Resp] | 0.00 | -0.16, 0.17 | 0.981 | 0.00 | -0.15, 0.16 | 0.954 |
| subject [12.PediSurg] | 0.00 | -0.21, 0.21 | 0.984 | 0.00 | -0.15, 0.16 | 0.966 |
| subject [13.Rare] | 0.00 | -0.21, 0.22 | 0.976 | 0.00 | -0.19, 0.19 | 0.987 |
| **Question characteristics** | | | | | | |
| qtype [realCases] | 0.00 | -0.10, 0.10 | 0.986 | 0.00 | -0.09, 0.09 | 0.944 |
| difficulty [medium] | 0.00 | -0.09, 0.09 | 0.991 | 0.00 | -0.08, 0.08 | 0.991 |
| difficulty [hard] | 0.00 | -0.13, 0.13 | 0.995 | -0.00 | -0.12, 0.12 | 0.998 |
| rare [yes] | -0.00 | -0.14, 0.14 | 0.987 | 0.00 | -0.13, 0.13 | 0.992 |
| diagnosis [yes] | 0.00 | -0.12, 0.12 | 0.996 | 0.00 | -0.10, 0.10 | 0.991 |
| treatment [yes] | 0.00 | -0.09, 0.10 | 0.952 | 0.00 | -0.09, 0.09 | 0.956 |
| procedure [yes] | -0.00 | -0.09, 0.09 | 0.960 | -0.00 | -0.08, 0.08 | 0.992 |
| qlen | -0.00 | -0.06, 0.06 | 0.979 | -0.00 | -0.06, 0.06 | 0.965 |
| Observations | | 9,856 | | | 11,995 | |
| $R^2$ / $R^2$ adjusted | | 0.870 / 0.869 | | | 0.869 / 0.869 | |

Table S7: A result table summarizing the multiple linear regression results for the per-question evaluation ranks (1 to 7, higher is better) with respect to question type (qtype), models (model, with "DS32B" as the reference), disciplines (subject, with 01.AE as the reference), difficulty (with easy as the reference), rare diseases, diagnosis, treatment and operational procedures. The reference levels for the last covariates are all [no]. The 9,856 records cover all independent evaluators' assessments on the model responses' medical strength and quality.

(A)

| | Model 4a: Ascending rank | | | Model 4b: Ascending rank | | |
|---|---|---|---|---|---|---|
| Predictors | Estimates | 95% C.I. | *p* | Estimates | 95% C.I. | *p* |
| **Models** | | | | | | |
| model [DS70B] | 2.47 | 2.28, 2.66 | **<0.001** | 2.48 | 2.31, 2.64 | **<0.001** |
| model [DS32B] | 2.78 | 2.60, 2.97 | < 0.001 | 2.79 | 2.63, 2.96 | **<0.001** |
| model [ChatGPT-4oL] | 4.22 | 4.03, 4.41 | **<0.001** | 4.17 | 4.01, 4.34 | **<0.001** |
| model [QWEN32b] | 4.23 | 4.05, 4.42 | **<0.001** | 4.19 | 4.03, 4.36 | **<0.001** |
| model [DS-r1 671B] | 4.44 | 4.26, 4.63 | **<0.001** | 4.42 | 4.26, 4.59 | **<0.001** |
| model [QWENMAX] | 4.71 | 4.53, 4.90 | **<0.001** | 4.66 | 4.50, 4.83 | **<0.001** |
| model [Gemini2F] | 5.14 | 4.95, 5.33 | **<0.001** | 5.25 | 5.09, 5.41 | **<0.001** |
| **Disciplines** | | | | | | |
| subject [01b.AE] | 0.00 | -0.21, 0.21 | 1.000 | 0.00 | -0.17, 0.18 | 0.981 |
| subject [02.Cardio] | 0.00 | -0.18, 0.18 | 1.000 | 0.00 | -0.17, 0.18 | 0.971 |
| subject [03.CardiacSurg] | 0.00 | -0.15, 0.15 | 1.000 | 0.00 | -0.14, 0.15 | 0.952 |
| subject [04.Neural] | 0.00 | -0.16, 0.16 | 1.000 | 0.00 | -0.13, 0.14 | 0.972 |
| subject [05.Nephro] | 0.00 | -0.15, 0.15 | 1.000 | 0.00 | -0.14, 0.14 | 0.961 |
| subject [06.Ortho] | 0.00 | -0.16, 0.16 | 1.000 | 0.00 | -0.14, 0.15 | 0.955 |
| subject [07.ICU] | 0.00 | -0.16, 0.16 | 1.000 | 0.00 | -0.14, 0.15 | 0.960 |
| subject [08.Pedi] | 0.00 | -0.17, 0.17 | 1.000 | 0.00 | -0.13, 0.14 | 0.957 |
| subject [09.Prenatal] | 0.00 | -0.21, 0.21 | 1.000 | 0.00 | -0.19, 0.20 | 0.978 |
| subject [10.Neonatal] | 0.00 | -0.17, 0.17 | 1.000 | 0.00 | -0.14, 0.14 | 0.955 |
| subject [11.Resp] | 0.00 | -0.17, 0.17 | 1.000 | 0.00 | -0.15, 0.16 | 0.966 |
| subject [12.PediSurg] | 0.00 | -0.21, 0.21 | 1.000 | 0.00 | -0.16, 0.16 | 0.969 |
| subject [13.Rare] | 0.00 | -0.21, 0.21 | 1.000 | 0.00 | -0.19, 0.20 | 0.981 |
| **Question characteristics** | | | | | | |
| qtype [realCases] | -0.00 | -0.10, 0.10 | 1.000 | 0.00 | -0.09, 0.09 | 0.966 |
| difficulty [medium] | 0.00 | -0.09, 0.09 | 1.000 | -0.00 | -0.08, 0.08 | 0.981 |
| difficulty [hard] | -0.00 | -0.14, 0.14 | 1.000 | -0.00 | -0.13, 0.12 | 0.985 |
| rare [yes] | -0.00 | -0.15, 0.15 | 1.000 | -0.00 | -0.13, 0.13 | 0.996 |
| diagnosis [yes] | -0.00 | -0.12, 0.12 | 1.000 | 0.00 | -0.10, 0.11 | 0.969 |
| treatment [yes] | 0.00 | -0.10, 0.10 | 1.000 | -0.00 | -0.09, 0.09 | 0.993 |
| procedure [yes] | 0.00 | -0.09, 0.09 | 1.000 | 0.00 | -0.08, 0.08 | 0.999 |
| qlen | 0.00 | -0.06, 0.06 | 1.000 | -0.00 | -0.06, 0.06 | 0.974 |
| Observations | | 9,856 | | | 11,995 | |
| $R^2$ / $R^2$ adjusted | | 0.867 / 0.866 | | | 0.866 / 0.866 | |

(B) ANOVA and AIC model comparison between Model 3a and Model 4a.

| Models | Residual DF | RSS | DF | Sum of Sq | F-value | *p*-value | AIC |
|---|---|---|---|---|---|---|---|
| Model 3a | 9,820 | 25,539 | - | - | - | - | 37209.90 |
| Model 4a | 9,829 | 24,979 | -9 | -559.99 | 24.461 | $< \mathbf{2.2 \times 10^{-16}}$ | 37410.41 |

Table S8: Linear mixed-effects modeling of raw strength scores, on setA and setABC. ICC: Intraclass Correlation Coefficient. References: rlenrank [1.0], qtype: OSCE, subject: 01a.AE, difficulty: easy, rare: no, diagnosis: no, treatment: no, procedure: no. Model LME1 and Model LME2 were fitted on SetA and SetABC, respectively.

| | **Model LME1: strength** | | | **Model LME2: strength** | | |
|---|---|---|---|---|---|---|
| **Fixed-effects** predictors | Estimate | 95% CI | p-value | Estimate | 95% CI | p-value |
| **Models** | | | | | | |
| model [DS70B] | 5.28 | 4.30, 6.25 | <**0.001** | 5.06 | 4.28, 5.84 | <**0.001** |
| model [DS32B] | 5.51 | 4.53, 6.48 | <**0.001** | 5.30 | 4.52, 6.07 | <**0.001** |
| model [QWEN32b] | 5.76 | 4.79, 6.74 | <**0.001** | 5.53 | 4.75, 6.31 | <**0.001** |
| model [ChatGPT-4oL] | 5.91 | 4.94, 6.89 | <**0.001** | 5.68 | 4.90, 6.46 | <**0.001** |
| model [Gemini2F] | 5.93 | 4.94, 6.91 | <**0.001** | 5.76 | 4.97, 6.55 | <**0.001** |
| model [QWENMAX] | 5.95 | 4.97, 6.93 | <**0.001** | 5.71 | 4.93, 6.50 | <**0.001** |
| model [DS-r1 671B] | 6.05 | 5.07, 7.03 | <**0.001** | 5.82 | 5.04, 6.61 | <**0.001** |
| rlenrank [1.5] | 0.42 | -0.21, 1.06 | 0.191 | -0.03 | -0.61, 0.56 | 0.929 |
| rlenrank [2] | 0.17 | 0.10, 0.24 | <**0.001** | 0.17 | 0.10, 0.24 | <**0.001** |
| rlenrank [3] | 0.40 | 0.29, 0.52 | <**0.001** | 0.43 | 0.32, 0.54 | <**0.001** |
| rlenrank [3.5] | 1.01 | -0.09, 2.11 | 0.073 | 1.11 | 0.16, 2.07 | 0.023 |
| rlenrank [4] | 0.53 | 0.41, 0.64 | <**0.001** | 0.55 | 0.43, 0.66 | <**0.001** |
| rlenrank [4.5] | 0.99 | -0.11, 2.10 | 0.077 | 1.03 | -0.13, 2.19 | 0.081 |
| rlenrank [5] | 0.61 | 0.48, 0.73 | <**0.001** | 0.60 | 0.49, 0.72 | <**0.001** |
| rlenrank [6] | 0.67 | 0.54, 0.80 | <**0.001** | 0.69 | 0.57, 0.82 | <**0.001** |
| rlenrank [7] | 0.92 | 0.75, 1.10 | <**0.001** | 0.97 | 0.80, 1.15 | <**0.001** |
| **Disciplines** | | | | | | |
| subject [01b.AEq20] | -0.18 | -1.54, 1.19 | 0.798 | -0.18 | -1.26, 0.91 | 0.746 |
| subject [02.Cardio] | 0.29 | -1.07, 1.64 | 0.677 | 0.45 | -0.74, 1.65 | 0.460 |
| subject [03.CardiacSurg] | 0.80 | -0.55, 2.15 | 0.243 | 0.96 | -0.23, 2.15 | 0.112 |
| subject [04.Neural] | 0.65 | -0.70, 2.01 | 0.343 | 1.30 | 0.23, 2.37 | 0.017 |
| subject [05.Nephro] | 0.03 | -1.21, 1.27 | 0.965 | 0.19 | -0.88, 1.26 | 0.727 |
| subject [06.Ortho] | 0.21 | -1.14, 1.56 | 0.759 | 0.38 | -0.81, 1.57 | 0.530 |
| subject [07.ICU] | -0.85 | -2.20, 0.50 | 0.215 | -0.67 | -1.86, 0.52 | 0.267 |
| subject [08.Pedi] | 0.09 | -1.26, 1.45 | 0.891 | 0.10 | -0.97, 1.17 | 0.849 |
| subject [09.Prenatal] | 1.57 | 0.21, 2.94 | 0.024 | 1.72 | 0.52, 2.92 | 0.005 |
| subject [10.Neonatal] | 0.18 | -1.17, 1.54 | 0.789 | 0.40 | -0.67, 1.47 | 0.460 |
| subject [11.Resp] | 0.68 | -0.67, 2.04 | 0.321 | 0.85 | -0.34, 2.04 | 0.161 |
| subject [12.PediSurg] | -0.91 | -2.56, 0.74 | 0.280 | -0.46 | -1.65, 0.73 | 0.450 |
| subject [13.Rare] | 0.22 | -1.04, 1.47 | 0.736 | 0.34 | -0.75, 1.43 | 0.538 |
| **Question characteristics** | | | | | | |
| qtype [realCases] | -0.02 | -0.15, 0.12 | 0.825 | -0.02 | -0.16, 0.11 | 0.733 |
| difficulty [medium] | 0.11 | -0.01, 0.24 | 0.081 | 0.10 | -0.03, 0.22 | 0.119 |
| difficulty [hard] | 0.03 | -0.16, 0.22 | 0.743 | 0.01 | -0.17, 0.20 | 0.894 |
| rare [yes] | 0.12 | -0.07, 0.32 | 0.214 | 0.16 | -0.04, 0.35 | 0.111 |
| diagnosis [yes] | -0.05 | -0.21, 0.11 | 0.533 | -0.01 | -0.17, 0.15 | 0.907 |
| treatment [yes] | 0.04 | -0.09, 0.18 | 0.538 | 0.04 | -0.09, 0.18 | 0.524 |
| procedure [yes] | 0.03 | -0.10, 0.15 | 0.686 | 0.03 | -0.09, 0.16 | 0.614 |
| qlen | -0.07 | -0.16, 0.02 | 0.119 | -0.06 | -0.15, 0.03 | 0.209 |
| **Random Effects** | | | | | | |
| Residual variance ($\sigma^2$) | | 0.96 | | | 1.07 | |
| QID variance ($\tau^2_{\text{QID}}$) | | 0.27 | | | 0.28 | |
| Evaluator variance ($\tau^2_{\text{evaluator}}$) | | 0.46 | | | 0.43 | |
| ICC | | 0.43 | | | 0.40 | |
| N (evaluator) | | 29 | | | 35 | |
| N (QID) | | 685 | | | 685 | |
| Observations | | 9,856 | | | 11,995 | |
| Marginal $R^2$ / Conditional $R^2$ | | 0.260 / 0.580 | | | 0.271 / 0.561 | |

Table S9: Comparison of linear model (LM) and linear mixed-effects model (LME).

| Model | npar | AIC | BIC | logLik | -2 logLik | Chisq | Df | Pr(> $\chi^2$) |
|---|---|---|---|---|---|---|---|---|
| Model 2a | 38 | 31643 | 31916 | -15783 | 31567 | | | |
| Model LME1 | 39 | 28860 | 29140 | -14391 | 28782 | 2785 | 2 | $< 2.2 \times 10^{-16}$ *** |

Significance codes: $^{***}p < 0.001$

Table S10: Fixed effects estimates and odds ratios with 95% confidence intervals from logistic mixed model predicting excellence (score $\geq$ 8), adjusted for evaluator random effects

| Predictor | Estimate | OR | 95% CI (OR) | $p$ |
|---|---|---|---|---|
| **(Intercept)** | -6.47 | 0.0016 | [0.0003, 0.0090] | <**0.001** |
| *Model (vs. reference [DS32B])* | | | | |
| DS70B | -0.18 | 0.83 | [0.61, 1.14] | 0.261 |
| QWEN32b | 1.40 | 4.06 | [3.08, 5.34] | <**0.001** |
| DS-r1 671B | 1.46 | 4.32 | [3.30, 5.66] | <**0.001** |
| ChatGPT-4oL | 1.49 | 4.43 | [3.39, 5.78] | <**0.001** |
| QWENMAX | 2.01 | 7.47 | [5.65, 9.88] | <**0.001** |
| Gemini2F | 2.50 | 12.16 | [9.21, 16.06] | <**0.001** |
| *Disciplines (vs. reference 01a.AE)* | | | | |
| ICU | 0.00 | 1.00 | [0.06, 16.47] | 0.998 |
| Pediatric Surgery | -1.21 | 0.30 | [0.01, 5.72] | 0.537 |
| AEq20 | -1.38 | 0.25 | [0.01, 4.34] | 0.448 |
| Pediatrics | 2.28 | 9.76 | [0.71, 133.58] | 0.095 |
| Neurology | 2.57 | 13.03 | [0.98, 173.15] | 0.067 |
| Rare Diseases | 2.93 | 18.74 | [1.45, 242.64] | **0.021** |
| Orthopedics | 3.06 | 21.41 | [1.70, 270.00] | **0.024** |
| Nephrology | 3.10 | 22.17 | [1.94, 253.11] | **0.013** |
| Cardiology | 3.77 | 43.27 | [3.32, 563.59] | **0.006** |
| Respiratory | 3.89 | 48.89 | [3.74, 639.61] | **0.004** |
| Cardiac Surgery | 4.06 | 57.87 | [4.46, 750.70] | **0.003** |
| Neonatology | 4.37 | 79.31 | [6.07, 1036.21] | <**0.001** |
| Prenatal | 5.80 | 330.36 | [25.06, 4353.63] | <**0.001** |

Table S11: Fixed effects estimates and odds ratios with 95% confidence intervals from logistic mixed model predicting excellence (score $\geq$ 8), adjusted for evaluator random effects

| | **Excellence (raw score $\geq$ 8)** | | |
|---|---|---|---|
| **Fixed-effects** predictors | O.R. est. | 95% C.I. | $p$ |
| (Intercept) | 0.00 | 0.00, 0.01 | <**0.001** |
| model [DS70B] | 0.80 | 0.58, 1.10 | 0.173 |
| model [QWEN32b] | 1.90 | 1.22, 2.94 | **0.004** |
| model [ChatGPT-4oL] | 2.44 | 1.61, 3.71 | <**0.001** |
| model [Gemini2F] | 2.45 | 1.40, 4.27 | **0.002** |
| model [QWENMAX] | 2.48 | 1.56, 3.93 | <**0.001** |
| model [DS-r1 671B] | 2.88 | 1.90, 4.38 | <**0.001** |
| rlenrank [1.5] | 2.60 | 0.18, 38.49 | 0.486 |
| rlenrank [2] | 1.40 | 1.02, 1.94 | **0.038** |
| rlenrank [3] | 1.49 | 0.95, 2.32 | 0.081 |
| rlenrank [3.5] | 21.09 | 0.15, 2886.30 | 0.224 |
| rlenrank [4] | 2.11 | 1.34, 3.30 | **0.001** |
| rlenrank [4.5] | 0.00 | 0.00, Inf | 0.975 |
| rlenrank [5] | 2.76 | 1.74, 4.37 | <**0.001** |
| rlenrank [6] | 4.15 | 2.57, 6.72 | <**0.001** |
| rlenrank [7] | 6.36 | 3.58, 11.30 | <**0.001** |
| subject [01b.AE] | 0.25 | 0.01, 9.10 | 0.452 |
| subject [12.PediSurg] | 0.30 | 0.01, 14.43 | 0.544 |
| subject [07.ICU] | 1.02 | 0.06, 17.06 | 0.988 |
| subject [08.Pedi] | 9.90 | 0.67, 147.15 | 0.096 |
| subject [04.Neural] | 13.07 | 0.82, 209.28 | 0.069 |
| subject [13.Rare] | 19.20 | 1.57, 234.82 | **0.021** |
| subject [06.Ortho] | 21.89 | 1.48, 323.38 | **0.025** |
| subject [05.Nephro] | 22.64 | 1.91, 267.71 | **0.013** |
| subject [02.Cardio] | 44.67 | 3.03, 657.94 | **0.006** |
| subject [11.Resp] | 50.24 | 3.42, 737.53 | **0.004** |
| subject [03.CardiacSurg] | 59.75 | 4.08, 875.90 | **0.003** |
| subject [10.Neonatal] | 82.35 | 5.61, 1209.21 | **0.001** |
| subject [09.Prenatal] | 352.50 | 24.02, 5173.71 | <**0.001** |
| **Random Effects** | | | |
| $\sigma^2$ | | 3.29 | |
| $\tau_{00 \text{ evaluator}}$ | | 1.76 | |
| ICC | | 0.35 | |
| $N_{\text{evaluator}}$ | | 29 | |
| Observations | | 9856 | |
| Marginal $R^2$ / Conditional $R^2$ | | 0.483 / 0.663 | |

Table S12: Fixed effects estimates and odds ratios (OR) with 95% confidence intervals from generalized linear mixed model predicting failure (score $\leq 4$), adjusted for evaluator random effects.

| | **Incompetence (raw score $\leq$ 4)** | | | |
|---|---|---|---|---|
| Predictor | Estimate | OR | 95% C.I. (OR) | $p$ |
| (Intercept) | -4.69 | 0.01 | 0.0, 0.04 | <**0.001** |
| **Models** (reference: DS32B) | | | | |
| DS70B | 0.67 | 1.95 | 1.55, 2.47 | <**0.001** |
| Qwen32B | -1.08 | 0.34 | 0.25, 0.47 | <**0.001** |
| ChatGPT-4oL | -1.39 | 0.25 | 0.18, 0.35 | <**0.001** |
| QwenMAX | -1.45 | 0.23 | 0.17, 0.31 | <**0.001** |
| DS-r1 671B | -1.56 | 0.21 | 0.15, 0.30 | <**0.001** |
| Gemini2F | -1.82 | 0.16 | 0.11, 0.22 | <**0.001** |
| **Disciplines** (reference: 01a.AE) | | | | |
| 01b.AE | -0.72 | 0.49 | 0.03, 7.11 | 0.611 |
| Prenatal | 0.41 | 1.50 | 0.18, 12.60 | 0.705 |
| Pediatrics | 1.51 | 4.51 | 0.56, 36.29 | 0.143 |
| Respiratory | 1.95 | 7.04 | 0.99, 50.23 | 0.055 |
| Rare Diseases | 2.10 | 8.20 | 1.31, 51.21 | **0.026** |
| Cardiac Surgery | 2.20 | 9.00 | 1.11, 72.77 | **0.030** |
| Neurology | 2.60 | 13.54 | 1.83, 100.06 | **0.010** |
| Orthopedics | 2.68 | 14.61 | 1.95, 109.52 | **0.008** |
| Nephrology | 2.72 | 15.19 | 2.45, 94.17 | **0.004** |
| Cardiology | 3.06 | 21.25 | 2.92, 154.60 | **0.002** |
| Neonatology | 3.59 | 36.43 | 4.85, 273.43 | <**0.001** |
| ICU | 3.70 | 40.58 | 5.42, 303.78 | <**0.001** |
| Pediatric Surgery | 3.76 | 43.02 | 4.97, 372.56 | **0.002** |

Table S13: Fixed effects estimates and odds ratios with 95% confidence intervals from logistic mixed model predicting failure (score $\leq 4$), adjusted for evaluator random effects and response lengths. Note that there were 714 instances of incompetence, causing fitting of the full model below difficult.

| | **Incompetence (raw score $\leq$ 4)** | | | |
|---|---|---|---|---|
| **Fixed-effects** predictors | Estimate | O.R. est. | 95% C.I. | $p$ |
| (Intercept) | -4.60 | 0.01 | 0.00, 0.02 | <**0.001** |
| **Models (ref: 32)** | | | | |
| DS70B | 0.68 | 1.98 | 1.57, 2.49 | <**0.001** |
| Gemini2F | -0.10 | 0.90 | 0.39, 2.09 | 0.808 |
| QWEN32b | -0.13 | 0.88 | 0.53, 1.45 | 0.613 |
| QWENMAX | -0.44 | 0.64 | 0.36, 1.14 | 0.125 |
| ChatGPT-4oL | -0.62 | 0.54 | 0.34, 0.88 | **0.008** |
| DS-r1 671B | -0.82 | 0.44 | 0.27, 0.71 | <**0.001** |
| **Response length (ref: 1)** | | | | |
| rlenrank 1.5 | -1.49 | 0.23 | 0.02, 2.56 | 0.210 |
| rlenrank 2 | -0.12 | 0.88 | 0.70, 1.10 | 0.293 |
| rlenrank 3 | -0.81 | 0.45 | 0.29, 0.69 | <**0.001** |
| rlenrank 3.5 | -9.97 | 0.00 | 0.00, 0.00 | 0.971 |
| rlenrank 4 | -1.00 | 0.37 | 0.23, 0.60 | <**0.001** |
| rlenrank 4.5 | -13.23 | 0.00 | 0.00, 0.00 | 0.978 |
| rlenrank 5 | -1.16 | 0.31 | 0.18, 0.51 | <**0.001** |
| rlenrank 6 | -1.11 | 0.33 | 0.19, 0.58 | <**0.001** |
| rlenrank 7 | -1.95 | 0.14 | 0.06, 0.33 | <**0.001** |
| **Disciplines (ref: AE set1)** | | | | |
| AE (set2) | -0.63 | 0.53 | 0.03, 9.47 | 0.674 |
| Prenatal | 0.41 | 1.51 | 0.18, 12.83 | 0.706 |
| Pediatrics | 1.53 | 4.60 | 0.61, 34.65 | 0.140 |
| Respiratory | 1.97 | 7.17 | 1.00, 51.29 | 0.054 |
| Rare Diseases | 2.12 | 8.36 | 1.31, 53.26 | **0.026** |
| Cardiac Surgery | 2.21 | 9.17 | 1.10, 76.80 | **0.030** |
| Neurology | 2.65 | 14.21 | 1.76, 114.92 | **0.009** |
| Orthopedics | 2.70 | 14.90 | 1.98, 112.10 | **0.007** |
| Nephrology | 2.74 | 15.53 | 2.32, 103.89 | **0.003** |
| Cardiology | 3.06 | 21.43 | 2.91, 157.81 | **0.002** |
| Neonatology | 3.62 | 37.22 | 4.99, 277.64 | <**0.001** |
| ICU | 3.74 | 42.27 | 5.63, 317.25 | <**0.001** |
| Pediatric Surgery | 3.79 | 44.10 | 4.82, 403.84 | **0.002** |

Table S14: Pearson correlation matrix between LLMs when they were asked to score the responses, based on 18,768 data-points from scores on 4,692 responses (belonging to 675 questions). LLMs were asked to rate all seven responses per question, which included their own responses in the previous round.

| | **LLMs (as evaluators)** | | | |
|---|---|---|---|---|
| **LLMs (as evaluators)** | DeepSeek r1 | Gemini 2 | GPT-4o | Qwen Max |
| DeepSeek r1 | 1.000 | 0.795 | 0.744 | 0.797 |
| Gemini 2 | 0.795 | 1.000 | 0.709 | 0.770 |
| GPT-4o | 0.744 | 0.709 | 1.000 | 0.746 |
| Qwen Max | 0.797 | 0.770 | 0.746 | 1.000 |

Table S15: Linear regression results for LLM-rated scores.

| | Strength given by LLMs | | |
|---|---|---|---|
| Predictors | Estimate | 95% C.I. | $p$ |
| **Models** | | | |
| DS70B | 5.2231 | [5.1388, 5.3073] | <**0.001** |
| DS32B | 5.9028 | [5.8177, 5.9879] | <**0.001** |
| QWEN32b | 6.5876 | [6.4752, 6.6999] | <**0.001** |
| DS-r1 671B | 6.9826 | [6.8737, 7.0915] | <**0.001** |
| QWENMAX | 6.9943 | [6.8777, 7.1109] | <**0.001** |
| Gemini2F | 7.1844 | [7.0396, 7.3291] | <**0.001** |
| ChatGPT-4oL | 7.2192 | [7.1104, 7.3280] | <**0.001** |
| **Evaluator** (ref: DeepSeek r1 671B) | | | |
| Gemini2F | 0.5869 | [0.5491, 0.6248] | <**0.001** |
| ChatGPT-4oL | 1.0035 | [0.9657, 1.0413] | <**0.001** |
| QWENMAX | 0.8301 | [0.7923, 0.8679] | <**0.001** |
| **Response-length** (ref: rlenrank [1]) | | | |
| rlenrank [1.5] | 0.1625 | [-0.2123, 0.5373] | 0.395 |
| rlenrank [2] | 0.3039 | [0.2528, 0.3550] | <**0.001** |
| rlenrank [3] | 0.6720 | [0.5928, 0.7512] | <**0.001** |
| rlenrank [3.5] | 0.6750 | [0.0237, 1.3263] | **0.042** |
| rlenrank [4] | 0.7665 | [0.6832, 0.8498] | <**0.001** |
| rlenrank [4.5] | 1.2663 | [0.6158, 1.9168] | <**0.001** |
| rlenrank [5] | 0.8988 | [0.8120, 0.9856] | <**0.001** |
| rlenrank [6] | 0.8990 | [0.8089, 0.9891] | <**0.001** |
| rlenrank [7] | 1.0621 | [0.9367, 1.1875] | <**0.001** |
| **Disciplines** (ref: A.E.) | | | |
| subject [02.Cardio] | 0.0374 | [-0.0360, 0.1108] | 0.324 |
| subject [03.CardiacSurg] | 0.0502 | [-0.0122, 0.1126] | 0.116 |
| subject [04.Neural] | 0.1511 | [0.0840, 0.2181] | <**0.001** |
| subject [05.Nephro] | 0.0890 | [0.0206, 0.1575] | **0.011** |
| subject [06.Ortho] | -0.0269 | [-0.0911, 0.0373] | 0.414 |
| subject [07.ICU] | 0.0489 | [-0.0141, 0.1119] | 0.134 |
| subject [08.Pedi] | 0.1091 | [0.0414, 0.1767] | **0.002** |
| subject [09.Prenatal] | 0.0304 | [-0.0547, 0.1155] | 0.483 |
| subject [10.Neonatal] | 0.0071 | [-0.0614, 0.0756] | 0.839 |
| subject [11.Resp] | 0.1329 | [0.0653, 0.2005] | <**0.001** |
| subject [12.PediSurg] | -0.0041 | [-0.0731, 0.0649] | 0.907 |
| subject [13.Rare] | 0.0335 | [-0.0586, 0.1255] | 0.475 |
| **Question characteristics** | | | |
| qtype [realCases] | 0.0001 | [-0.0403, 0.0406] | 0.995 |
| difficult [medium] | -0.0487 | [-0.0871, -0.0103] | **0.013** |
| difficult [hard] | -0.1197 | [-0.1766, -0.0627] | <**0.001** |
| rare [yes] | 0.0358 | [-0.0214, 0.0930] | 0.223 |
| diagnosis [yes] | -0.0294 | [-0.0772, 0.0185] | 0.229 |
| treatment [yes] | 0.0473 | [0.0066, 0.0879] | **0.022** |
| procedure [yes] | 0.0169 | [-0.0209, 0.0546] | 0.382 |
| qlen (per '000 characters) | -0.0248 | [-0.0518, 0.0021] | 0.072 |

Residual standard error: 0.9285 on 18,453 degrees of freedom.
Multiple $R^2$: 0.9865, Adjusted $R^2$: 0.9865.
F-statistic: 34,540 on 39 and 18,453 DF, p-value: $< 2.2 \times 10^{-16}$.

#### Supplementary References

[1] Liu, Xihan, Jie Feng, Chenmian Liu, Ran Chu, Ming Lv, Ning Zhong, Yuchun Tang, Li Li, and Kun Song. "Medical education systems in China: development, status, and evaluation." Academic Medicine 98, no. 1 (2023): 43-49.

[2] Vaswani, Ashish, Noam Shazeer, Niki Parmar, Jakob Uszkoreit, Llion Jones, Aidan N. Gomez, Łukasz Kaiser, and Illia Polosukhin. "Attention is all you need." Advances in neural information processing systems 30 (2017).

[3] Thirunavukarasu, Arun James, Darren Shu Jeng Ting, Kabilan Elangovan, Laura Gutierrez, Ting Fang Tan, and Daniel Shu Wei Ting. "Large language models in medicine." Nature medicine 29, no. 8 (2023): 1930-1940.

[4] Singhal, Karan, Shekoofeh Azizi, Tao Tu, S. Sara Mahdavi, Jason Wei, Hyung Won Chung, Nathan Scales et al. "Large language models encode clinical knowledge." Nature 620, no. 7972 (2023): 172-180.

[5] Mehnen, Lars, Stefanie Gruarin, Mina Vasileva, and Bernhard Knapp. "ChatGPT as a medical doctor? A diagnostic accuracy study on common and rare diseases." MedRxiv (2023): 2023-04.

[6] Lim, Zhi Wei, Krithi Pushpanathan, Samantha Min Er Yew, Yien Lai, Chen-Hsin Sun, Janice Sing Harn Lam, David Ziyou Chen et al. "Benchmarking large language models' performances for myopia care: a comparative analysis of ChatGPT-3.5, ChatGPT-4.0, and Google Bard." EBioMedicine 95 (2023).

[7] Truhn, Daniel, Jorge S. Reis-Filho, and Jakob Nikolas Kather. "Large language models should be used as scientific reasoning engines, not knowledge databases." Nature medicine 29, no. 12 (2023): 2983-2984.

[8] Goodman, Rachel S., J. Randall Patrinely, Cosby A. Stone, Eli Zimmerman, Rebecca R. Donald, Sam S. Chang, Sean T. Berkowitz et al. "Accuracy and reliability of chatbot responses to physician questions." JAMA network open 6, no. 10 (2023): e2336483-e2336483.

[9] Zelin, Charlotte, Wendy K. Chung, Mederic Jeanne, Gongbo Zhang, and Chunhua Weng. "Rare disease diagnosis using knowledge guided retrieval augmentation for ChatGPT." Journal of Biomedical Informatics 157 (2024): 104702.

[10] Du, Xinsong, John Novoa-Laurentiev, Joseph M. Plasek, Ya-Wen Chuang, Liqin Wang, Gad A. Marshall, Stephanie K. Mueller et al. "Enhancing early detection of cognitive decline in the elderly: a comparative study utilizing large language models in clinical notes." EBioMedicine 109 (2024).

[11] Chen, Xuanzhong, Xiaohao Mao, Qihan Guo, Lun Wang, Shuyang Zhang, and Ting Chen. "RareBench: Can LLMs Serve as Rare Diseases Specialists?." In Proceedings of the 30th ACM SIGKDD Conference on Knowledge Discovery and Data Mining, pp. 4850-4861. 2024.

[12] Gallifant, Jack, Majid Afshar, Saleem Ameen, Yindalon Aphinyanaphongs, Shan Chen, Giovanni Cacciamani, Dina Demner-Fushman et al. "The TRIPOD-LLM reporting guideline for studies using large language models." Nature Medicine (2025): 1-10.

[13] Goh, Ethan, Robert Gallo, Jason Hom, Eric Strong, Yingjie Weng, Hannah Kerman, Joséphine A. Cool et al. "Large language model influence on diagnostic reasoning: a randomized clinical trial." JAMA Network Open 7, no. 10 (2024): e2440969-e2440969.

[14] Guo, Daya, Dejian Yang, Haowei Zhang, Junxiao Song, Ruoyu Zhang, Runxin Xu, Qihao Zhu et al. "Deepseek-r1: Incentivizing reasoning capability in llms via reinforcement learning." arXiv preprint arXiv:2501.12948 (2025).

[15] Svetozarević, Mihailo, Isidora Janković, Stevo Lukić, and Sonja Janković. "Standards for Use of LLM in Medical Diagnosis." (2024).

[16] Ong, Jasmine Chiat Ling, Shelley Yin-Hsi Chang, Wasswa William, Atul J. Butte, Nigam H. Shah, Lita Sui Tjien Chew, Nan Liu et al. "Medical ethics of large language models in medicine." NEJM AI 1, no. 7 (2024): AIra2400038.

[17] Li, Jin, Yiyan Deng, Qi Sun, Junjie Zhu, Yu Tian, Jingsong Li, and Tingting Zhu. "Benchmarking large language models in evidence-based medicine." IEEE Journal of Biomedical and Health Informatics (2024).

[18] McGrath, Scott P., Beth A. Kozel, Sara Gracefo, Nykole Sutherland, Christopher J. Danford, and Nephi Walton. "A comparative evaluation of ChatGPT 3.5 and ChatGPT 4 in responses to selected genetics questions." Journal of the American Medical Informatics Association 31, no. 10 (2024): 2271-2283.

[19] Rydzewski, Nicholas R., Deepak Dinakaran, Shuang G. Zhao, Eytan Ruppin, Baris Turkbey, Deborah E. Citrin, and Krishnan R. Patel. "Comparative evaluation of LLMs in clinical oncology." Nejm Ai 1, no. 5 (2024): AIoa2300151.

[20] Kweon, Sunjun, Jiyoun Kim, Heeyoung Kwak, Dongchul Cha, Hangyul Yoon, Kwang Kim, Jeewon Yang, Seunghyun Won, and Edward Choi. "EHRNoteQA: An LLM Benchmark for Real-World Clinical Practice Using Discharge Summaries." Advances in Neural Information Processing Systems 37 (2025): 124575-124611.

[21] Goh, Ethan, Robert Gallo, Jason Hom, Eric Strong, Yingjie Weng, Hannah Kerman, Joséphine A. Cool et al. "Large language model influence on diagnostic reasoning: a randomized clinical trial." JAMA Network Open 7, no. 10 (2024): e2440969-e2440969.

[22] Bedi, Suhana, Yutong Liu, Lucy Orr-Ewing, Dev Dash, Sanmi Koyejo, Alison Callahan, Jason A. Fries et al. "Testing and evaluation of health care applications of large language models: a systematic review." JAMA (2024).

[23] Pieri, Sara, Sahal Shaji Mullappilly, Fahad Shahbaz Khan, Rao Muhammad Anwer, Salman Khan, Timothy Baldwin, and Hisham Cholakkal. "Bimedix: Bilingual medical mixture of experts llm." arXiv preprint arXiv:2402.13253 (2024).

[24] Chen, David, Ryan S. Huang, Jane Jomy, Philip Wong, Michael Yan, Jennifer Croke, Daniel Tong, Andrew Hope, Lawson Eng, and Srinivas Raman. "Performance of Multimodal Artificial Intelligence Chatbots Evaluated on Clinical Oncology Cases." JAMA Network Open 7, no. 10 (2024): e2437711-e2437711.

[25] Johri, Shreya, Jaehwan Jeong, Benjamin A. Tran, Daniel I. Schlessinger, Shannon Wongvibulsin, Leandra A. Barnes, Hong-Yu Zhou et al. "An evaluation framework for clinical use of large language models in patient interaction tasks." Nature Medicine (2025): 1-10.

[26] Flores-Gouyonnet, Jaime, María C. Cuéllar-Gutiérrez, Gabriel Figueroa-Parra, Bradly Kimbrough, Elena K. Joerns, Erika Navarro-Mendoza, Cynthia

[27] S. Crowson et al. "Performance of large language models in rheumatology board-like questions: accuracy, quality, and safety." The Lancet Rheumatology (2025).

[28] Huo, Bright, Amy Boyle, Nana Marfo, Wimonchat Tangamornsuksan, Jeremy P. Steen, Tyler McKechnie, Yung Lee et al. "Large Language Models for Chatbot Health Advice Studies: A Systematic Review." JAMA Network Open 8, no. 2 (2025): e2457879-e2457879.

[29] Tordjman, Mickael, Zelong Liu, Murat Yuce, Valentin Fauveau, Yunhao Mei, Jerome Hadjadj, Ian Bolger et al. "Comparative benchmarking of the DeepSeek large language model on medical tasks and clinical reasoning." Nature Medicine (2025): 1-1.

[30] Sandmann, Sarah, Stefan Hegselmann, Michael Fujarski, Lucas Bickmann, Benjamin Wild, Roland Eils, and Julian Varghese. "Benchmark evaluation of DeepSeek large language models in clinical decision-making." Nature Medicine (2025): 1-1.
